# Influence of the Built Environment on Airflow, Contamination, and Infection in the Operating Room: A Systematic Literature Review

**DOI:** 10.1101/2022.07.20.22277856

**Authors:** Shirley M. Klimkiewicz, Molly E. Gallagher, Anastasia S. Lambrou, Oluwaferanmi E. Adeyemo, Amy M. Andrelchik, Kavita Braun, Madeline B. Ford, Terrence J. Garcia, Stephanie Ku, Kaitlin Rainwater-Lovett, Julee A. Rendon, Sophia H. Oluic, Steven L. Patterson, Joshua Yoon, Alexander J. Yuan, Winnie Wang, Lucy Carruth, Brian Damit

## Abstract

**Background:** Healthcare-associated infections (HAIs) constitute a significant financial strain on healthcare systems across the world, with surgical site infections (SSIs) being the costliest form. Despite the existence of diverse sources of infection in the operating room (OR), current literature focuses on human and procedural sources of contamination that could lead to an infection. Comparatively, the OR built environment is understudied as a potential disease transmission interface between the environment, patients, and surgical staff. This systematic literature review aims to investigate how the physical characteristics and components of the built environment impact airflow, infection risk, aerosols, particle counts, contamination, and pathogens in operating rooms.

**Methods and Findings:** Literature searches were conducted in the PubMed and Web of Science Core Collection databases on December 21, 2020, ultimately retrieving 2,965 articles after duplicates were removed. During abstract screening, all abstracts were independently reviewed by two authors and conflicts were resolved by a third author. All articles published since January 1, 2010, that reported primary data investigating an aspect of the built environment inside an OR in relation to airflow, contamination, and/or infection for which the full text in English was available were included. This resulted in the inclusion of 138 articles, which includes studies conducted in ORs during active surgeries, computer modeling studies, and simulations in which a real OR was used for a mock surgical procedure. Six major built environment categories were identified based on the collected literature: OR layout, disinfection systems, surgical lights, doors, ventilation, and portable airflow devices. A survey created on Qualtrics software was used to record the aspect of the built environment and the outcome of each study, as well as the relationship between the two.

**Conclusions:** While OR ventilation has been studied extensively, the OR built environment as a whole is understudied in relation to airflow, contamination, and infection. The current literature is inconsistent in both its findings and subsequent recommendations, making it difficult to inform hospital design in the context of SSIs. No articles were identified that discussed respiratory infection transmission in the OR, and very few addressed healthcare worker (HCW) safety in relation to the OR built environment. The significant discrepancies in the literature identified in this review highlight the need for future studies that assess the quality and bias of these studies before firm recommendations can be made. Future work should also focus on addressing the lack information regarding respiratory infection transmission in the OR, especially in the context of HCW safety.

## 1. BACKGROUND

The influence of operating room (OR) air on the occurrence of healthcare-associated infections (HAIs) has been studied since the 1800s. In 1867, the surgeon Sir Joseph Lister described “floating particles” responsible for the “septic property of the atmosphere” inside ORs.^1^ Following over a decade of studying wound contamination and various antiseptic solutions, however, Lister walked back some of his previous work and concluded that any contaminants present in OR air could be disregarded during surgical practice.^2^ The discussion surrounding the influence of OR air on infections would not reemerge until 1933, when Dr. Frank Meleney spoke to the New York Surgical Society about his investigation into possible sources of infection in the OR.^3^ One of the possible sources identified by Dr. Meleney was the OR air, which he studied by placing exposed culture plates in several different ORs and observing bacterial growth. This led to the observation that, in the ORs supplied with filtered air, the bacterial settle rate was half that of the ORs without special ventilation.^3^ Dr. Meleney further observed that culture plates placed in active ORs grew ten times as many colonies as those placed in empty ORs, which he ascribed to the “people moving about and doors opening and closing.”^3^ Based on his report, it is clear that the idea of the OR as a complex environment that is sensitive to the activities performed in it has been explored for decades.

Since Dr. Meleney’s talk in 1933, significant progress has been made towards understanding and mitigating air contamination inside operating rooms. Today, healthcare professionals across the world generally agree that contamination in the OR air should be kept to a minimum. This is perhaps best exemplified by the fact that many developed countries have created recommendations and guidelines regarding ventilation systems used in ORs.^4–10^ The recommendations include ideal temperature and relative humidity ranges for inside the OR and in other healthcare facilities, as well as guidelines surrounding the preferred type of ventilation and the recommended air renewal rate. Despite the existence of such guidelines and the volume of research that preceded their creation, HAIs still occur and have detrimental effects on the healthcare system. In the United States, surgical site infections (SSIs) are the most costly type of HAI, costing about $3.3 billion and contributing an additional 1 million inpatient days annually.^11^ Due to their economic impact and mortality rate of 3%, these infections have been studied for decades, and information regarding SSI incidence for a variety of inpatient surgical procedures is reported every year by the Centers for Disease Control and Prevention (CDC) in their annual National and State Healthcare-Associated Infections Progress Report.^12^ This report provides data on several HAIs, including central-line associated bloodstream infections, ventilator-associated events, *Clostridium difficile* events, and SSIs associated with 39 different inpatient surgical procedure categories, all broken down by facility and procedure type.^12^ While the report is thorough in terms of the aforementioned HAIs and in tracking infections over time, it lacks any information regarding the transmission of respiratory infections in healthcare settings, including inside the OR. The onset of the coronavirus disease 2019 (COVID-19) pandemic brought about concerns regarding the transmission of this respiratory virus between patient and healthcare workers (HCWs) inside the OR. While widespread vaccination against severe acute respiratory syndrome coronavirus 2 (SARS-CoV-2) has helped mitigate these concerns, the pandemic brought attention to the fact that respiratory infection transmission is extremely understudied in all settings, including inside ORs.

The COVID-19 pandemic is not the only recent outbreak to highlight a critical need for this work. Since 2014, the relatively novel pathogen *Mycobacterium chimera* has been implicated in an unprecedented number of infections suffered by patients after undergoing heart surgery. Following multiple investigations into the outbreak, the origin of the infections was discovered to be heater-cooler units (HCUs) that had been contaminated with *Mycobacterium chimera*, likely during assembly at a production plant.^13^ The infections were traced to a specific HCU model, whose exhaust vent was capable of transmitting aerosols from the contaminated water tanks in the equipment. Several studies since conducted have determined that aerosols containing *Mycobacterium chimera* travelled from the exhaust vent of the HCUs to exposed parts of the body during surgery, leading to infection.^13^ While this outbreak has been largely contained, it serves to demonstrate how aerosolized pathogens in the OR pose a significant threat to public health. Additionally, this event reveals a gap in understanding how not only medical equipment such as HCUs but other aspects of the OR environment interact with the air.

As previously mentioned, airborne contamination in the OR has been studied for over a century by researchers around the world. The extensive body of information collected during these studies has led to implementation of ventilation and pressure requirements designed to keep “dirty” air out of the OR and continuously filter out contaminants produced inside of the OR during surgical procedures. While these strategies have generally shown to be effective at maintaining a degree of overall OR air cleanliness, the pathway of contaminants from their point of production inside the OR to an exhaust vent is poorly understood. Furthermore, although these strategies exhibit some effectiveness against contamination produced from day-to-day OR activities, the *Mycobacterium chimera* outbreak exemplifies these strategies are not as well equipped to protect patients from unexpected pathogenic sources of infection. Overall, the recent and well-documented occurrence of airborne infection transmission inside the OR via HCUs despite current standards of safety highlights the need for a deeper understanding of how all parts of the OR interact and what methods can be employed to create a safer surgical environment.

There are diverse sources of infections in the OR, such as surgical tools, medical equipment, surgical staff, patients, and the built environment. A substantial body of literature investigates human and procedural sources of contamination and infection, which has led to interventions that aim to mitigate these risks. It has been well-established that, for example, use of personal protective equipment (PPE) by surgical staff and administration of antibiotic prophylaxis to patients both reduce the risk of SSIs.^14–16^ In 1999, the CDC’s “Guideline for Prevention of Surgical Site Infection” outlined the importance of proper PPE donning by surgical staff, antimicrobial prophylaxis, proper ventilation, and surface disinfection, among other interventions.^16^ While some guidelines are provided with respect to OR ventilation, such as maintaining positive pressure and conducting a minimum of 15 air changes per hour, little emphasis is given to how physical aspects of the OR can influence SSIs. These guidelines were updated in 2017 with a focus on the use of antimicrobial prophylaxis and other procedure-based approaches, without any discussion on how the OR built environment is involved in infection.^17^ In addition to not being considered in infection prevention guidelines, the OR built environment is generally understudied as a potential infectious disease transmission interface between patients and the surgical staff. Finally, whereas SSI prevention recommendations are intended for use by surgical staff, these individuals tend to have little if any control over their respective built environments; this indicates need for a potentially different target towards developing recommendations.

The OR built environment can be described as the human-made surroundings and parts of those surroundings that provide the setting for surgeries, including the physical characteristics, climatic characteristics, and parts of the room. Given that the built environment framework should be applicable to all surgical procedures, this explicitly does not include medical equipment and devices. Operating rooms are ecosystems with specific environmental conditions and decisions as to their temperature, pressure, and other characteristics are guided by institutions such as the CDC and ASHRAE for ORs in the US.^5,16,17^ As was mentioned previously, while the 1999 version of the CDC’s infection prevention guidelines offer basic parameters for OR ventilation, this and other aspects of the built environment are not addressed in the 2017 version. During the 18 years between these guidelines, hundreds of studies have been published that investigate the OR built environment. This field of study is of high importance because the built environment is a tractable level at which to implement infection control interventions, as it does not rely on changes to human behavior or added procedures to complicated surgical workflows. In the hierarchy of controls, it can be more effective and sustainable to institute engineering controls and redesign the built environment, rather than intervening at the individual level (i.e., clinical staff).

While the body of information surrounding the OR built environment is large and continuously growing, gaps exist in compiling the literature to better understand how different components interact to create the OR ecosystem. The majority of publications in this space focus on individual components of the built environment, such as OR doors, surgical lights, and ventilation, among others. While several reviews have also been published, these mostly compile literature related to a single component rather than aiming to synthesize information on all aspects of the built environment. One notable exception is Joseph et al. 2018, in which a literature review was conducted that focused on OR design by investigating physical features of the environment.^18^ While that review encompasses all aspects of the built environment and synthesizes existing literature in the field, it is not systematic, meaning that the process of article selection may have been subject to bias or otherwise limited in scope, and the data from the peer-reviewed literature were not extracted or analyzed. Additionally, Joseph et al. 2018 did not explicitly include airflow and airborne contamination as study outcomes, rather focusing on performance and satisfaction outcomes.^18^ While the review also addresses “patient safety,” this outcome is not consistently considered across design categories, such that it is hard to compare findings across aspects of the built environment. That review also focused only on empirical studies, and therefore excluded all modeling and simulation studies that have investigated the OR built environment.

Given the current lack of a comprehensive set of modern infection prevention guidelines surrounding OR built environment design and the growing body of literature describing its individual components, a thorough systematic literature review is warranted. Additionally, the aforementioned *Mycobacterium chimera* outbreak and the COVID-19 pandemic have highlighted the need for a deeper understanding of the mechanisms behind aerosolization and transmission of airborne infection inside the OR. It is critical to understand how the OR built environment impacts airflow, air quality, and infection risk to inform future improvements in both OR design and sustainable infection control interventions. To address these gaps, this review aims to systematically investigate how the physical characteristics and components of the built environment impact airflow, infection, aerosols, particle counts, contamination, and pathogens in operating rooms.

## 2. METHODS

### 2.1 Search strategy and selection criteria

A systematic literature review was conducted in accordance with the PRISMA statement.^19^ A literature search was conducted in December 2020 using the PubMed and Web of Science Core Collection databases. Articles were restricted by year of publication such that only those published since January 1, 2010, were eligible. The search conducted in the Web of Science Core Collection included a filter by document type to limit the results to articles. For the purposes of this review, the OR built environment was defined as the human-made surroundings and parts of those surroundings that provide the setting for surgeries, including the physical characteristics, climatic characteristics, and parts of the room; this explicitly does not include medical equipment/devices, personal protective equipment (PPE), and human behavior/movement. An exception to this was made in the discussion of OR doors, however: this aspect of the built environment relies on HCW interaction in order to function (i.e., open and close), and OR door operation has implications for the positive pressure gradient often maintained in ORs, which is a part of the built environment. Based on this definition, search terms related to the OR built environment were used to design the search phrases included in Table 1.

**Table 1.**
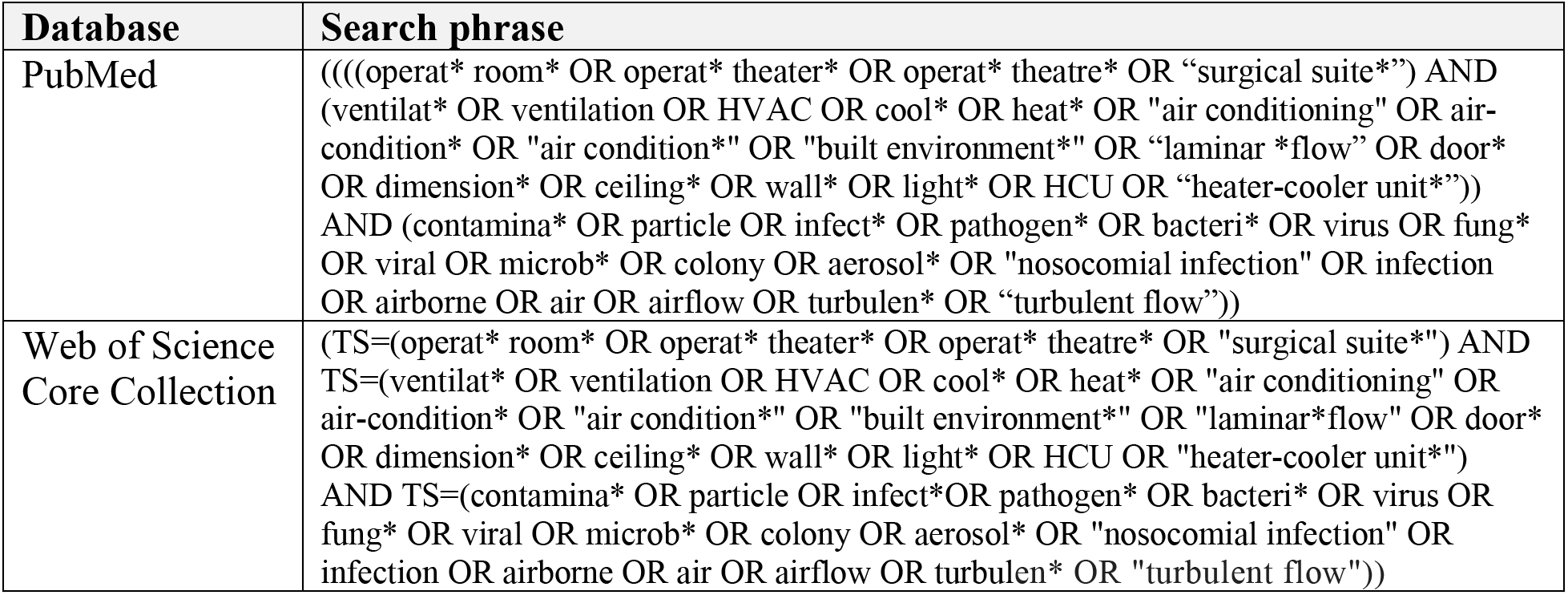
Search phrases used in the literature search conducted in December 2020.

All abstracts were screened independently by any two of thirteen individuals (SMK, ASL, OEA, AMA, MBF, TJG, SK, KRL, JAR, SHO, SLP, JY, and AJY) and discrepancies were resolved by SMK and ASL through discussion and consensus. English-language studies of any design that investigated the OR built environment in relation to airflow, contamination, and infection were eligible for inclusion. During the full text review, articles were excluded according to the hierarchy of criteria listed in Table 2.

**Table 2.** Exclusion criteria applied during the systematic literature review.

| Hierarchy | Exclusion criteria |
| --- | --- |
| 1 | Full text not available |
| 2 | The article did not take place in nor aim to study an operating room |
| 3 | The article did not compare and/or contrast aspects of the built environment |
| 4 | The article did not include airflow, contamination, or infection as a measured outcome |
| 5 | The article did not report any primary data (i.e., literature review, meta-analysis, textbook chapter, etc.) |

### 2.2 Data abstraction

Data were abstracted using a standardized survey developed using Qualtrics software (Qualtrics, Provo, UT).^20^ This survey included questions related to study design, location, sample size, OR characteristics (i.e., size, age, specialty, etc.), aspect(s) of the built environment, outcome measured, study results, and overall conclusions/recommendations. The articles were grouped by study design to further characterize current research approaches and identify gaps in how the built environment is studied and assessed. Articles were grouped into one of the following study designs: empirical, computational fluid dynamics (CFD) modeling, modeling, or simulation. For the purposes of this review, studies that took place in real ORs but only collected data during mock surgical procedures were classified as simulations.

## 3. RESULTS AND DISCUSSION

### 3.1 Article selection process

After duplicates were removed, a total of 2,965 articles were identified in the literature search. After reviewing abstracts, 2,588 articles were excluded. After eight additional duplicates were identified and removed, 231 articles were excluded in the full-text review, for a final total of 138 articles (Figure 1).

**Figure 1.**
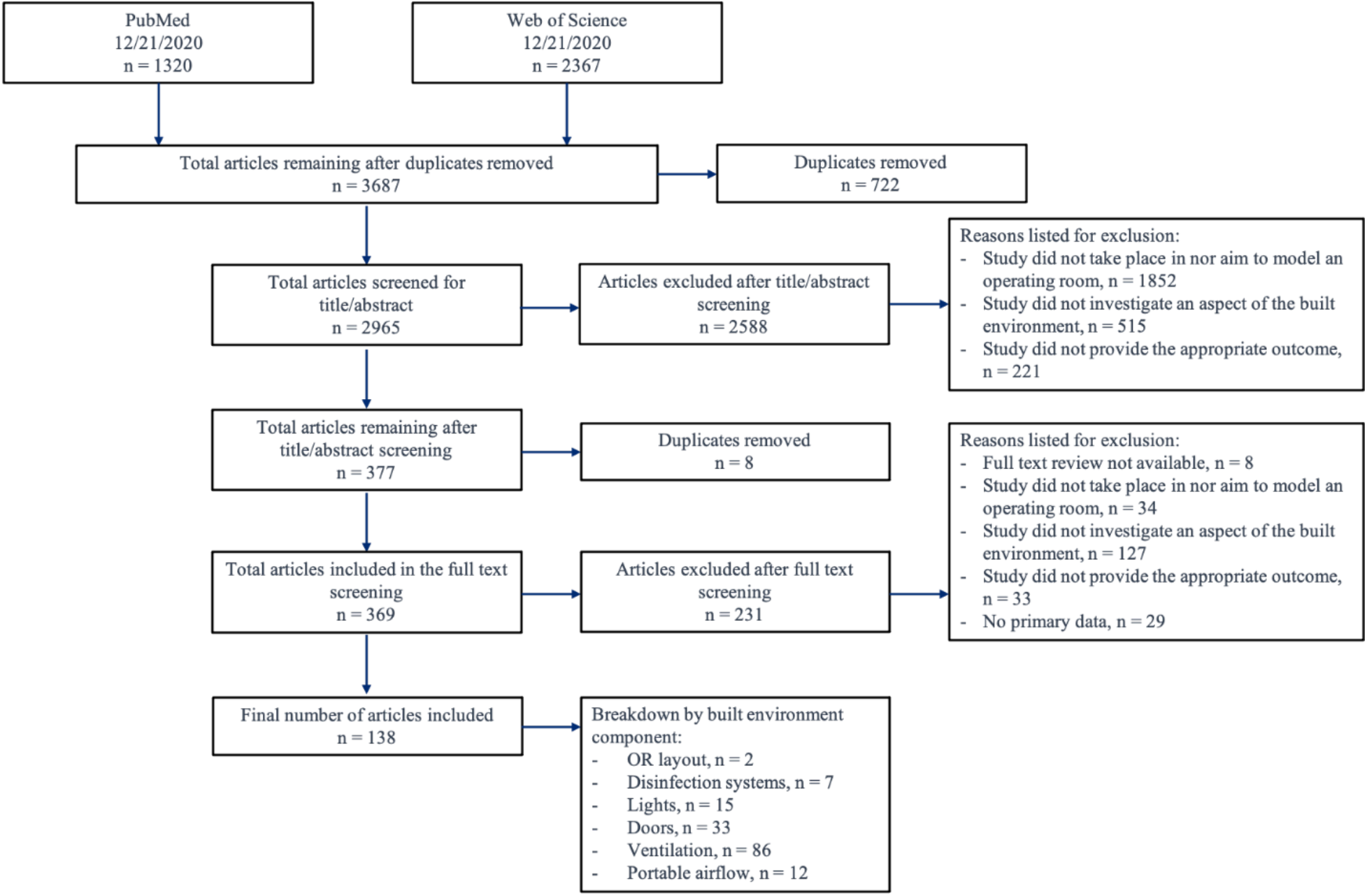
Article selection process where n = number of articles. The “breakdown by built environment component” section exceeds 138 articles included because certain articles investigated multiple aspects of the built environment.

### 3.2 Findings

The results of this systematic literature review are discussed below according to the following built environment categories: OR layout, disinfection systems, OR lights, OR doors, OR ventilation systems, and portable airflow devices. While the following sections provide an overview of the individual findings of each study, more detailed information can be found in the tables included in Supplemental Information.

The country in which each study was conducted was recorded and is included in Supplemental Information. Additionally, the survey used for data abstraction aimed to collect information on general OR characteristics, such as age and size. While a discussion of these findings was not included in the text, the Supplemental Information features graphs that aim to illustrate the distribution in area and volume of the ORs described in the studies collected in this review. While the survey allowed data to be abstracted from published literature reviews and meta-analyses, these were ultimately not included in the systematic review at hand. Despite this, these were collected and briefly summarized in the Supplemental Information, as they provide essential information on the current state of the literature surrounding the OR built environment.

#### 3.2.1 Operating room layout

##### Ceilings

None of the studies related to OR layout considered the impact of ceiling design on airflow or particle counts within the OR. Ceiling dimensions were listed, but the main focus of the articles was on other aspects of the built environment. Laminar flow units and other aspects of the HVAC system installed on or near the ceiling were the primary sources that had an effect on airflow and particle counts within the OR. These aspects of the built environment are discussed in other sections of the literature review.

No studies were identified that focused on the dimensions of the ceiling and the potential effects that it could have on airflow or particle count. This is a notable research gap. Determining the optimal dimensions of an OR can allow for greater control of the airflow going in and out of the OR, and can also affect how airflow can change at different heights. Testing has been done on airflow at different heights, but with no investigation of how the ceiling height or dimensions affect airflow. Tests could be done to determine if there are any differences in airflow with different ceiling heights. While these tests can assess particle counts, the primary focus for this part of the built environment would be ceiling dimension.

##### Floors

Only two studies were found that investigated OR floors, one conducted in Italy and one in the Netherlands, and both of these measured subsequent infection rates. In one study, the floor sections of four different ORs in an eye hospital were demarcated with colored tape to better indicate the region below the laminar flow hood, which measured 162 cm x 224 cm.^21^ The incidence of ophthalmic infections (endophthalmitis) decreased after the floor markings were implemented, although this was not a statistically significant result. In another study, 35 ORs within 30 hospitals were examined based on the complexity of their floorplan.^22^ In this study, a simple floor layout was defined as having more symmetry while a complex floor layout was characterized by having a larger presence of beams, pillars, and cavities at various spots. The median total viable count (TVC), which was the chosen metric to assess microbial air quality, among more geometrically complex ORs was 87% higher than in ORs with a simple layout, although this did not reach a level of statistical significance. These studies are described in more detail in the Supplemental Information. While these studies did not identify a statistically significant impact of OR floor layout on infection rates, they indicated that such modifications may have indirect downstream effects on OR personnel performance during surgical procedures. Nevertheless, there is a need for performing more rigorous studies that would better inform the overall understanding of the link between OR floor arrangements and infection rates.

#### 3.2.2 Disinfection systems

Seven articles were identified that discussed the implementation of disinfection systems in the OR with the goal of reducing bacterial contamination and subsequent healthcare-associated infection/surgical site infection (HAI/SSI) rates. Six articles were empirical studies and one was a simulation study; five articles were published in the USA, one in China, and one in Germany. Generally, disinfection systems refer to any system that functions to eliminate or reduce environmental contamination in the air and on surfaces. While researchers are pursuing different mechanisms for disinfection systems, current research predominantly focused on ultraviolet light (UVL) sources that illuminate OR surfaces for the purpose of eliminating or reducing colony forming units (CFU) associated with infection-causing microorganisms.

Two empirical studies investigated the use of portable crystalline ultraviolet C (C-UVC) filtered units. One such study found that, in a positive-pressured OR with simulated OR traffic, total particle counts (TPC) sized 0.5-10 μm were approximately 70% lower when the C-UVC units were used compared to when they were not used.^23^ Similarly, another study found that C-UVC reduced TPC by approximately 45% and viable particle counts (VPC) by roughly 32% when compared to the control OR.^24^ Additionally, this study found overall bacterial contamination to be lower in the C-UVC group when compared to the control (10.9 CFU/m^3^ versus 13.7 CFU/m^3^, respectively).

Only one study investigated the use of a portable pulsed xenon ultraviolet disinfection (PPX-UVD) system. This study found that the use of a PPX-UVD system reduced the presence of bacteria in the OR.^25^ Bacterial growth was observed in 17% fewer touch and settle agar plates when terminal cleaning was conducted with the PPX-UVD system as compared to traditional terminal cleaning operations. Additionally, the mean colony count per sample was reduced from 2.8 CFU to 1.6 CFU after PPX-UVD use.

Three additional studies explored the use of ultraviolet light (UVL) as a disinfection system in the OR.^26,27^ One such study, which specifically looked at UV lamps with a minimum intensity of 70 μW/cm^2^, found that significantly fewer SSIs occurred following neurosurgical procedures in the UV-equipped OR as opposed to the control.^27^ It is important to note however that this intervention was part of a greater infection control strategy being studied, and the individual impact of isolated variables was not addressed by the authors. Given this limitation, further research into the 70 μW/cm^2^ threshold is warranted. Another study found that more post-operative infections occurred in ORs equipped with standard HEPA filtration systems than in those equipped with an ultraviolet-C (UV-C) air decontamination unit.^26^ Despite these promising results, the use of UVL as a disinfection method in occupied ORs is controversial due to the potential health threats of prolonged exposure, including burns, eye damage, and increased risk of cancer.^28^ Before these disinfection systems can be studied more thoroughly in real ORs, further study on the effect of prolonged UVL exposure is warranted. Meanwhile, the use of systems employing UVL can be studied as a terminal cleaning procedure for unoccupied ORs, as suggested by an aforementioned study.^25^

In addition to UV systems, researchers have identified several other systems with potential decontamination applications for ORs. One simulation study on the application of cold atmospheric-pressure plasma (CAP) modules combined with ionic wind in an OR found a 31-89% reduction in the number of CFUs for *Escherichia coli* K12 DSM 11250/NCTC 10538 and the multi-drug resistant strains *E. coli* 21181 and 21182, with *E. coli* K12 being the strain most susceptible to inactivation by CAP.^29^ There is also interest in the use of visible-light continuous environmental disinfection (CED) systems as a method of combatting microbial surface contamination and SSIs. Following the installation of a visible-light CED system, researchers observed an 81% reduction in total CFU in the OR in which the system was installed and a 49% reduction in the contiguous OR.^30^ Furthermore, SSI rates were monitored for one year prior to and after visible-light CED system installation. SSI rates decreased from 1.4% to 0.4% in the OR in which the system was installed and from 1.2% to 0.3% in the contiguous OR. This study is especially interesting because it is the only one that explored changes in contamination in an OR adjacent to that in which the intervention was conducted.

The impact of disinfection systems on environmental bioburden has been investigated, and it has been demonstrated that these devices can reduce the concentration of viable bacteria in the OR by anywhere from 17% to 89%, depending on factors including the type of system in use, the bacterial species and strain, and the method of measurement. The subsequent effect of these systems on SSI rates has not been studied as thoroughly, however, and thus requires further analysis. The few studies that analyzed SSI rates suggest that implementation of disinfection devices may result in 1 to 3% decreases in SSI or postoperative infection rates,^26,27,30^ but strong evidence for the effectiveness against specific viruses and multi-drug resistant organisms (MDROs) is generally lacking and more research should be done. Additionally, further investigation into the efficacy and occupational safety of non-UV methods such as CAP and visible-light CED systems is warranted.

Overall, researchers have demonstrated that the use of UVL and other disinfection systems are promising methods for reducing biological contamination within the OR. However, due to potentially harmful exposure to OR staff, UV cleaning is not readily recommended by all researchers and instead is proposed as a method for terminal cleaning of empty ORs until more research has been conducted.

#### 3.2.3 Operating room lights

This systematic literature review collected 15 studies that investigated the impact of surgical lights on airflow, contamination, and infections in the OR. Of the 15 studies, three were empirical, seven were simulations, four were CFD modeling studies, and one was a non-CFD modeling study. Two of these studies were published in North America (both in the United States), 10 in Europe, and three in Asia.

The impact of overhead and surgical lights on OR air quality, airflow, and pathogens is relatively understudied as compared to other facets of the built environment. This literature search yielded 15 articles on the impact of OR lights on airflow, air quality, and infection risk. The articles examined diverse outcomes related to OR lights such as temperature, particle counts, air velocity, wound contamination, laminar airflow disruption by the lights, thermal plumes, infection risk, and bacterial and fungal contamination. Lights are a critical component of the built environment as they generate heat, which can interfere with airflow and affect aerosol content. Specific characteristics of OR lights such as their placement, shape, and type also affect airflow and potential downstream factors such as air quality and infections. Lastly, lights may also serve as a fomite, or a surface source of pathogen contamination in the OR that may facilitate infection transmission. Ten articles (67%) evaluated the effect of the placement and configuration of lights in the OR on airflow and infection related outcomes.^31–39^ Additionally, many articles studied more than one aspect of surgical lights; six articles examined the shape and type of lights,^36,40–44^ three articles observed the impacts of light heat generation,^31,32,41^ and two articles studied the lights with respect to infections and contamination.^36,45^ Modeling and simulation studies were leveraged to study multiple lighting scenarios and the empirical studies on lights utilized data from ORs to better understand real-world implications of lights on airflow and infection. The OR lights were often not the focus of the studies, but an added element without full evaluation.

The main outcomes investigated in the light studies were related to disruptions in airflow and more specifically laminar flow (LAF). Surgical and overhead lights can physically interfere with airflow, decrease air velocity, and direct airflow depending upon configuration, shape, and size.^35,38^ Turbulence from light positioning or heat generation may also impact the sterile surgical field, although the temperature of the light might be more impactful than its placement.^31,32^ The decreased air velocity distribution may also increase the exposure of the patient and healthcare workers to various indoor airborne pollutants and pathogens during the period of the surgery.^32,39^ Studies emphasized that it is important to consider airflow type and direction to better understand interactions with the lights. One CFD modeling study compared a novel LED surgical lamp design to the typical lamp type, although what defines a “typical” surgical lamp was not described in detail. The study found that the LED surgical lighting system caused fewer turbulent airflow eddies to form near the OR table. The authors equated this reduction in swirling airflow and reversed air currents with a more sterile environment.^44^ There was very limited literature on the impact of light characteristics on infections and contamination. One article demonstrated that bacteria and fungi could be isolated from OR lights and this may have been caused by cross-contamination from cleaning procedures.^45^ Another study aimed to describe the relationship between light placement, disruptions in LAF, and downstream SSIs.^36^ These studies did not provide strong evidence, due to small sample sizes and limited pathogen assessment, to support the impact of lights on HAI risk in the operating room.

Particular light characteristics and positions were associated with decreased airflow interference; however, the literature did not provide a consensus on specific optimal lighting selections. The overarching recommendation from the articles was that lighting selection is a critical component of OR design and airflow dynamics. Generally, it is recommended to select light shapes that minimize turbulence such as rounded lights as compared to square with the same surface area. Light configurations should also be selected to minimize impact on LAF and lights should be placed far from the operating field. It may also be helpful to select light placement such that human movement or interaction is discouraged in order to reduce potential airflow interference changes or contamination. CFD modeling studies have indicated that LED lights or other lower heat-generating light technologies may interfere less with airflow and particle dynamics, warranting further study in real-world ORs. It is also important to robustly disinfect lights and to prevent cross-contamination between surfaces in the OR during cleaning procedures.

The limited research on lights has highlighted particular research gaps that may be critical to explore further. The thermal impact of various lights and light geometries on airflow and potential pathogen transmission should be examined further. More research is needed on surgical lights and biological outcomes such as respiratory infection transmission, SSIs, pathogen viability, and aerosol contamination. Further research and consensus are needed on optimal light shapes and configurations. Lastly, research is needed on the relative impact of lights on airflow as compared to other factors to better understand what components of the built environment are most critical to re-engineer to ensure optimal airflow conditions to support OR infection control.

#### 3.2.4 Operating room doors

A total of 33 studies investigated the impact of doors on airflow, contamination, and infection within the operating room. These included 22 studies based on empirical data, four CFD modeling studies, two non-CFD modeling studies, and five simulation studies. Of the papers that specified an OR specialty, most were orthopedic, and surgery types that were specifically mentioned usually involved total joint arthroplasty (TJA). Twelve studies were based in North America (all in the USA), 13 in Europe, five in Asia, and three in Africa. While the overall design and findings of these studies are discussed below, more detailed information can be found in the Supplemental Information.

Twenty-six articles evaluated the frequency of door openings, but only eight articles reported the number of times doors were opened. Most articles provided the average frequency count or reported a range for the number of door openings. In general, all but one of the 26 studies agreed that there was a positive correlation between the number of door openings and biological contamination measured in colony-forming units. Only two studies concluded that door openings did not have a significant impact on microbial load, regardless of the location of in-room sampling.^7,46^ Many of these studies included general statements about higher CFU/m^3^ counts following door openings, but very few included quantitative measurements.

Although most of the studies collected in this review agreed that there is a positive correlation between frequency of door opening and bacterial contamination in the OR, the experimental methods used to measure contamination differed. One Swedish study concluded that a single door cycle increased the bacteria-carrying particles (BCPs) in the OR by approximately 2.1 CFU/m^3,47^ while another concluded that every door opening lead to a relative increase in bacterial contamination of 3% if the OR used displacement ventilation, but caused no significant change under an LAF system.^48^ Similarly, a US study found that increased door openings were statistically associated with higher CFU counts at sampling locations outside of the LAF boundaries, but not within them.^49^ Furthermore, an empirical study from the Netherlands concluded that every door opening led to a 5% increase in the odds of reaching greater than 20 CFU/m^3^ at the perimeter of the clean zone,^50^ which is the standard set by the Hospital Infection Society Working Party on Infection Control in Operating Theatres (HISWPICOT).^4^ The Healthcare Infection Society (HIS) is a charity registered in England and Wales consisting of a network of experts that focuses on minimizing HAIs worldwide by developing best practice guidelines.^51^ Finally, another empirical study concluded that the CFU count on Replicate Organism Detection and Counting (RODAC) plates placed in the OR increased by approximately 70% with a single door opening event.^52^

Three studies evaluated door openings in terms of open versus closed rather than frequency of door openings.^27,53,54^ All three studies found an increased level of contamination within the OR when the door was open as opposed to closed, regardless of whether particulate matter or CFUs were measured.

Another article varied door opening time and width as part of its study design, and found that longer door openings led to a significant increase in number of CFU counts on both bacterial and combined microbial settle plates (bacteria and fungi combined).^55^ This same study compared the width of the door opening at 45 versus 90 degrees and found that wider door openings lead to higher total CFU on combined settle plates. Two articles reported door size in addition to their results, but neither of these evaluated the impact of OR door size on particle counts or another relevant outcome.^56,57^ Similarly, seven studies documented the type of doors in the study OR (sliding versus hinged) but did not directly assess the influence of door type on any outcome of interest.

Surprisingly, only four studies measured SSIs as an outcome. One such study took place in an OR with two doors, ultimately comparing the effects of opening the internal door (which connects to an instrument preparation room) versus the external door (which connects to the hallway) on resultant SSI risk.^58^ Surprisingly, there was increased risk for SSIs that was driven by the internal door opening, while the external door opening had no association with SSI risk. The authors proposed several possible explanations for this, the most enlightening of which was the discrepancy in differential pressure across the two doors. While the pressure inside the OR was 0.9 Pa higher than that of the hallway, the pressures of the OR and instrument preparation room were equal.^58^ While this is not the only possible explanation, it suggests that differential pressure may play a significant role in the transport of contamination air into the OR. Another study found a higher mean number of door openings in the five surgical cases that developed post-operative SSIs versus the 41 cases that did not develop an SSI, although this difference was not statistically significant.^59^ One study concluded that the occurrence of more than 100 door openings during an abdominal operation significantly increased a patient’s risk of developing an SSI.^60^ Lastly, one study found significantly fewer incidents of SSIs in cases where the OR remained closed during the entirety of the procedure, meaning that no door openings were allowed, as opposed to the control cases, in which OR traffic was unregulated.^27^ However, it is important to note that this intervention was part of a greater infection control strategy being studied, and the individual impact of isolated variables was not addressed by the authors. Nevertheless, this study is notable because it suggests that such an intervention is possible, and that future research should further explore the advantages and disadvantages of completely closing ORs during surgical procedures.

Four articles agreed that there was a significant pressure decrease in the OR when doors were opened.^56,61–63^ One study found that an 11-second door opening event with the entrance of a single person through a sliding door caused the pressure differential between OR and corridor to drop from 32.6 Pa to 1.2 Pa.^56^ Similarly, a CFD modeling study found that the pressure differential between the OR and the adjacent hallway dropped from 17 Pa to 4 Pa within three seconds of the sliding door opening.^63^ This study found that the pressure change over time was almost identical regardless of whether no people, one person, or two people entered the OR when the sliding door opened. While this study did not offer information on time to repressurization, a different study found that it takes 14.11-14.93 seconds to recover from depressurization after a door opening event, depending on which door was opened and whether equipment or personnel passed through it.^61^ While these studies focused on pressure changes during and immediately following a door opening event, another study demonstrated that door openings affect the minimum pressure recorded in the OR but not the average room pressure, suggesting that positive pressure is easy to recover after a door opening.^62^ However, this same study found that for 77 of the 191 procedures studied the door was open long enough for positive pressure in the OR to be defeated, meaning that the pressure of the OR was allowed to equilibrate with that of the hallway, thus suggesting that contaminated air may have entered. While the aforementioned studies agree that door openings lead to a pressure decrease in the OR, they did not specify that the positive pressure gradient was completely defeated. Given this observation, more research aimed at determining whether door openings indeed defeat the positive pressure in ORs is warranted. More specific information as to whether any single door opening event can defeat positive pressure and the circumstances that lead to this would help outline what interventions are needed to prevent the infiltration of contaminated air during door openings.

A total of 12 out of the 33 articles that discussed OR doors as they relate to airflow and contamination did not include recommendations following the results of their studies.^27,46,50,53–57,62,63,64,65^ Of the 21 articles remaining, five proposed recommendations that focused on altering specific behaviors of the OR staff.^7,66–69^ These recommendations included limiting unnecessary staff movement related to OR door openings, limiting the number and duration of door openings, and limiting the number of staff in the OR in general. Seven articles included recommendations pertaining to the improvement of education, awareness, training, and organizational culture.^52,59–61,67,70,71^ These included increasing knowledge and awareness relating to OR door openings and their relation to SSIs, and education and training in general for improving current practices. Two papers recommended actively discouraging unnecessary social visits and nonessential OR entries.^60,61^ Seven articles recommended further adjustments and maintenance of existing environmental components.^7,47–49,61,72,73^ These included improving and maintaining OR ventilation, continuous maintenance and risk assessments of LAF and other technical systems, increasing the frequency of air exchanges when more surgical personnel are present, decreasing the exhaust rate while the OR door is open, making use of intercoms and other communication technologies, limiting usage to a single OR door before and during surgery, and ensuring the temperature difference between inside and outside of the OR is minimal. Two studies recommended implementing new environmental components, such as a non-visible door counter, door locks, and a C-UVC unit placed closer to potential sources of contamination.^23,52^ Five articles recommended organizational and logistical improvements, including robust auditing processes, improving coordination in the instrument prep room, ensuring that all required surgery equipment is inside the OR at the time of incision, and including a central logistics distribution area.^52,58–60,71^ Only one article recommended designing and adapting ORs to compensate for the high number of door opening events of both short and long duration, including utilizing an ultraclean lobby, minimizing the number of doors in an OR, and optimizing the geometry of the OR and the air control system.^74^ Similarly, only one article specifically recommended integrating ventilation and door operation in order to create a compensatory response to door opening events.^47^ This study demonstrated that reducing the OR exhaust rate during door opening events helped minimize the entrance of contaminated air through the door, therefore reducing the impact of such events on subsequent infection risk.

Overall, most recommendations provided by the literature surrounding OR doors involved changes to human behavior such as implementing additional training to discourage unnecessary door openings. In contrast, very few studies addressed the possibility of re-engineering ORs and their ventilation systems in order to better accommodate necessary door opening events. While a few papers did make such recommendations, there remains a large gap in studying how changes to the OR built environment can reduce the need for or impact of door openings. Future research on engineering OR layouts, instrument prep rooms, corridors, etc. could be more useful in providing solutions and recommendations specific to changing the environment.

#### 3.2.5 Operating room ventilation

In total, this review identified 86 articles that investigated the relationship between ventilation systems and the operating room environment. Of these articles, 46 are empirical studies, ten are simulation studies, 27 are CFD modeling studies, and three are non-CFD modeling studies. Thirty-nine studies were conducted in Europe, 24 in Asia, 11 in North America (all in the USA), three in Africa, three in South America (all in Brazil), four in Eurasia (Turkey), and two in Oceania (both in New Zealand). Of the 86 articles, four could not be linked to a specific country or region. The USA produced the most publications, followed by Italy and Sweden.

##### Empirical studies

Overall, 46 empirical studies of OR ventilation were captured in this systematic review.^7,22,33,46,48,52–54,66,70,75–84,85–94,95–104,105–110^ Twenty-four of these studies investigated LAF with 22 comparing LAF to another type of ventilation system^33,48,54,75–81,82–91,92,93^ and two investigating the configuration and system settings of LAF canopies.^52,104^ One of the later articles sought to compare the performance of a newly installed LAF system with a LAF system that was overdue for an air filter change. In the latter LAF system, performance was measured both before and after filter replacement.^104^ It was found that replacing the old filter substantially increased the air renewal rate and decreased the particle concentration by 97%, such that these measurements were similar to those in the OR with the newly installed LAF system. However, while the average viable particle concentration decreased by over 50% following the filter change, it still remained significantly higher than that of the new system.^104^ A second article investigating LAF configuration compared bacterial contamination sampled from inside versus outside the LAF area in an OR.^52^ Ultimately, the findings showed that samples taken inside the LAF zone were consistently less contaminated than those taken outside, and this difference was statistically significant. Furthermore, of the specific organisms investigated in this study (namely *Staphylococcus aureus*, *Bacillus* sp., *Micrococcus* sp., diphtheroids, mold, and gram-negative rods), all were found to have similar distributions among the plates, regardless of sample collection location relative to the LAF canopy; this finding suggests that LAF’s ability to decrease bacterial contamination is consistent across various pathogens.^52^ These were the only empirical studies collected that investigated LAF configuration without comparing it to another ventilation system.

The 22 articles that compared LAF to another type of ventilation system, often considered conventional or mixing systems, but many failed to specify the details of the HVAC system to which LAF was being compared. Given this shortcoming, it is perhaps more meaningful to analyze these studies in terms of the outcomes being reported. Of the 22 studies, ten investigated SSI rates, nine investigated bacterial contamination, two investigated particle loads, and one investigated aspects of airflow in the operating room.

Of the ten studies that explored SSI rates, five failed to find a statistically significant difference between LAF and another ventilation system,^83,84,86,89,90^ two found higher incidence rates of SSIs under LAF conditions,^85,93^ and three found that surgeries performed in LAF-equipped operating theaters have lowered risk or rate of SSIs.^87,91,92^ Interestingly, seven of the ten studies investigated the outcomes of general orthopedic surgeries,^83,85,86,89,90,92,93^ while the other three studies investigated orthopedic trauma surgeries,^84^ gastric surgeries,^87^ and vascular surgeries.^91^ The two studies that found a positive correlation between the use of LAF and SSIs were both restricted to total knee and total hip arthroplasties (TKAs; THAs), and both defined the outcome of the study as the number of patients requiring a revision surgery due to the development of a deep infection. One such study included a total of 88,311 procedures, 35.5% of which were conducted under LAF conditions.^85^ This study found that, for hip replacement surgery, the percentage of patients requiring revision surgery was 0.148% under LAF and 0.061% under conventional ventilation (CV); similarly, for knee replacement surgeries, 0.193% of patients under LAF required a revision surgery versus 0.100% under CV conditions. The second study included 91,585 THAs, of which 38.6% were performed under LAF conditions. This study found that the incidence of revision surgery was 0.20% under LAF versus 0.11% under CV at 6 months post-surgery, and 0.25% under LAF versus 0.17% under CV at 12 months following the original procedure.^93^ Although the sample sizes in these two studies were unusually large, it is important to note that one of the five studies that found no effect of LAF systems on SSI rates reported a much greater sample size of 803,065 surgeries.^84^ This study focused on orthopedic trauma surgeries and measured the incidence rate of SSIs within 90 days post-surgery, but did not find a statistically significant difference between the use of LAF and plenum ventilation systems. Additionally, one of the studies which found that LAF conditions reduced SSI rates investigated 51,292 THAs, ultimately finding that surgeries performed in ORs equipped with a high-volume vertical LAF system had a lower risk of revision surgery due to infection incidence.^92^ These examples serve to illustrate the fact that conflicting results about the use of LAF have been found, even by studies of similar design and magnitude.

Of the 22 articles comparing LAF with an alternative ventilation system, nine articles focused on differences in bacterial contamination (i.e., air and/or surface) as the primary outcome.^48,75–82^ All nine studies found that bacterial contamination was significantly lower in operating rooms equipped with LAF systems than those equipped with other ventilation systems. The largest of these studies included data from 71,655 surgical procedures conducted across 15 operating rooms over the span on 12 years.^80^ This study found that the use of an LAF system decreased the mean microbial air contamination from 40.02 UFC/m^3^ to 1.46 UFC/m^3^, a statistically significant result. The authors of this study did not specify the definition of “UFC” beyond indicating that it was a unit for measuring microbial contamination; it may be the result of a translation issue, and refer to the more common term “colony-forming unit” or “CFU.” In addition to monitoring microbial air contamination, the study found that LAF decreased the mean airborne particle concentration from 189.96 particles/m^3^ to 3.92 particles/m^3^, which was also statistically significant.^80^ One of the nine studies collected samples in the morning and in the evening under both types of ventilation.^78^ A statistically significant decrease in bacterial contamination in LAF ORs was observed when considering morning samples only. No statistically significant differences were observed between the samples taken in the evening, regardless of the ventilation system. Overall, there is a general agreement across the articles collected in this review that ORs equipped with LAF ventilation systems have lower levels of microbial contamination than ORs with other ventilation systems such as conventional ventilation (CV) and turbulent ventilation (TV).

Two of the 22 empirical studies investigating the use of LAF over other ventilation systems focused on particle sizes and concentration as the main outcomes.^54,88^ One study found that CO_2_ levels and particulate matter of all sizes were statistically significantly lower in the ORs equipped with LAF versus those with natural ventilation.^54^ A definition for “natural ventilation” was not provided in this study. The second study found that particle loads were significantly reduced with the use of LAF ventilation regardless of sampling location, even outside of the LAF boundary, and this was observed throughout the entire timespan of the surgery.^88^

The last of the 22 papers explored various characteristics of the airflow inside the OR as the outcome.^33^ This study found that the velocity of airflow directly above the patient was significantly lower under LAF conditions than under mixing ventilation conditions. The authors attributed this difference to the existence of equipment or other objects that disrupted the LAF ventilation but not the mixing ventilation, leading to a failure in achieving the minimum standard velocity requirements.

As discussed above, while there is general agreement in the literature that LAF reduces airborne contamination in the OR, this does not translate to widespread agreement on the relationship between LAF use and SSIs. Despite this discrepancy, the overall recommendations provided by studies on the use of LAF do not vary greatly. Regardless of the finding, the vast majority of these studies conclude that more research about the use of LAF must be conducted before any concrete recommendations are made. The few studies that did provide a recommendation generally suggest that hospitals leave their ORs in their current state: if LAF is currently installed, it should not be removed without further research, and if it is not installed, it should not be installed without further research. Additionally, since few studies provided a recommendation regarding the use of LAF systems, specific suggestions regarding LAF settings such as diffuser size and airflow velocity are lacking in the literature based on empirical data.

Of the 46 studies on OR ventilation that were based on empirical data, 22 did not investigate LAF.^7,22,46,53,66,70,94–97,98–103,105–108,109,110^ It is important to note that the conflicting way in which authors use the term “unidirectional airflow” may have led to an underestimate of the number of papers that investigated LAF. Some authors equate unidirectional airflow with LAF and use these terms interchangeably, while others do not; some studies explicitly differentiate between the two. In the previous section discussing LAF, a conservative approach was taken and only studies in which these two types of airflow were specifically equated were discussed. Studies that addressed unidirectional airflow without any reference to LAF are included in the group to be discussed below.

Of the 22 non-LAF empirical studies, eleven articles compared two or more ventilation systems, most of which involved a comparison between turbulent or mixed ventilation and a unidirectional airflow system.^22,70,99,100,102,103,106–109,110^ Eight articles included microbial contamination in the operating room as one of the recorded outcomes, although the specific ventilation systems under study varied between articles.^22,70,99,106–110^ Similar to the literature on LAF, there was a general consensus that microbial contamination is lower in operating rooms equipped with unidirectional airflow as opposed to turbulent and mixed airflow systems, regardless of how this was measured (i.e., active versus passive sampling methods). Those studies that compared mixed and turbulent airflow systems found that the microbial contamination is often very similar, and any differences tend not to be statistically significant. One study stood out due to its investigation of upward displacement ventilation (UWD), which was compared to downward unidirectional airflow (UDF) combined with mixing ventilation.^108^ This study ultimately found that the microbial contamination in operating rooms equipped with UWD averaged around 27 CFU/m^3^, as opposed to the average 1 CFU/m^3^ found in UDF ORs; whether this result was statistically significant was not specified. Two of the 11 articles only reported particle concentrations as the outcome of their study.^100,102^ One of these articles was unusual due to the unique HVAC systems studied, namely rotary desiccant air conditioning (RDAC) and liquid desiccant air conditioning (LDAC), neither of which were mentioned in any other study captured by this review.^100^ This study found the lowest particulate matter concentration in RDAC-equipped operating rooms, regardless of particle size. However, only ear, nose, and throat (ENT) procedures were performed in RDAC ORs, whereas trauma surgeries were performed in LDAC and conventional HVAC ORs, so the difference in PM concentration cannot be solely attributed to the type of ventilation. Another study warranting special mention compared upward displacement airflow (UWD) and unidirectional downward airflow (UDV), with healthcare worker exposure to the ultra-fine particles (UFP) in surgical smoke as the outcome. This study found that HCW exposure to surgical smoke was up to 13 times higher under UWD conditions than under UDV conditions.^102^ Finally, only one article measured the incidence of SSIs as an outcome, and failed to find a statistically significant difference in SSI rates between unidirectional, turbulent, and mixed airflow ventilation systems.^103^

As was the case with the studies focusing on LAF, concrete recommendations about the use of any one specific ventilation system are lacking from the literature, as are specific suggestions regarding ventilation settings. Furthermore, the few studies that do provide recommendations usually suggest better air quality monitoring programs to help ensure that the ventilation system, regardless of type, is functioning properly and maintaining contamination levels below the recommended limits. This is likely due to the fact that, given the limited number of studies that measure the incidence of SSIs as the outcome, authors are hesitant to make recommendations based on changes in airborne particles and bacteria that might not directly translate to lowered infection rates.

The remaining 11 empirical studies investigated various aspects of ventilation function, such as air renewal rate, the use of HEPA filters, and air supply diffuser size, among others.^7,46,53,66,94–98,101,105^ One article reported the rate of positive cerebral-spinal fluid (CSF) cultures and postoperative central nervous system infections (PCNSIs) before and after a defective air filtration system in an operating room was replaced.^105^ The study found that, in the year before the system was replaced, the rate of positive CSF cultures was 1.9% and that of PCNSIs was 19.6%, while in the three years following the replacement, these rates decreased to 0.0% and 0.6%, respectively. Interestingly, there was disagreement among the studies that explored ambient parameters such as temperature and humidity. While one study found that neither ambient temperature nor humidity had a significant impact on microbial loads in the operating room,^46^ another found that the microbial count increased by an average of 9.4 CFU/m^3^ with each increase of 1°C in the ambient temperature of the operating room.^7^ Furthermore, one study of 18,910 patients across eight hospitals found that both higher ambient temperature and increased humidity were significant risk factors for the occurrence of superficial SSIs.^98^ Two studies exploring air renewal rates reported contrasting results of significant interest. Both studies investigated the effect of air renewal rate on airborne particulate matter and bacterial contamination, but one study was conducted in operating rooms at rest while the other in active ORs. The first study found that varying the air renewal rate between 6 and 30 air changes per hour (ACH) did not have a significant effect on either particle or bacterial contamination,^101^ while the latter found that increasing the air renewal rate from 20 to 30 ACH significantly decreased the concentration of particles of all sizes, regardless of the OR specialty.^66^ Interestingly, this study also found that the effect on bacterial contamination differed between the two specialties investigated; more specifically, the increase in air renewal rate was found to significantly decrease the bacterial concentration (CFU/m^3^) in colorectal ORs, whereas in trauma ORs only the bacterial settle rate (CFU/hr) was significantly decreased.^66^ The remaining articles that investigated ventilation function assessed ventilation quality and air supply, but did not provide any specific recommendations beyond general advice about regular HVAC maintenance and HEPA filter replacement.^53,94–97^ Overall, a large volume of empirical data describes the relationship between various aspects of ventilation in ORs and outcomes such as airborne contaminants, risk and incidence of SSIs, and airflow. Given the lack of consensus regarding empirical findings related to SSIs, however, specific recommendations for the optimal design of OR ventilation systems are limited and inconsistent.

##### Simulation studies

Of the 86 articles that investigated ventilation, ten were simulation studies, here defined as studies in which mock surgeries were conducted in a real OR setting. Four of these explored aspects related to LAF ventilation,^40,111–113^ four investigated exhaust vent/grille/diffuser layouts and air renewal rates,^114–118^ and one studied the use of HEPA filters.^119^ One article measured the microbial contamination at the patient table when the LAF system was equipped with an air curtain, a multi-diffuser array, or a single large diffuser.^112^ The level of contamination under the single large diffuser set up was 2 CFU/m^3^, whereas that of the multi-diffuser set up was 11 and 24 CFU/m^3^ for air supply velocities of 30 and 50 feet per minute respectively, and that under the air curtain condition was 87 CFU/m^3^. These results indicate that, amongst the configurations studied, a single large diffuser that covers the entire operating table is the most effective at protecting the patient from microbial contamination.^112^ A subsequent study led by the same author compared airborne contamination in ORs equipped with single large diffusers versus those with multi-diffuser arrays or the older 4-way throw diffusers.^117^ Although the later publication expressly did not study LAF, the authors reached similar conclusions, namely that airborne contamination in the OR is minimized when a single source of unidirectional, downward flow is in use (Figure 2)^120^. Two other studies investigated the influence of LAF on the pathway of contaminants leaving a patient’s mouth during normal breathing and coughing, one with a breathing simulator^113^ and another with human volunteers.^111^ Both studies found that, without the use of LAF, contaminants flowed freely towards the surgeons’ breathing area, whereas the engaged LAF system successfully directed the exhaled air away from surgeons. This is a noteworthy finding, as the vast majority of the LAF-related articles collected by this systematic literature review discussed their results in the context of patient rather than healthcare worker safety. One of the simulation studies measured the number of particles that shifted from the mannequin patient head into the surgical field in two different ORs, one built to meet International Organization for Standardization (ISO)-5 cleanliness and another built to ISO-6 standards.^116^ In this study, the ISO-5 OR was held to the standard of <3,520 particles/m^3^ greater than or equal to 0.5 μm, while ISO-6 was held to the standard <35,200 particles/m^3^ greater than or equal to 0.5 μm. Interestingly, a significantly larger number of particles shifted from the head to the surgical field in the ISO-5 OR. In discussing this, the authors concluded that these results may be explained by the difference in air-conditioner outlet layout (ACOL) between the ISO-5 and ISO-6 ORs, since the ACOL of the ISO-5 OR did not fully cover the operating table.^116^ This conclusion is in agreement with the aforementioned studies of diffuser layout, which also found that a single large diffuser that covers the patient table entirely could most effectively minimize contamination in the surgical field.^112,117^ The remainder of the simulation articles assessed exhaust layout and air supply options, but did not offer any specific recommendations beyond the incorporation of a HEPA filter in ventilation systems. Similar to the empirical studies discussed above, while some simulation studies offer specific recommendations about ventilation installation and settings, these recommendations are inconsistent across the literature.

**Figure 2.**
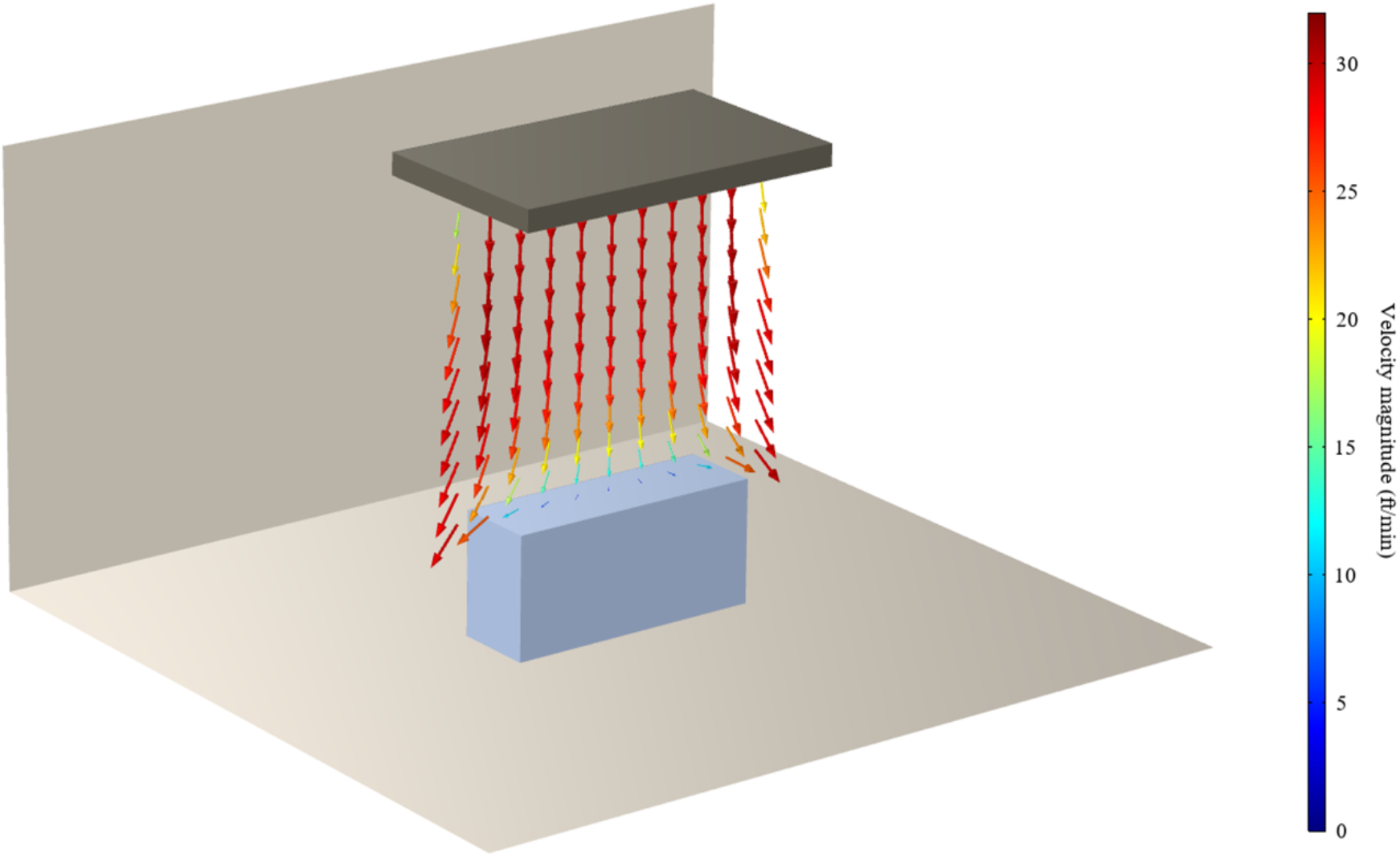
CFD model showing a single source of unidirectional, downward airflow over an operating table inside an OR. Vertical LAF is typically installed in this configuration, with downward airflow designed to cover the entire operating table. Produced with COMSOL Multiphysics software.

##### Modeling studies

In addition to studies reporting empirical data and simulations of mock surgeries, studies based on modeling of OR ventilation were captured in this systematic literature review. Of the 86 total articles investigating OR ventilation, 30 used modeling approaches to investigate the effects of HVAC components on the built environment.^31,41,64,121–127,128–137,138–147^ Twenty-seven of those 30 articles utilized computational fluid dynamics (CFD) modeling.^31,41,64,121–127,128–137,138–144^ The numerical parameters in CFD models are often validated to some degree via small-scale or simplified experimental setups, or using previously published measurements.

Eight of the 26 CFD modeling articles studied the effect of inlet air velocity on the built environment. Six of these studied inlet airflow velocity’s effects on contamination, particle count, and particle deposition, but did not come to a consensus on an optimal inlet air velocity value.^31,123,125,126,133,134^ One article concluded that contamination was minimized at a velocity of 2.37 m/s, but cautioned that airflow can easily be disrupted by slight changes in the design of the OR air curtain and exhaust vents^124^. The last article studied inlet airflow and its impact on airflow patterns.^137^ The study concluded that if temperature is held constant, inlet air velocity has very little effect on airflow patterns in the OR.

Six of the 26 CFD modeling papers studied the configuration of exhaust vents and its impact on airflow. Two studies reported that the addition of extra exhaust vents impacts airflow and reduces particles compared with the original configurations.^122,141^ One study showed that when both floor and ceiling exhaust vents are implemented, particle counts near the operating table decrease,^139^ while another study warned that four-way suppliers (ceiling, walls, and floor exhaust) could increase air velocity near the floor, which could lift settled particles off of the floor and re-introduce them into the sterile area.^140^ Another study reported that eight exhaust vents distributed between the four corners of the OR resulted in improved air distribution and minimized particle dispersion, as compared to an OR with four exhaust grilles divided between two corners.^132^ One study compared a legacy (unidirectional) HVAC system with a layout containing ceiling exhaust vents and another layout with extra ceiling diffusers outside of the sterile zone but no ceiling exhaust.^135^ It was found that both new layouts performed just as well as the legacy system at sweeping particles created by HCWs out of the sterile zone, suggesting that extra exhaust and supply placement might not be of great importance.

One of the 26 CFD modeling articles studied configurations of HEPA filters (i.e., number of filters utilized, size of filters) with and without a UCV system and their effects on bacterial contamination measured as BCP/m^3^.^41^ The study determined that the configuration which utilized HEPA filters (at least 3 m x 3 m) without the presence of an ultra-clean air system performed in the same class of cleanliness (<29 BCP/m^3^ within 0.5 m of the wound) as the configuration that utilized both HEPA filters and a UCV system, suggesting that the use of a HEPA filter was the most impactful aspect of the system.

Two of the 26 CFD modeling papers studied air renewal rate by modeling different rates and their impact on particle count. The study results were somewhat contradictory, as one study found that increasing air renewal rate decreased airborne particles (tested 15, 23, and 31 ACH)^143^ while the other study found that particulate counts were consistent for all four air renewal rates tested (5, 10, 20, 39 ACH).^128^ This discrepancy is not surprising, given that there are multiple differences such as exhaust layout in the ventilation systems modelled in each of these studies that are readily apparent.

Five of the 26 CFD modeling papers compared different ventilation schemes. Two studies comparing horizontal and vertical LAF agreed that horizontal LAF systems are better at reducing particle counts.^130,138^ Three other studies modelled different ventilation schemes, including LAF and temperature-controlled airflow (TAF), having found conflicting results.^64,127,142^ One such study found that while LAF was able to meet the 10 CFU/m^3^ threshold at two of the three sampling locations, TAF only met this standard at one location.^127^ The second study found that TAF consistently met the 10 CFU/m^3^ standard across all sampling locations at nearly every air renewal rate tested, whereas neither the horizontal nor vertical LAF systems were able to achieve such cleanliness, regardless of air renewal rate.^142^ A third study found that LAF and TAF both met the threshold at the wound and instrument table, but LAF failed to maintain the recommended cleanliness in the periphery of the room.^64^ Despite these discrepancies, the three studies tended to agree that both LAF and TAF performed better than the alternative ventilation systems, such as mixed and turbulent airflow.

Four of the 26 CFD modeling articles studied diffusers’ impact on air quality. Two studies concluded that increasing the size of an LAF diffuser reduced particles at the operating table.^121,129^ One study tuned the air velocity of an LAF diffuser which influenced the degree to which contaminants entered the sterile zone.^144^ The study determined that the lowest level of contamination was observed with the air curtain speed at 1.5 m/s. The last paper compared the use of a unidirectional horizontal diffuser alone to the use of a unidirectional diffuser in conjunction with an air curtain.^131^ The paper found the scheme with an air curtain was more successful at reducing CO_2_ concentration at HCW breathing height than the unidirectional diffuser alone.

All three non-CFD modeling studies investigated how different air renewal rates impacted contamination in the OR. Two of these studies concluded that increasing air renewal rates led to decreased contamination.^146,147^ One of these also concluded that LAF was associated with lower contamination levels than TV, with median values of 12 CFU/m^3^ and 71 CFU/m^3^ for LAF and TV, respectively. The third study found that increased air renewal rates did not consistently provide an overall cleaner environment.^145^ This study also conducted a cost-benefit analysis which concluded that a more efficient way of distributing or managing the OR air might be a more cost-effective alternative to increasing air renewal rates.

Lastly, one of the 26 CFD modeling articles studied the angle of distribution of the supply diffusers in an Angular Air Distribution (AAD) system, which delivers laminar flow over the operating table at an angle rather than vertically downwards.^136^ This study found that the new 45-degree and 60-degree designs yielded the same acceptable air velocities but caused less turbulence in the OR than the traditional vertical LAF. It is interesting to note that, while other CFD modeling studies collected by this review made an effort to validate their models through empirical studies in real or mock ORs, this study did so by conducting smoke visualization studies in an OR prototype that is about one-fourth the size of a real OR. The widespread use of such OR prototypes with accurate ventilation system layouts could provide increased flexibility in the researchers’ ability to study OR ventilation, while alleviating the need to rely on a hospital’s willingness to allow such studies to take place in their ORs.

##### Overarching observations in ventilation

Studies that investigated OR ventilation systems and their impact on airflow, contamination, and infection accounted for over half of the publications in this systematic review. Despite the wealth of data surrounding OR ventilation that is available in the literature, specific recommendations stemming from these data are somewhat limited in scope and subject to inconsistencies. The few studies that provided recommendations usually focused on vague policy-based solutions such as additional staff training, more robust surveillance practices, and SSI prevention plans that do not include built environment interventions. While multiple studies were clear in saying that their results supported one ventilation system or specific setting over another, most stopped short of specifically recommending a change to current procedures. As is described above, a common theme amongst empirical, simulation, and modeling studies is the study of LAF ventilation systems. Across all three study designs, there is general agreement that vertical LAF reduces particle counts and bacterial contamination to a greater extent than traditional mixed and turbulent ventilation, especially in the area directly under the diffuser. There is similar agreement regarding vertical unidirectional airflow when compared with mixed and turbulent ventilation. Even though better air quality in ORs is generally thought of as being beneficial, the relationship between air quality and subsequent infection rates is understudied. This perhaps explains the lack of agreement among empirical studies that investigated the impact of using LAF on surgical site infection rates. Even though there is evidence to say that LAF significantly reduces airborne contaminants, if these are not a reasonable indicator of SSI risk, then ventilation system might not be the most impactful choice in the design of an OR built environment.

Another common theme across all three study designs is the focus on patient safety. Nearly all of the studies collected by this review discuss airflow, contamination, and infection in the OR in the context of the patient rather than the surgical staff and HCWs in general. More specifically, the discussion surrounding infection in the OR is centered on SSIs, with almost no studies having discussed respiratory infection transmission in the OR. While there is general agreement in the literature that common interventions such as positive pressure and HEPA filters are in the interest of both the patient and the HCWs, more nuanced interventions such as specific ventilation systems and diffuser airflow velocity are rarely studied in the context of respiratory infection transmission between patient and surgical staff.

Overall, while there is a significant volume of data in the literature surrounding OR ventilation systems, information on outcomes related to aerosol movement and respiratory infection transmission is scarce. Future studies on this aspect of the OR built environment should investigate aerosol generation and dissemination under various ventilation systems and settings, with a special focus on aerosols reaching the patient, HCW breathing zones, and surgical sites.

#### 3.2.6 Portable airflow devices

The results of this systematic literature review yielded 12 papers that explored the use of portable airflow devices in the OR. None of these studies directly assessed the impact of portable airflow devices on SSI rates or other healthcare-associated infections.^148,149,150–157,158,159^ Instead, they measured or simulated levels of airborne contamination, such as particle counts and bacterial contamination (typically measured as CFU per volume or CFU per hour), which may serve as potential proxies for infection risk. Of the studies reviewed, 11 took place in the United States or Europe and one study was a collaborative effort between researchers in the US and China.^159^ The collection is composed of five empirical studies,^148–150,154,157^ one simulation study,^152^ and six CFD modeling studies.^151,153,155,156,158,159^ The surgical focus of the research evaluating portable airflow devices was similar to that of HVAC, with the majority of the empirical studies focused on orthopedic surgeries.^148–150,154^ Only two empirical studies considered non-orthopedic surgeries: one assessed the use of portable airflow devices during neurosurgical procedures^157^ and the other considered simulated intravitreal injections.^152^

Most of the limited research in this area is focused on mobile laminar flow (MLAF) devices, although one empirical paper evaluated the use of a mobile ultra-clean unidirectional airflow screen.^152^ Two studies used CFD modeling to explore the use of prototype devices: one proposes the use of additional jets and/or suction devices placed on either side of the wound area,^159^ and the other describes a ventilated blanket designed to direct additional airflow directly across the surgical site.^158^

Evidence for the benefits of MLAF devices is mixed. Two studies agree that device function is optimized when the air velocity is around 0.4 m/s.^151,155^ Reportedly, MLAF devices can reduce BCP sedimentation rates by up to 70%^153^ and can reduce airborne particles of any size by more than 60%.^154^ Several studies found that employing an MLAF device in the OR, in addition to an existing conventional or turbulent HVAC system, significantly reduced airborne contamination.^148,150,157^ Another study went so far as to suggest that a mobile ultra-clean UDF device alone was sufficient to limit the risk of airborne infection, without the use of the main HVAC system.^152^ However, a modeling study that evaluated the use of MLAF in conjunction with an LAF HVAC system recommended caution when using MLAF devices. The study showed that due to disruption of airflow patterns in the OR by the mobile device, the concentration of BCPs above the operating table actually increased.^156^ The authors also caution that such portable airflow devices should not be used without careful assessment of the airflow patterns in the OR. This finding is in sharp contrast to an empirical study of MLAF used to ventilate the instrument table in an LAF-equipped OR, which concluded that the addition of the MLAF device was safe.^149^

Two novel portable airflow devices are investigated using CFD models.^158,159^ This approach allows for the collection of preliminary data and results, without the inherent risks that would be involved in testing prototype devices during surgery. One of these papers describes the use of a ventilated blanket device that provides increased airflow directly over the surgical site.^158^ The authors note that such a device might be of particular benefit in emergency or resource-limited settings, where any reduction in airborne contamination might have a protective benefit during surgery. Results from the other study suggest that increased local ventilation, when designed appropriately, can reduce particle deposition near the surgical site.^159^ This finding is in agreement with the majority of work on MLAF devices, suggesting there may be some benefits to the use of portable airflow.

Relatively few research studies have focused on the use of portable airflow devices in the OR and their potential to reduce the risk of airborne contamination and therefore reduce infection risk. Existing efforts are limited by small sample sizes, and have focused almost exclusively on the use of MLAF devices in the US and Europe. Outcomes are evaluated in terms of airborne particles and bacteria. This is a common approach when evaluating many aspects of the OR built environment and airflow, but particularly notable in this case because the systematic review methods employed here did not return a single study assessing the relationship between the use of portable airflow and the risk of SSIs or other HAIs. Bridging this gap by investigating whether portable airflow devices ultimately influence SSI rates is a common recommendation in existing studies;^148,149,154^ unfortunately, it can be difficult to assess the risk of SSIs directly, due to limited sample sizes and the overall low rates of SSIs in modern operating theaters. However, the link between the relatively low levels of airborne contamination in modern ORs and subsequent infection risk is increasingly controversial, especially in light of improved patient-care practices and sterile techniques. The two novel portable airflow devices have only been tested using models, and the existing prototypes are appropriate only for proof-of-concept testing, rather than development toward commercial use. Their utility could be better understood with optimized prototypes and by collecting empirical data from the OR environment. Ultimately, larger-scale studies that control for the existing HVAC ventilation system, describe the orientation of the portable device, and evaluate infection rates are needed to assess the benefits of portable airflow devices.

## 4. CONCLUSIONS

Based on the findings of this systematic review, we have reached a set of conclusions about aspects of the OR built environment and their effect on airflow, contamination, and infection risk. Many of our findings are specific to one aspect of the built environment, considered independently of the general OR layout.

- Studies agree that it is better to keep OR doors closed as much as possible, although most research in this area focuses on modifying HCW behavior, which is outside the scope of this review; it is clear that more investigation of OR door engineering is warranted
- Lights can disrupt airflow patterns due to their physical structure/shape, especially when placed in the flow path of LAF
- Airflow disruption due to heat produced by surgical lights is actually more significant than their physical interference
- UV disinfection systems have potential to be effective in terminal cleaning of the OR, or possibly as a within-vent option, though the benefits of this usage are less clear
- New disinfection systems, the majority of which rely on UVL, are under development, but much more research into the effectiveness and practicality of these technologies is needed
- Modern HVAC systems used in the US are generally very effective in maintaining appropriate air quality, as long as they are maintained correctly and new filters are regularly installed
- The benefits of LAF specifically are still unclear:

- Overall, ORs equipped with LAF systems have less airborne contamination than ORs with other types of ventilation, but this may not correlate with any decrease in infection risk to HCWs or patients
- SSI risk is already quite low in modern OR settings due to other interventions in the patient care pathway, especially antibiotic prophylaxis
- The level of airborne contamination at which the risk of infection increases is not known
- Modern HVAC systems of both LAF and non-LAF design may well be sufficient to maintain a level of air contamination that is well below the unknown threshold for increased infection risk
- The literature is largely in agreement that LAF systems function most effectively when airflow is directed at a downward angle, rather than horizontally or at any other angle
- Portable airflow devices can also improve air quality and reduce contamination, but may interrupt airflow patterns established by LAF and are generally not well-studied

Considering the broader context of the built environment, we can also draw some more general conclusions about our current understanding of airflow and infection risk in the OR. Standards for temperature and humidity in the OR are well-established, and within the current range do not have a notable impact on airflow as it relates to infection risk. It should be noted that the OR temperature can be low enough in some cases to require patient warming devices to maintain normothermia and avoid increased infection risk; these devices indeed may contribute to the complexity of OR air movement, but we did not explore this issue in detail as it is not specific to the built environment. Pressure is somewhat less standardized than humidity and temperature. Maintenance of positive pressure in the OR is the norm for most surgeries, although the COVID-19 pandemic has resulted in discussion of negative pressure ORs in the case of patients with known infectious diseases. Additionally, the pressure gradient – whether positive or negative – is sensitive to door openings, and therefore the degree of this gradient is not standardized. To the best of our knowledge an optimal value has not been established. Common approaches to understanding airflow include CFD modeling along with empirical measurements using air samplers and/or settle plates. Airborne contamination is usually then measured and reported in terms of particles and/or CFUs. Finally, it is worth noting that although this systematic review focused explicitly on the built environment, it is also important to consider how human behavior and movement as well as the presence, movement, and thermal properties of other objects can impact these outcomes.

Despite the long and extensive history of research into contamination risk in the operating room, however, we also found many notable gaps in the published literature. As we summarize below, these gaps limit our understanding of the built environment as it relates to airflow and infection risk.

## 5. RESEARCH GAPS SUMMARY

This systematic review was designed to provide a comprehensive overview of key features of the OR built environment and how they impact airflow and ultimately infection risk to both patients and healthcare workers. Despite the enormous body of literature related to infection risk in operating rooms, we found notable gaps in the existing research, especially as it relates to non-SSI infectious diseases and to HCWs.

Broadly, we found that research regarding the optimal OR layout is surprisingly lacking. Ideally, the different aspects of the built environment should be examined as an interacting complex system to better understand the relative contributions to turbulence in the room. We found that the majority of studies consider one aspect of the built environment in isolation, such as surgical lights or hinged vs. swinging doors. Research efforts that investigated interacting parts of the OR were largely limited to CFD models, with varying degrees of empirical validation.

We were surprised to find no studies in this review that explored the role of the built environment in the transmission of *respiratory* infections in ORs. Many critical questions are left unanswered. The potential for respiratory infection transmission in an OR setting (albeit perhaps much lower than in other parts of a healthcare facility), and the impact of the OR built environment has on transmission risk, is unknown. Best practices for monitoring respiratory infections in patients and HCWs have not been established, nor have risk mitigation practices such as masking patients or improving disinfection systems. A notable related gap in the research is the benefit of LAF versus other HVAC systems: despite an extensive body of literature, there is no conclusive evidence that LAF reduces SSI risk as compared to other systems. However, ORs equipped with LAF systems do typically have less airborne contamination than others. This reduction of airborne contamination could potentially lower the risk of respiratory infection transmission in LAF-equipped ORs, but this possibility has not been carefully explored.

Unlike respiratory infections, factors influencing the risk of SSIs have been studied extensively. However, it is difficult to establish causal links between OR conditions and SSIs, largely because the rates of SSI are very low in most modern care settings. Additionally, confounding factors are often present: for example, in some hospitals high infection risk surgeries are exclusively conducted in the most modernized ORs. In many publications, these confounding factors are actually part of the study design, in which infection prevention pathways or programs are implemented within a healthcare system and are designed to reduce the rate of SSIs as efficiently as possible. In this situation, interventions are implemented as a packaged effort that usually includes modifications to patient preparation and care. In such cases, it is difficult to determine causal links between SSI risk and built environment components. In fact, as we discussed previously regarding the equivocal evidence for the benefits of LAF, the relationship between air quality and subsequent SSI infection risk is not entirely clear in modern healthcare settings and patient care practices.

While this review explicitly excluded surgical equipment and human behavior, it is worth noting that among studies that identified the type of procedure or OR considered, the vast majority were focused on orthopedic procedures such as THAs and TKAs. More representative data are needed regarding other specialties and procedure types, and particularly aerosol-generating procedures (AGPs). Presumably AGPs can result in exposure and increase the respiratory infection risk of HCWs, but our review did not capture any articles that explored this possibility. Overall, we found a lack of attention to AGPs in the context of the built environment. The necessary OR characteristics to ensure patient and HCW safety and reduce transmission risk have not been established.

Finally, research concerning the risks to HCWs in general was also very limited. Most studies in our review that discuss personnel, rather than patients, focused on surgical smoke inhalation or potential UV exposure. Determining the risk of respiratory infection transmission to surgical staff, or how this could be established through future studies, was not discussed. Ultimately, many of the interacting factors impacting airflow and infection risk in the operating room environment remain poorly understood. Targeted studies to address these gaps would benefit the field of OR built environment research and improve health outcomes for HCWs and patients alike.

## Data Availability

This is a systematic literature review, as such all data discussed has been previously published.

## 6. ACKNOWLEDGMENTS

The authors would like to gratefully acknowledge the U.S. Centers for Disease Control and Prevention (CDC) for funding this work. This material is based upon work supported by the Naval Sea Systems Command under Contract No. N00024-13-D-6400, Task Order NH076. Any opinions, findings and conclusions or recommendations expressed in this material are those of the author(s) and do not necessarily reflect the views of the Naval Sea Systems Command (NAVSEA) or the U.S. Centers for Disease Control and Prevention.

The authors would also like to thank Ryan Darragh and Christopher Stiles for assistance with Figure 2.

## LIST OF ACRONYMS AND ABBREVIATIONS

AAD: angular air distribution
ACH: air changes per hour
ACOL: air-conditioner outlet layout
AGP: aerosol-generating procedure
ASHRAE: American Society of Heating, Refrigerating, and Air-Conditioning Engineers
BCP: bacteria-carrying particle
CAP: cold atmospheric-pressure plasma
CDC: Centers for Disease Control and Prevention
CED: continuous environmental disinfection
CFD: computational fluid dynamics
CFU: colony-forming units
COVID-19: coronavirus disease 2019
CSF: cerebral-spinal fluid
C-UVC: crystalline ultraviolet C
CV: conventional ventilation
DV: displacement ventilation
DVAF: differential vertical airflow ventilation
ENT: ear, nose, and throat
EPR: effective protection ratio
GNR: gram-negative rod
HAI: healthcare-associated infection
HCW: healthcare worker
HEPA: high-efficiency particulate air
HVAC: heating, ventilation, and air conditioning
IMA: Index of Microbial Air contamination
ISO: International Organization for Standardization
JHU/APL: Johns Hopkins University Applied Physics Laboratory
LAF: laminar airflow
LDAC: liquid desiccant air conditioning
LED: light-emitting diode
MDRO: multi-drug resistant organism
MLAF: mobile laminar airflow
MV: mixing ventilation
OR: operating room
OT: operating theater
PCNSI: post-operative central nervous system infection
PJI: prosthetic joint infection
PM: particulate matter
PPE: personal protective equipment
PPX-UVD: portable pulsed xenon ultraviolet disinfection
RDAC: rotary desiccant air conditioning
RODAC: replicate organism detection and counting
RR: relative risk
SARS-CoV-2: severe acute respiratory syndrome coronavirus 2
SSI: surgical site infection
TAF: temperature-controlled airflow
TBC: total bacterial count
TcAF: temperature-controlled airflow
THA: total hip arthroplasty
TI: turbulence intensity
TJA: total joint arthroplasty
TKA: total knee arthroplasty
TMA: turbulent mixed airflow
TMV: turbulent mixing ventilation
TPC: total particle count
TV: turbulent (airflow) ventilation
TVC: total viable count
TVOC: total volatile organic compound
UCV: ultra-clean ventilation
UDF: unidirectional airflow^1^
UDF: unidirectional displacement airflow^2^
UDF: unidirectional downflow^3^
UDV: unidirectional downward airflow
UFC: *undefined*^4^
UFP: ultra-fine particle
UV: ultraviolet
UV-C: ultraviolet-C
UVGI: ultraviolet germicidal irradiation
UVL: ultraviolet light
UWD: upward displacement airflow
VPC: viable particle count

## Footnotes

1 This is the most commonly used definition of “UDF” among the studies included in this review.

2 This definition of UDF comes from reference number ^107^

3 This definition of UDF comes from reference number ^94^

4 The authors of this study did not specify the definition of “UFC” beyond indicating that it was a unit for measuring microbial contamination^80^; this may be the result of a translation issue, and refer to the more common term “colony-forming unit” or “CFU”

## REFERENCES

1. Lister J. On the antiseptic principle in the practice of surgery. Br Med J. 1867;2(351):246–248. doi:10.1136/bmj.2.351.246

2. Lister J. An address on the present position of antiseptic surgery. Br Med J. 1890;2(1546):377–379. doi:10.1136/bmj.2.1546.377

3. Meleney FL. The control of wound infections. In: Transactions of the New York Surgical Society.; 1932:151–153.

4. Hoffman PN, Williams J, Stacey A, et al. Microbiological commissioning and monitoring of operating theatre suites: A report of a working party of the Hospital Infection Society. J Hosp Infect. 2002;52(1):1–28. doi:10.1053/jhin.2002.1237

5. Engineers AS of HR and C. ANSI/ASHRAE/ASHE Standard 170-2017: ASHRAE/ASHE Standard Ventilation of Health Care Facilities. Society of Heating, Refrigerating and Air-Conditioning Engineers (ASHRAE); 2021. https://ashrae.iwrapper.com/ASHRAE_PREVIEW_ONLY_STANDARDS/STD_170_2021

6. Hygiene requirements in operations and others invasive surgery. Bundesgesundheitsbl - Gesundheitsforsch - Gesundheitsschutz. 2000;43(8):644–648.

7. Fu Shaw L, Chen IH, Chen CS, et al. Factors influencing microbial colonies in the air of operating rooms. BMC Infect Dis. 2018;18:1–8. doi:10.1186/s12879-017-2928-1

8. NHFPC (National Health and Family Planning Commission of China). Special Requirements for Heating, Ventilation and Air-Conditioning (HVAC) Systems in Health Care Facilities. Canadian Standards Association; 2019. https://www.csagroup.org/store/product/CSA Z317.2:19/

9. NHFPC (National Health and Family Planning Commission of China). Architetural technical code for hospital clean operating department GB50333-2013. Published online 2013. https://www.codeofchina.com/standard/GB50333-2013.html

10. L’ASPEC. NF S 90351: Quels Changements 2003-2013.; 2013. https://www.aspec.fr

11. National Healthcare Safety Network (NHSN). Surgical Site Infection Event (SSI).; 2021.

12. Centers for Disease Control and Prevention (CDC). Current HAI Progress Report. Published 2019. https://www.cdc.gov/hai/data/portal/progress-report.html

13. Bolcato M, Rodriguez D, Aprile A. Risk Management in the New Frontier of Professional Liability for Nosocomial Infection: Review of the Literature on Mycobacterium Chimaera. Int J Env Res Public Heal. 2020;17(19 PG-). doi:10.3390/ijerph17197328

14. Chauveaux D. Preventing surgical-site infections: measures other than antibiotics. Orthop Traumatol Surg Res. 2015;101(1 Suppl PG-S77-83):S77–83. doi:10.1016/j.otsr.2014.07.028

15. Merollini KMD, Zheng H, Graves N. Most relevant strategies for preventing surgical site infection after total hip arthroplasty: Guideline recommendations and expert opinion. Am J Infect Control. 2013;41(3):221–226. doi:10.1016/j.ajic.2012.03.027

16. Mangram AJ, Horan TC, Pearson ML, Silver LC, Jarvis WR. Guideline for Prevention of Surgical Site Infection, 1999. Am J Infect Control. 1999;27(2):97–132. doi:10.1097/01.NAJ.0000521963.77728.c0

17. Berriós-Torres SI, Umscheid CA, Bratzler DW, et al. Centers for disease control and prevention guideline for the prevention of surgical site infection, 2017. JAMA Surg. 2017;152(8):784–791. doi:10.1001/jamasurg.2017.0904

18. Joseph A, Bayramzadeh S, Zamani Z, Rostenberg B. Safety, Performance, and Satisfaction Outcomes in the Operating Room: A Literature Review. Heal Environ Res Des J. 2018;11(2):137–150. doi:10.1177/1937586717705107

19. Moher D, Liberati A, Tetzlaff J, et al. Preferred reporting items for systematic reviews and meta-analyses: The PRISMA statement. PLoS Med. 2009;6(7). doi:10.1371/journal.pmed.1000097

20. Qualtrics. Published online 2005.

21. De Korne DF, Van Wijngaarden JDH, Van Rooij J, Wauben LSGL, Hiddema UF, Klazinga NS. Safety by design: effects of operating room floor marking on the position of surgical devices to promote clean air flow compliance and minimise infection risks. BMJ Qual Saf. 2012;21(9):746–752. doi:10.1136/bmjqs-2011-000138

22. D’Amico A, Montagna MT, Caggiano G, et al. Observational study on hospital building heritage and microbiological air quality in the orthopedic operating theater: The IM.PA.C.T. Project. Ann di Ig. 2019;31(5):482–495. doi:10.7416/ai.2019.2309

23. Curtis GL, Faour M, Jawad M, Klika AK, Barsoum WK, Higuera CA. Reduction of Particles in the Operating Room Using Ultraviolet Air Disinfection and Recirculation Units. J Arthroplasty. 2018;33(7):S196–S200. doi:10.1016/j.arth.2017.11.052

24. Anis HK, Curtis GL, Klika AK, et al. In-Room Ultraviolet Air Filtration Units Reduce Airborne Particles During Total Joint Arthroplasty. J Orthop Res. 2020;38(2):431–437. doi:10.1002/jor.24453

25. Green C, Pamplin JC, Chafin KN, Murray CK, Yun HC. Pulsed-xenon ultraviolet light disinfection in a burn unit: Impact on environmental bioburden, multidrug-resistant organism acquisition and healthcare associated infections. Burns. 2017;43(2):388–396. doi:10.1016/j.burns.2016.08.027

26. Cook TM, Piatt CJ, Barnes S, Edmiston CE. The Impact of Supplemental Intraoperative Air Decontamination on the Outcome of Total Joint Arthroplasty: A Pilot Analysis. J Arthroplasty. 2019;34(3):549–553. doi:10.1016/j.arth.2018.11.041

27. Dong C, Yuan H, Xu R, et al. Efficacy of infection control pathway in reducing postoperative infections in patients undergoing neurosurgery. J Infect Dev Ctries. 2020;14(1):74–79. doi:10.3855/jidc.11747

28. Evans RP. Current concepts for clean air and total joint arthroplasty: Laminar airflow and ultraviolet radiation: A systematic review. Clin Orthop Relat Res. 2011;469(4):945–953. doi:10.1007/s11999-010-1688-7

29. Prehn F, Timmermann E, Kettlitz M, Schaufler K, Günther S, Hahn V. Inactivation of airborne bacteria by plasma treatment and ionic wind for indoor air cleaning. Plasma Process Polym. 2020;17(9):e2000027. doi:10.1002/ppap.202000027

30. Murrell LJ, Hamilton EK, Johnson HB, Spencer M. Influence of a visible-light continuous environmental disinfection system on microbial contamination and surgical site infections in an orthopedic operating room. Am J Infect Control. 2019;47(7):804–810. doi:10.1016/j.ajic.2018.12.002

31. Sajadi B, Saidi MH, Ahmadi G. Computer modeling of the operating room ventilation performance in connection with surgical site infection. Sci Iran. 2020;27(2):704–714. doi:10.24200/sci.2018.5514.1359

32. Aganovic A, Cao G, Stenstad L-I, Skogas JG. An experimental study on the effects of positioning medical equipment on contaminant exposure of a patient in an operating room with unidirectional downflow. Build Environ. 2019;165. doi:10.1016/j.buildenv.2019.04.032

33. Cao G, Nilssen AM, Cheng Z, Stenstad LI, Radtke A, Skogås JG. Laminar airflow and mixing ventilation: Which is better for operating room airflow distribution near an orthopedic surgical patient? Am J Infect Control. 2019;47(7):737–743. doi:10.1016/j.ajic.2018.11.023

34. Külpmann R, Christiansen B, Kramer A, et al. Hygiene guideline for the planning, installation, and operation of ventilation and air-conditioning systems in health-care settings - Guideline of the German Society for Hospital Hygiene (DGKH). GMS Hyg Infect Control. 2016;11:Doc03–Doc03. doi:10.3205/dgkh000263

35. Kai T, Ayagaki N, Setoguchi H. Influence of the Arrangement of Surgical Light Axes on the Air Environment in Operating Rooms. J Healthc Eng. 2019;2019:1–8. doi:10.1155/2019/4861273

36. Sadeghian P, Wang C, Duwig C, Sadrizadeh S. Impact of surgical lamp design on the risk of surgical site infections in operating rooms with mixing and unidirectional airflow ventilation: A numerical study. J Build Eng. 2020;31. doi:10.1016/j.jobe.2020.101423

37. Cao G, Storås MCA, Aganovic A, Stenstad LI, Skogås JG. Do surgeons and surgical facilities disturb the clean air distribution close to a surgical patient in an orthopedic operating room with laminar airflow? Am J Infect Control. 2018;46(10):1115–1122. doi:10.1016/j.ajic.2018.03.019

38. Refaie R, Rushton P, McGovern P, et al. The effect of operating lights on laminar flow: an experimental study using neutrally buoyant helium bubbles. Bone Joint J. 2017;99-B(8):1061–1066. doi:10.1302/0301-620X.99B8.BJJ-2016-0581.R2

39. Aganovic A, Cao G, Stenstad LI, Skogås JG. Impact of surgical lights on the velocity distribution and airborne contamination level in an operating room with laminar airflow system. Build Environ. 2017;126:42–53. doi:10.1016/j.buildenv.2017.09.024

40. Zoon WAC, Loomans MGLC, Hensen JLM. Testing the effectiveness of operating room ventilation with regard to removal of airborne bacteria. Build Environ. 2011;46(12):2570–2577. doi:10.1016/j.buildenv.2011.06.015

41. Al-Waked R. Effect of Ventilation Strategies on Infection Control Inside Operating Theatres. Eng Appl Comput Fluid Mech. 2010;4(1):1–16. doi:10.1080/19942060.2010.11015295

42. Zoon WAC, van der Heijden MGM, Loomans MGLC, Hensen JLM. On the applicability of the laminar flow index when selecting surgical lighting. Build Environ. 2010;45(9):1976–1983. doi:10.1016/j.buildenv.2010.02.011

43. Traversari AAL, Bottenheft C, Louman R, van Heumen SPM, Böggemann J. The effect of operating lamps on the protected area of a unidirectional down flow (UDF) system. Heal Environ Res Des J. 2017;10(3):40–50. doi:10.1177/1937586716671292

44. Liu P, Zhang Y, Zheng Z, Li H, Liu X. LED surgical lighting system with multiple free-form surfaces for highly sterile operating theater application. Appl Opt. 2014;53(16):3427–3437. doi:10.1364/AO.53.003427

45. Kalava A, Midha M, Kurnutala LN, Schianodicola J, Yarmush JM. How clean are the overhead lights in operating rooms? Am J Infect Control. 2013;41(4):387–388. doi:10.1016/j.ajic.2012.04.335

46. Taaffe K, Lee B, Ferrand Y, et al. The Influence of Traffic, Area Location, and Other Factors on Operating Room Microbial Load. Infect Control Hosp Epidemiol. 2018;39(4):391–397. doi:10.1017/ice.2017.323

47. Wang C, Holmberg S, Sadrizadeh S. Impact of door opening on the risk of surgical site infections in an operating room with mixing ventilation. Indoor Built Environ. 2021;30(2):166–179. doi:10.1177/1420326X19888276

48. Erichsen Andersson A, Petzold M, Bergh I, Karlsson J, Eriksson BI, Nilsson K. Comparison between mixed and laminar airflow systems in operating rooms and the influence of human factors: Experiences from a Swedish orthopedic center. Am J Infect Control. 2014;42(6):665–669. doi:10.1016/j.ajic.2014.02.001

49. Perez P, Holloway J, Ehrenfeld L, et al. Door openings in the operating room are associated with increased environmental contamination. Am J Infect Control. 2018;46(8):954–956. doi:10.1016/j.ajic.2018.03.005

50. Mathijssen NMC, Hannink G, Sturm PDJ, et al. The Effect of Door Openings on Numbers of Colony Forming Units in the Operating Room during Hip Revision Surgery. Surg Infect (Larchmt). 2016;17(5):535–540. doi:10.1089/sur.2015.174

51. About. Healthcare Infection Society (HIS). doi:10.3917/afcul.105.0090

52. Smith EB, Raphael IJ, Maltenfort MG, Honsawek S, Dolan K, Younkins EA. The Effect of Laminar Air Flow and Door Openings on Operating Room Contamination. J Arthroplasty. 2013;28(9):1482–1485. doi:10.1016/j.arth.2013.06.012

53. El Awady MY, Abd El Rahman AT, Al Bagoury LS, Mossad IM. Air Quality in Ain Shams University Surgery Hospital. J Egypt Soc Parasitol. 2014;44(3):749–759. doi:10.12816/0007878

54. Nimra A, Ali Z, Khan MN, et al. Comparative ambient and indoor particulate matter analysis of operation theatres of government and private (trust) hospitals of Lahore, Pakistan. J Anim PLANT Sci. 2015;25(3, 2, SI):628–635.

55. Taaffe KM, Allen RW, Fredendall LD, et al. Simulating the effects of operating room staff movement and door opening policies on microbial load. Infect Control Hosp Epidemiol. Published online December 21, 2020:1–5. doi:10.1017/ice.2020.1359

56. Balocco C, Petrone G, Cammarata G, Vitali P, Albertini R, Pasquarella C. Experimental and numerical investigation on airflow and climate in a real operating theatre under effective use conditions. Int J Vent. 2015;13(4):351–368. doi:10.1080/14733315.2015.11684060

57. Villafruela JM, San José JF, Castro F, Zarzuelo A. Airflow patterns through a sliding door during opening and foot traffic in operating rooms. Build Environ. 2016;109:190–198. doi:10.1016/j.buildenv.2016.09.025

58. Roth JA, Juchler F, Dangel M, Eckstein FS, Battegay M, Widmer AF. Frequent door openings during cardiac surgery are associated with increased risk for surgical site infection: A prospective observational study. Clin Infect Dis. 2019;69(2):290–294. doi:10.1093/cid/ciy879

59. Young RS, O’Regan DJ. Cardiac surgical theatre traffic: time for traffic calming measures? Interact Cardiovasc Thorac Surg. 2010;10(4):526–529. doi:10.1510/icvts.2009.227116

60. Bediako-Bowan AAA, Mølbak K, Kurtzhals JAL, Owusu E, Debrah S, Newman MJ. Risk factors for surgical site infections in abdominal surgeries in Ghana: emphasis on the impact of operating rooms door openings. Epidemiol Infect. 2020;148, e147:1–5. doi:10.1017/S0950268820001454

61. Weiser MC, Shemesh S, Chen DD, Bronson MJ, Moucha CS. The Effect of Door Opening on Positive Pressure and Airflow in Operating Rooms. J Am Acad Orthop Surg. 2018;26(5):e105–e113. doi:10.5435/JAAOS-D-16-00891

62. Mears SC, Blanding R, Belkoff SM. Door Opening Affects Operating Room Pressure During Joint Arthroplasty. Orthopedics. 2015;38(11):e991–e994. doi:10.3928/01477447-20151020-07

63. Balocco C, Petrone G, Cammarata G. Assessing the effects of sliding doors on an operating theatre climate. Build Simul. 2012;5(1):73–83. doi:10.1007/s12273-012-0071-x

64. Alsved M, Civilis A, Ekolind P, et al. Temperature-controlled airflow ventilation in operating rooms compared with laminar airflow and turbulent mixed airflow. J Hosp Infect. 2018;98(2):181–190. doi:10.1016/j.jhin.2017.10.013

65. Rezapoor M, Alvand A, Jacek E, Paziuk T, Maltenfort MG, Parvizi J. Operating Room Traffic Increases Aerosolized Particles and Compromises the Air Quality: A Simulated Study. J Arthroplasty. 2018;33(3):851–855. doi:10.1016/j.arth.2017.10.012

66. Liang C-C, Wu F-J, Chien T-Y, et al. Effect of ventilation rate on the optimal air quality of trauma and colorectal operating rooms. Build Environ. 2020;169:106548. doi:10.1016/j.buildenv.2019.106548

67. Stauning MT, Bediako-Bowan A, Andersen LP, et al. Traffic flow and microbial air contamination in operating rooms at a major teaching hospital in Ghana. J Hosp Infect. 2018;99(3):263–270. doi:10.1016/j.jhin.2017.12.010

68. Sadrizadeh S, Pantelic J, Sherman M, Clark J, Abouali O. Airborne particle dispersion to an operating room environment during sliding and hinged door opening. J Infect Public Health. 2018;11(5):631–635. doi:10.1016/j.jiph.2018.02.007

69. Birgand G, Azevedo C, Rukly S, et al. Motion-capture system to assess intraoperative staff movements and door openings: Impact on surrogates of the infectious risk in surgery. Infect Control Hosp Epidemiol. 2019;40(5):566–573. doi:10.1017/ice.2019.35

70. Agodi A, Auxilia F, Barchitta M, et al. Operating theatre ventilation systems and microbial air contamination in total joint replacement surgery: results of the GISIO-ISChIA study. J Hosp Infect. 2015;90(3):213–219. doi:10.1016/j.jhin.2015.02.014

71. Andersson AE, Bergh I, Karlsson J, Eriksson BI, Nilsson K. Traffic flow in the operating room: An explorative and descriptive study on air quality during orthopedic trauma implant surgery. Am J Infect Control. 2012;40(8):750–755. doi:10.1016/j.ajic.2011.09.015

72. Teter J, Guajardo I, Al-Rammah T, Rosson G, Perl TM, Manahan M. Assessment of operating room airflow using air particle counts and direct observation of door openings. Am J Infect Control. 2017;45(5):477–482. doi:10.1016/j.ajic.2016.12.018

73. Zhou B, Ding L, Li F, Xue K, Nielsen P V., Xu Y. Influence of opening and closing process of sliding door on interface airflow characteristic in operating room. Build Environ. 2018;144(August):459–473. doi:10.1016/j.buildenv.2018.08.050

74. Lydon GP, Ingham DB, Mourshed MM. Ultra clean ventilation system performance relating to airborne infections in operating theatres using CFD modelling. Build Simul. 2014;7(3):277–287. doi:10.1007/s12273-013-0145-4

75. Stather P, Salji M, Hassan SU, et al. A comparison of airborne bacterial fallout between orthopaedic and vascular surgery. Ann R Coll Surg Engl. 2017;99(4):295–298. doi:10.1308/rcsann.2016.0352

76. Birgand G, Toupet G, Rukly S, et al. Air contamination for predicting wound contamination in clean surgery: A large multicenter study. Am J Infect Control. 2015;43(5):516–521. doi:10.1016/j.ajic.2015.01.026

77. Oguz R, Diab-Elschahawi M, Berger J, et al. Airborne bacterial contamination during orthopedic surgery: A randomized controlled pilot trial. J Clin Anesth. 2017;38:160–164. doi:10.1016/j.jclinane.2017.02.008

78. Nasir ZA, Mula V, Stokoe J, Colbeck I, Loeffler M. Evaluation of total concentration and size distribution of bacterial and fungal aerosol in healthcare built environments. Indoor Built Environ. 2015;24(2):269–279. doi:10.1177/1420326X13510925

79. Diab-Elschahawi M, Berger J, Blacky A, et al. Impact of different-sized laminar air flow versus no laminar air flow on bacterial counts in the operating room during orthopedic surgery. Am J Infect Control. 2011;39(7):e25–e29. doi:10.1016/j.ajic.2010.10.035

80. Barbadoro P, Bruschi R, Martini E, et al. Impact of laminar air flow on operating room contamination, and surgical wound infection rates in clean and contaminated surgery. Eur J Surg Oncol. 2016;42(11):1756–1758. doi:10.1016/j.ejso.2016.06.409

81. Squeri R, Genovese C, Trimarchi G, et al. Nine years of microbiological air monitoring in the operating theatres of a university hospital in Southern Italy. Ann DI Ig Med Prev E DI COMUNITA. 2019;31(1, SI):1–12. doi:10.7416/ai.2019.2272

82. Singh VK, Hussain S, Javed S, Singh I, Mulla R, Kalairajah Y. Sterile surgical helmet system in elective total hip and knee arthroplasty. J Orthop Surg (Hong Kong). 2011;19(2):234–237. doi:10.1177/230949901101900222

83. Din A, Foden P, Mathew M, Periasamy K. Does laminar flow reduce the risk of early surgical site infection in hip fracture patients? J Orthop. 2020;18:13–15. doi:10.1016/j.jor.2019.08.026

84. Pinder EM, Bottle A, Aylin P, Loeffler MD. Does laminar flow ventilation reduce the rate of infection? an observational study of trauma in England. Bone Joint J. 2016;98-B(9):1262–1269. doi:10.1302/0301-620X.9869.37184

85. Hooper GJ, Rothwell AG, Frampton C, Wyatt MC. Does the use of laminar flow and space suits reduce early deep infection after total hip and knee replacement?: the ten-year results of the New Zealand Joint Registry. J BONE Jt SURGERY-BRITISH Vol. 2011;93-B(1):85–90. doi:10.1302/0301-620X.93B1.24862

86. Agarwal SK, Khan AA, Solan M, Lemon M. Hip fracture surgery in mixed-use emergency theatres: is the infection risk increased? A retrospective matched cohort study. Ann R Coll Surg Engl. 2017;99(8):641–644. doi:10.1308/rcsann.2017.0183

87. Jeong SJ, Ann HW, Kim JK, et al. Incidence and risk factors for surgical site infection after gastric surgery: A multicenter prospective cohort study. Infect Chemother. 2013;45(4):422–430. doi:10.3947/ic.2013.45.4.422

88. Kirschbaum S, Hommel H, Strache P, Horn R, Falk R, Perka C. Laminar air flow reduces particle load in TKA-even outside the LAF panel: a prospective, randomized cohort study. Knee Surgery, Sport Traumatol Arthrosc. Published online 2020:1–9. doi:10.1007/s00167-020-06344-3

89. Breier A-C, Brandt C, Sohr D, Geffers C, Gastmeier P. Laminar Airflow Ceiling Size: No Impact on Infection Rates Following Hip and Knee Prosthesis. Infect Control Hosp Epidemiol. 2011;32(11):1097–1102. doi:10.1086/662182

90. Teo BJX, Woo YL, Phua JKS, Chong H-C, Yeo W, Tan AHC. Laminar flow does not affect risk of prosthetic joint infection after primary total knee replacement in Asian patients. J Hosp Infect. 2020;104(3):305–308. doi:10.1016/j.jhin.2019.12.014

91. Bosanquet D, Jones CN, Gill N, Jarvis P, Lewis MH. Laminar flow reduces cases of surgical site infections in vascular patients. Ann R Coll Surg Engl. 2013;95(1):15–19. doi:10.1308/003588413X13511609956011

92. Langvatn H, Schrama JC, Cao G, et al. Operating room ventilation and the risk of revision due to infection after total hip arthroplasty: assessment of validated data in the Norwegian Arthroplasty Register. J Hosp Infect. 2020;105(2):216–224. doi:10.1016/j.jhin.2020.04.010

93. Smith JO, Frampton CMA, Hooper GJ, Young SW. The Impact of Patient and Surgical Factors on the Rate of Postoperative Infection After Total Hip Arthroplasty—A New Zealand Joint Registry Study. J Arthroplasty. 2018;33(6):1884–1890. doi:10.1016/j.arth.2018.01.021

94. Traversari AAL, van Heumen SPM, van Tiem FLJ, Bottenheft C, Hinkema MJ. Design variables with significant effect on system performance of unidirectional displacement airflow systems in hospitals. J Hosp Infect. 2019;103(1):e81–e87. doi:10.1016/j.jhin.2019.03.009

95. Traversari AAL, Bottenheft C, van Heumen SPM, Goedhart CA, Vos MC. Effect of switching off unidirectional downflow systems of operating theaters during prolonged inactivity on the period before the operating theater can safely be used. Am J Infect Control. 2017;45(2):139–144. doi:10.1016/j.ajic.2016.07.019

96. Elnour AA, Abdelfattah MM, Negm S, Kassim T. Microbiological Surveillance of Air Quality: A comparative Study Using Active and Passive Methods in Operative Theater. Int J Pharm Phytopharm Res. 2018;8(1):33–38.

97. Yunusa U, Golfa T, Dathini H. Prognosticators of surgical site infections (SSIs) among patients undergoing major surgery at general hospital Funtua, Katsina State, Nigeria. Pielęgniarstwo Chir i Angiol. 2015;2:111–117.

98. Alfonso-Sanchez JL, Martinez IM, Martín-Moreno JM, González RS, Botía F. Analyzing the risk factors influencing surgical site infections: The site of environmental factors. Can J Surg. 2017;60(3):155–161. doi:10.1503/cjs.017916

99. Hirsch T, Hubert H, Fischer S, et al. Bacterial burden in the operating room: Impact of airflow systems. Am J Infect Control. 2012;40(7):e228–e232. doi:10.1016/j.ajic.2012.01.007

100. Chien T-Y, Liang C-C, Wu F-J, Chen C-T, Pan T-H, Wan G-H. Comparative Analysis of Energy Consumption, Indoor Thermal-Hygrometric Conditions, and Air Quality for HVAC, LDAC, and RDAC Systems Used in Operating Rooms. Appl Sci. 2020;10(11):1–16. doi:10.3390/app10113721

101. Lee S-T, Liang C-C, Chien T-Y, Wu F-J, Fan K-C, Wan G-H. Effect of ventilation rate on air cleanliness and energy consumption in operation rooms at rest. Environ Monit Assess. 2018;190(3). doi:10.1007/s10661-018-6556-z

102. Romano F, Gusten J, De Antonellis S, Joppolo CM. Electrosurgical Smoke: Ultrafine Particle Measurements and Work Environment Quality in Different Operating Theatres. Int J Environ Res Public Health. 2017;14(2). doi:10.3390/ijerph14020137

103. Pasquarella C, Barchitta M, D’Alessandro D, et al. Heating, ventilation and air conditioning (HVAC) system, microbial air contamination and surgical site infection in hip and knee arthroplasties: The GISIO-SItI Ischia study. Ann di Ig. 2018;30(5 Suppl. 2):22–35. doi:10.7416/ai.2018.2248

104. Pereira ML, Vilain R, Kawase PR, Tribess A, Morawska L. Impact of Filtration Conditions on Air Quality in an Operating Room. Int J Environ Res. 2020;14(6):685–692. doi:10.1007/s41742-020-00286-x

105. Chidambaram S, Vasudevan MC, Nair MN, Joyce C, Germanwala A V. Impact of Operating Room Environment on Postoperative Central Nervous System Infection in a Resource-Limited Neurosurgical Center in South Asia. World Neurosurg. 2018;110:e239–e244. doi:10.1016/j.wneu.2017.10.142

106. Romano F, Milani S, Ricci R, Joppolo CM. Operating Theatre Ventilation Systems and Their Performance in Contamination Control: “At Rest” and “In Operation” Particle and Microbial Measurements Made in an Italian Large and Multi-Year Inspection Campaign. Int J Environ Res Public Health. 2020;17(19):7275. doi:10.3390/ijerph17197275

107. Fischer S, Thieves M, Hirsch T, et al. Reduction of airborne bacterial burden in the OR by installation of unidirectional displacement airflow (UDF) systems. Med Sci Monit. 2015;21:2367–2374. doi:10.12659/MSM.894251

108. Romano F, Milani S, Gustén J, Joppolo CM. Surgical Smoke and Airborne Microbial Contamination in Operating Theatres: Influence of Ventilation and Surgical Phases. Int J Environ Res Public Health. 2020;17(15):5395. doi:10.3390/ijerph17155395

109. Albertini R, Colucci ME, Turchi S, Vitali P. The management of air contamination control in operating theaters: the experience of the Parma University Hospital (IT). Aerobiologia (Bologna). 2020;36(1, SI):119–123. doi:10.1007/s10453-019-09572-4

110. Montagna MT, Rutigliano S, Trerotoli P, et al. Evaluation of Air Contamination in Orthopaedic Operating Theatres in Hospitals in Southern Italy: The IMPACT Project. Int J Environ Res Public Health. 2019;16(19):3581. doi:10.3390/ijerph16193581

111. Srivastava S, Vasavada V, Vasavada AR, Sudhalkar A, Kothari A, Vasavada SA. Realtime Imaging of Airflow Patterns and Impact of Infection Control Measures in Ophthalmic Practice. J Cataract Refract Surg. 2020;Publish Ah. doi:10.1097/j.jcrs.0000000000000538

112. Wagner JA, Schreiber KJ, Cohen R. Using Cleanroom Technology: Improving Operating Room Contamination Control. ASHRAE J. 2014;56(2):18–27.

113. Loth AG, Guderian DB, Haake B, Zacharowski K, Stöver T, Leinung M. Aerosol Exposure During Surgical Tracheotomy in SARS-CoV-2 Positive Patients. Shock. 2021;Publish Ah. doi:10.1097/SHK.0000000000001655

114. Pereira ML, Vilain R, Galvão FHF, Tribess A, Morawska L. Experimental and numerical analysis of the relationship between indoor and outdoor airborne particles in an operating room. Indoor Built Environ. 2013;22(6):864–875. doi:10.1177/1420326X12460707

115. Xue K, Cao G, Liu M, et al. Experimental study on the effect of exhaust airflows on the surgical environment in an operating room with mixing ventilation. J Build Eng. 2020;32. doi:10.1016/j.jobe.2020.101837

116. Shirozu K, Setoguchi H, Araki K, Ando T, Yamaura K. Impact of air-conditioner outlet layout on the upward airflow induced by forced air warming in operating rooms. Am J Infect Control. 2021;49(1):44–49. doi:10.1016/j.ajic.2020.06.202

117. Wagner JA, Greeley DG, Gormley TC, Markel TA. Comparison of operating room air distribution systems using the environmental quality indicator method of dynamic simulated surgical procedures. Am J Infect Control. 2019;47(1):e1–e6. doi:10.1016/j.ajic.2018.07.020

118. Wagner JA, Dexter F, Greeley DG, Schreiber K. Operating room air delivery design to protect patient and surgical site results in particles released at surgical table having greater concentration along walls of the room than at the instrument tray. Am J Infect Control. 2020;000:1–4. doi:10.1016/j.ajic.2020.10.003

119. Gormley T, Markel TA, Jones HW, et al. Methodology for analyzing environmental quality indicators in a dynamic operating room environment. Am J Infect Control. 2017;45(4):354–359. doi:10.1016/j.ajic.2016.11.001

120. COMSOL Multiphysics. www.comsol.com

121. Agirman A, Cetin YE, Avci M, Aydin O. Effect of laminar airflow unit diffuser size on pathogen particle distribution in an operating room. Sci Technol Built Environ. 2020;0(0):1–12. doi:10.1080/23744731.2020.1816405

122. Wong KY, M. Kamar H, Kamsah N. Enhancement of Airborne Particles Removal in a Hospital Operating Room. Int J Automot Mech Eng. 2019;16(4):7447–7463. doi:10.15282/ijame.16.4.2019.17.0551

123. Amiraslanpour M, Ghazanfarian J, Nabaei H, Taleghani MH. Evaluation of laminar airflow heating, ventilation, and air conditioning system for particle dispersion control in operating room including staffs: A non-Boussinesq Lagrangian study. J Build Phys. 2020;00(0):1–29. doi:10.1177/1744259120932932

124. Keshtkar MM, Nafteh M. Investigation of influence of linear diffuser in the ventilation of operating rooms. Adv ENERGY Res. 2016;4(3):239–253. doi:10.12989/eri.2016.4.3.239

125. Ufat H, Kaynakli O, Yamankaradeniz N, Yamankaradeniz R. Investigation of the number of particles in an operating room at different ambient temperatures and inlet velocities. Int J Vent. 2018;17(3):209–223. doi:10.1080/14733315.2017.1392107

126. Sajadi B, Saidi MH, Ahmadi G. Numerical evaluation of the operating room ventilation performance: Ultra-Clean Ventilation (UCV) systems. Sci Iran. 2019;26(4):2394–2406. doi:10.24200/sci.2018.5431.1269

127. Wang C, Holmberg S, Sadrizadeh S. Numerical study of temperature-controlled airflow in comparison with turbulent mixing and laminar airflow for operating room ventilation. Build Environ. 2018;144:45–56. doi:10.1016/j.buildenv.2018.08.010

128. Wang F, Hung J, Chen Y, Hsu C. Performance evaluation for operation rooms by numerical simulation and field measurement. Int J Vent. 2017;16(3):189–199. doi:10.1080/14733315.2017.1299515

129. Zhai ZJ, Osborne AL. Simulation-based feasibility study of improved air conditioning systems for hospital operating room. Front Archit Res. 2013;2(4):468–475. doi:10.1016/j.foar.2013.09.003

130. Sadrizadeh S, Holmberg S. Surgical clothing systems in laminar airflow operating room: a numerical assessment. J Infect Public Health. 2014;7(6):508–516. doi:10.1016/j.jiph.2014.07.011

131. Balocco C, Petrone G, Cammarata G. Thermo-fluid dynamics analysis and air quality for different ventilation patterns in an operating theatre. Int J Heat Technol. 2015;33(4):25–32. doi:10.18280/ijht.330404

132. Ufat H, Kaynakli O, Yamankaradeniz N, Yamankaradeniz R. Three-dimensional air distribution analysis of different outflow typed operating rooms at different inlet velocities and room temperatures. Adv Mech Eng. 2017;9(7):1–12. doi:10.1177/1687814017707414

133. Yau YH, Ding LC. A case study on the air distribution in an operating room at Sarawak General Hospital Heart Centre (SGHHC) in Malaysia. Indoor Built Environ. 2014;23(8):1129–1141. doi:10.1177/1420326X13499359

134. Yau YH, Ding LC. A comprehensive computational fluid dynamics simulation on the air distribution in an operating room at University of Malaya Medical Centre Malaysia. Indoor Built Environ. 2015;24(3):355–369. doi:10.1177/1420326X13516349

135. Khankari K. Computational Fluid Dynamics (CFD) Analysis of Hospital Operating Room Ventilation Systems - Part II: Analyses of HVAC Configurations. ASHRAE J. 2018;60(6):16–26.

136. Rahate SD, Sarode AD. Design of Air Distribution System for Operation Theatre Using Flow Visualization Techniques to Improve Flow Characteristics. Int J Eng. 2020;33(1):164–169. doi:10.5829/ije.2020.33.01a.19

137. Khankari K. Dynamics of unidirectional airflow. ASHRAE J. 2019;61(7):20–39.

138. Sadrizadeh S, Holmberg S, Tammelin A. A numerical investigation of vertical and horizontal laminar airflow ventilation in an operating room. Build Environ. 2014;82:517–525. doi:10.1016/j.buildenv.2014.09.013

139. Agirman A, Cetin YE, Avci M, Aydin O. Effect of air exhaust location on surgical site particle distribution in an operating room. Build Simul. 2020;13(5, SI):979–988. doi:10.1007/s12273-020-0642-1

140. Baracat TM, da Silva CA, Lofrano FC, Kurokawa FA. Assessment of the performance of airflow in an operating rooms using ceiling supply and sidewall inlet systems. J BRAZILIAN Soc Mech Sci Eng. 2020;42(41):1–13. doi:10.1007/s40430-019-2117-9

141. Abed IM, Amer R. Modeling and Experimental Investigation of Laminar Ceiling Air Distribution System for Operating Room in Merjan Teaching Hospital. J Eng Technol Sci. 2018;50(6):870–883. doi:10.5614/j.eng.technol.sci.2018.50.6.9

142. Liu Z, Liu H, Yin H, Rong R, Cao G, Deng Q. Prevention of surgical site infection under different ventilation systems in operating room environment. Front Environ Sci Eng. 2021;15(3):36. doi:10.1007/s11783-020-1327-9

143. Khankari K. Computational Fluid Dynamics (CFD) Analysis of Hospital Operating Room Ventilation Systems - Part I: Analysis of Air Change Rates. ASHRAE J. 2018;60(5):14–26.

144. Bahador M, Keshtkar MM. Reviewing and modeling the optimal output velocity of slot linear diffusers to reduce air contamination in the surgical site of operating rooms. Int J Comput Sci Netw Secur. 2017;17(8):82–89.

145. Gormley T, Markel TA, Jones H, et al. Cost-benefit analysis of different air change rates in an operating room environment. Am J Infect Control. 2017;45(12):1318–1323. doi:10.1016/j.ajic.2017.07.024

146. Vonci N, De Marco MF, Grasso A, Spataro G, Cevenini G, Messina G. Association between air changes and airborne microbial contamination in operating rooms. J Infect Public Health. 2019;12(6):827–830. doi:10.1016/j.jiph.2019.05.010

147. Zhang Y, Cao G, Feng G, et al. The impact of air change rate on the air quality of surgical microenvironment in an operating room with mixing ventilation. J Build Eng. 2020;32. doi:10.1016/j.jobe.2020.101770

148. Morris BJ, Kiser CJ, Laughlin MS, et al. A localized laminar flow device decreases airborne particulates during shoulder arthroplasty: a randomized controlled trial. J Shoulder Elb Surg. 2021;30(3):580–586. doi:10.1016/j.jse.2020.08.035

149. Nilsson K, Lundholm R, Friberg S. Assessment of horizontal laminar air flow instrument table for additional ultraclean space during surgery. J Hosp Infect. 2010;76(3):243–246. doi:10.1016/j.jhin.2010.05.016

150. Sossai D, Dagnino G, Sanguineti F, Franchin F. Mobile laminar air flow screen for additional operating room ventilation: reduction of intraoperative bacterial contamination during total knee arthroplasty. J Orthop Traumatol. 2011;12(4):207–211. doi:10.1007/s10195-011-0168-5

151. Sadrizadeh S, Holmberg S. Effect of a portable ultra-clean exponential airflow unit on the particle distribution in an operating room. PARTICUOLOGY. 2015;18:170–178. doi:10.1016/j.partic.2014.06.002

152. Lapid-Gortzak R, Traversari R, van der Linden JW, Lesnik Oberstein SY, Lapid O, Schlingemann RO. Mobile ultra-clean unidirectional airflow screen reduces air contamination in a simulated setting for intra-vitreal injection. Int Ophthalmol. 2017;37(1):131–137. doi:10.1007/s10792-016-0236-1

153. Sadrizadeh S, Holmberg S, Nielsen P V. Three distinct surgical clothing systems in a turbulent mixing operating room equipped with mobile ultraclean laminar airflow screen: A numerical evaluation. Sci Technol Built Environ. 2016;22(3):337–345. doi:10.1080/23744731.2015.1113838

154. Stocks GW, O’Connor DP, Self SD, Marcek GA, Thompson BL. Directed Air Flow to Reduce Airborne Particulate and Bacterial Contamination in the Surgical Field During Total Hip Arthroplasty. J Arthroplasty. 2011;26(5):771–776. doi:10.1016/j.arth.2010.07.001

155. Sadrizadeh S, Tammelin A, Nielsen P V, Holmberg S. Does a mobile laminar airflow screen reduce bacterial contamination in the operating room? A numerical study using computational fluid dynamics technique. Patient Saf Surg. 2014;8(27). doi:10.1186/1754-9493-8-27

156. Casagrande D, Piller M. Conflicting effects of a portable ultra-clean airflow unit on the sterility of operating rooms: A numerical investigation. Build Environ. 2020;171. doi:10.1016/j.buildenv.2020.106643

157. von Vogelsang A-C, Förander P, Arvidsson M, Löwenhielm P. Effect of mobile laminar airflow units on airborne bacterial contamination during neurosurgical procedures. J Hosp Infect. 2018;99(3):271–278. doi:10.1016/j.jhin.2018.03.024

158. Loomans MGLC, de Visser IM, Loogman JGH, Kort HSM. Alternative ventilation system for operating theaters: Parameter study and full-scale assessment of the performance of a local ventilation system. Build Environ. 2016;102:26–38. doi:10.1016/j.buildenv.2016.03.012

159. Li H, Zhong K, Zhai Z (John). Investigating the influences of ventilation on the fate of particles generated by patient and medical staff in operating room. Build Environ. 2020;180:107038. doi:10.1016/j.buildenv.2020.107038

