## Supplemental Information for "Influence of the Built Environment on Airflow, Contamination, and Infection in the Operating Room: A Systematic Literature Review"

Shirley M. Klimkiewicz[1]; Molly E. Gallagher, Ph.D.[1]; Anastasia S. Lambrou, Ph.D.[1]; Oluwaferanmi E. Adeyemo[1]; Amy M. Andrelchik[1]; Kavita Braun[1]; Madeline B. Ford, M.S.[1]; Terrence J. Garcia[1]; Stephanie Ku, M.S.[1]; Kaitlin Rainwater-Lovett, Ph.D.[1]; Julee A. Rendon, M.P.H.[1]; Sophia H. Oluic, M.P.H.[1]; Steven L. Patterson, M.S.[1]; Joshua Yoon, Ph.D.[1]; Alexander J. Yuan[1]; Winnie Wang, M.P.H.[1]; Lucy Carruth, Ph.D.[1]; Brian Damit, Ph.D.[1,†]

[1]The Johns Hopkins University Applied Physics Laboratory, Laurel MD

Appendix S1. Geographical Information .......... 1–6

Appendix S2. Operating Room Characteristics .......... 2–1

Appendix S3. Literature reviews and meta-analyses .......... 3–1
- Disinfection systems .......... 3–1
- Operating room doors .......... 3–1
- Operating room ventilation system .......... 3–2

Appendix S4. Summary of Included Articles .......... 4–1
- Operating room layout .......... 4–1
- Disinfection systems .......... 4–1
- Operating room lights .......... 4–3
- Operating room doors .......... 4–7
- Operating room ventilation .......... 4–15
- Portable airflow systems .......... 4–37

Appendix S5. PRISMA Checklist .......... 5–1

#### LIST OF ACRONYMS AND ABBREVIATIONS

| | |
|---|---|
| AAD | angular air distribution |
| ACH | air changes per hour |
| ACOL | air-conditioner outlet layout |
| BCP | bacteria-carrying particle |
| CDC | Centers for Disease Control and Prevention |
| CED | continuous environmental disinfection |
| CFD | computational fluid dynamics |
| CFU | colony-forming units |
| CSF | cerebral-spinal fluid |
| CV | conventional ventilation |
| DV | displacement ventilation |
| DVAF | differential vertical airflow ventilation |
| ENT | ear, nose, and throat |
| EPR | effective protection ratio |
| GNR | gram-negative rod |
| HAI | hospital-acquired infection |
| HCW | healthcare worker |
| HEPA | high-efficiency particulate air |
| HVAC | heating, ventilation, and air conditioning |
| IMA | Index of Microbial Air contamination |
| ISO | International Organization for Standardization |
| JHU/APL | Johns Hopkins University Applied Physics Laboratory |
| LAF | laminar airflow |
| LDAC | liquid desiccant air conditioning |
| LED | light-emitting diode |
| MDRO | multi-drug resistant organism |
| MLAF | mobile laminar airflow |
| MV | mixing ventilation |
| OR | operating room |
| OT | operating theater |
| PCNSI | post-operative central nervous system infection |
| PJI | prosthetic joint infection |
| PM | particulate matter |
| PPE | personal protective equipment |
| PPX-UVD | portable pulsed xenon ultraviolet disinfection |
| RDAC | rotary desiccant air conditioning |
| RODAC | replicate organism detection and counting |
| RR | relative risk |
| SSI | surgical site infection |
| TAF | temperature-controlled airflow |
| TBC | total bacterial count |
| TcAF | temperature-controlled airflow |
| THA | total hip arthroplasty |
| TI | turbulence intensity |

| | |
|---|---|
| TJA | total joint arthroplasty |
| TKA | total knee arthroplasty |
| TMA | turbulent mixed airflow |
| TMV | turbulent mixing ventilation |
| TPC | total particle count |
| TV | turbulent (airflow) ventilation |
| TVC | total viable count |
| TVOC | total volatile organic compound |
| UCV | ultra-clean ventilation |
| UDF | unidirectional airflow[1] |
| UDF | unidirectional displacement airflow[2] |
| UDF | unidirectional downflow[3] |
| UFC | *undefined*[4] |
| UFP | ultra-fine particle |
| UV | ultraviolet |
| UVGI | ultraviolet germicidal irradiation |
| UVL | ultraviolet light |
| UWD | upward displacement airflow |
| VPC | viable particle count |

[1] This is the most commonly used definition of "UDF" among the studies included in this review.

[2] This definition of UDF comes from reference number [126]

[3] This definition of UDF comes from reference number [98]

[4] The authors of this study did not specify the definition of "UFC" beyond indicating that it was a unit for measuring microbial contamination[95]; this may be the result of a translation issue, and refer to the more common term "colony-forming unit" or "CFU"

#### LIST OF FIGURES

Figure S1. Distribution of the 138 included articles by country.............................................1–6

Figure S2. Plot of OR dimensions collected from the publications captured by this systematic review. The dimensions of real ORs and those used in modeling studies are both included...............................................................................................................2–1

Figure S3. Distribution of OR floor areas collected from publications captured by this systematic review. The dimensions of real ORs and those used in modeling studies are both included............................................................................................2–2

Figure S4. Distribution of OR volumes collected from the publications captured in this systematic review. The dimensions of real ORs and those used in modeling studies are both included............................................................................................2–2

**LIST OF TABLES**

Table S1. Literature reviews and meta-analyses not included in this systematic literature review. ........ 3–1

Table S2. Summary of the two articles included that investigated the impact of the operating room layout on airflow, contamination, and infections. ........ 4–1

Table S3. Summary of the several articles included that investigated the impact of disinfection systems on airflow, contaminations, and infections. ........ 4–1

Table S4. Summary of the fifteen articles included that investigated the impact of lights in the operating room on airflow, contamination, and infections. ........ 4–3

Table S5. Summary of the 33 articles included that investigated the impact of doors in the operating room on airflow, contamination, and infections. ........ 4–7

Table S6. Summary of the 86 articles included that investigated the impact of ventilation in the operating room on airflow, contamination, and infections. ........ 4–15

Table S7. Summary of the 12 articles included that investigated the impact of portable airflow systems in the operating room on airflow, contaminations, and infections. ........ 4–37

Table S8. PRISMA 2020 Checklist[159] ........ 5–1

### APPENDIX S1.GEOGRAPHICAL INFORMATION

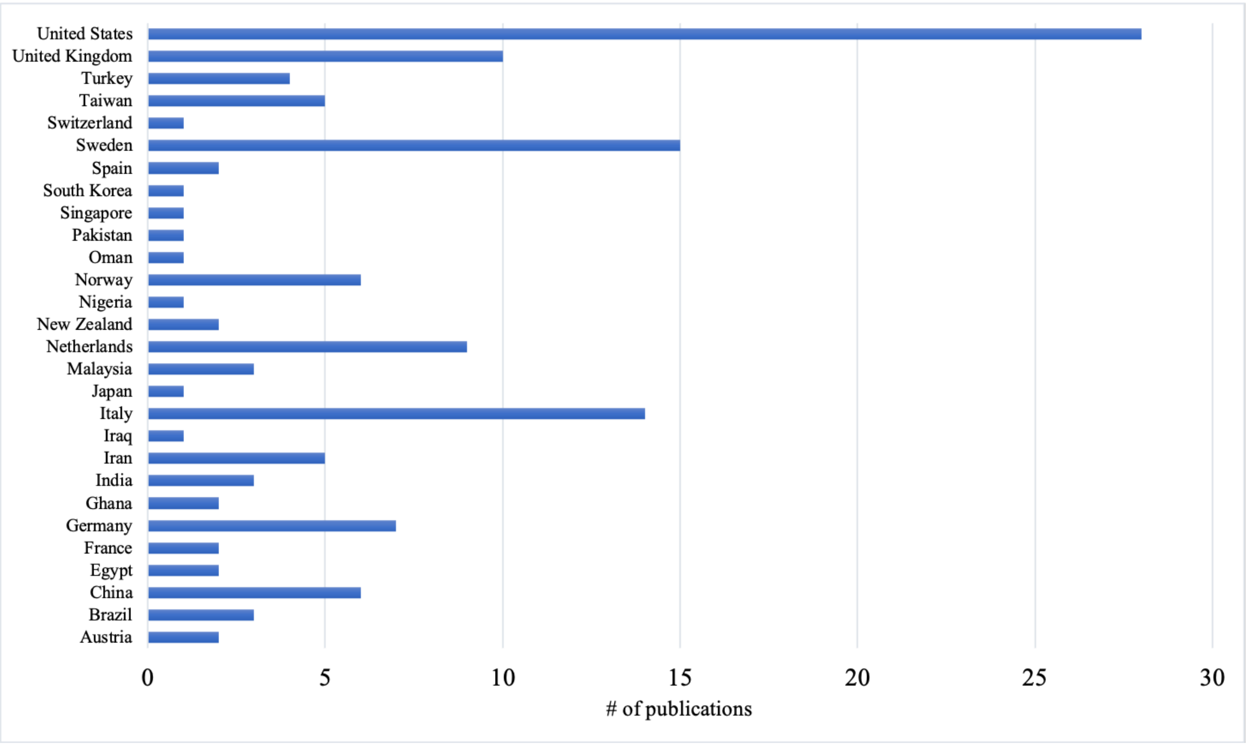

**Figure S1.** Distribution of the 138 included articles by country.

### APPENDIX S2. OPERATING ROOM CHARACTERISTICS

As part of the data extraction process, various operating room characteristics such as specialty, age, and overall dimensions were collected when available. Both the real-world measurements of ORs investigated in empirical studies and hypothetical dimensions used in CFD models were collected, as the latter were often based on specific existing ORs. While most articles reported such characteristics, they were not consistent in the type of information reported: some articles only reported area or volume without providing ceiling height, thus not allowing calculation of the missing dimension, while other articles provided both measurements. As such, a few different strategies were taken to summarize these data. **Error! Reference source not found.** features the measurements collected from all articles that reported enough information to determine both OR area and volume. For articles that described several ORs of different dimensions, the average area and volume are reported here. The near-linear relationship between area and volume shown by this graph supports our observation that, although ORs across hospitals vary significantly in their dimensions in terms of width and length, ceiling heights are fairly consistent at around 3 meters.

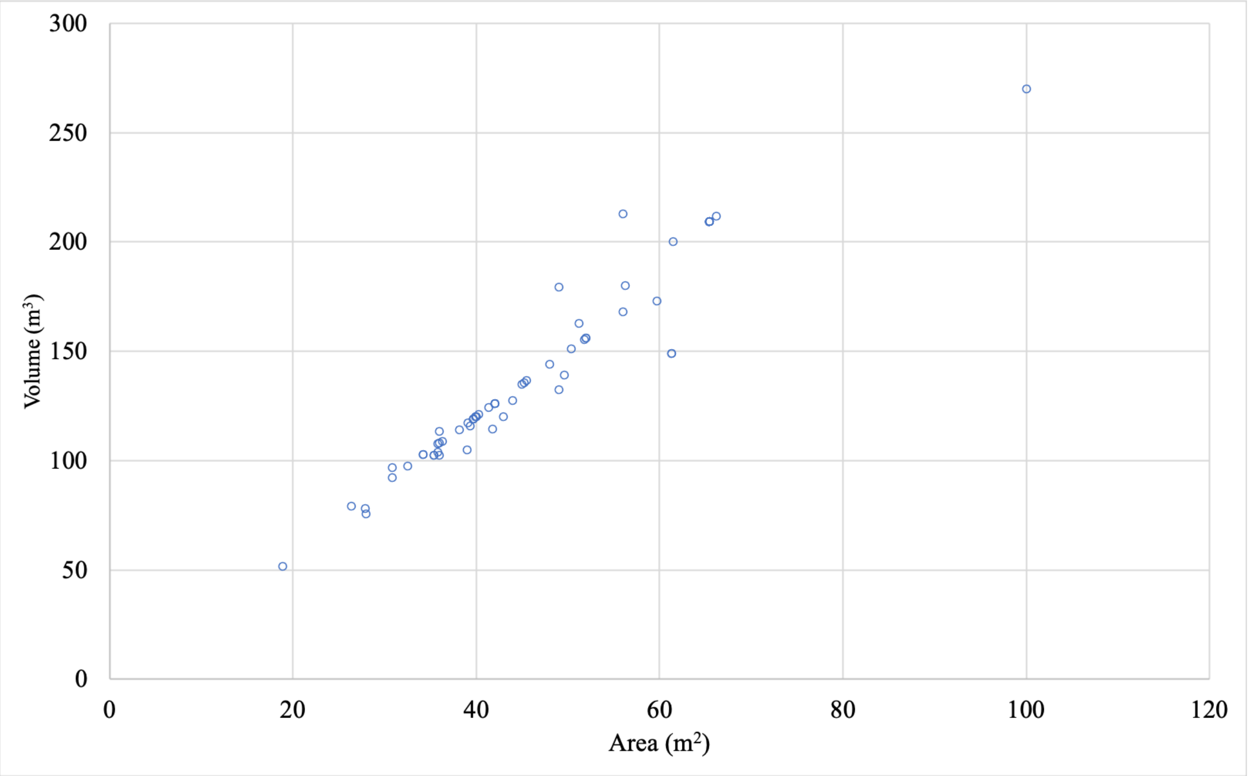

**Figure S2.** Plot of OR dimensions collected from the publications captured by this systematic review. The dimensions of real ORs and those used in modeling studies are both included.

**Error! Reference source not found.** and **Error! Reference source not found.** feature the OR areas and volumes, respectively, reported in the articles collected in this review. These graphs include data from articles that reported both area and volume, which were captured in **Error! Reference source not found.**, as well as data from those which only reported one measurement. Given this inconsistency in OR dimension reporting from the articles collected, the two figures below were created from different numbers of data points and described different overall sets of ORs. This may explain why the histogram for OR floor area is unimodal while that of OR volume is bimodal, an unexpected observation given that ceiling heights were found to be fairly consistent across studies. Overall, **Error! Reference source not found.** is perhaps the most informative of these supplementary figures, as it best demonstrates one of the only commonalities: OR ceiling height.

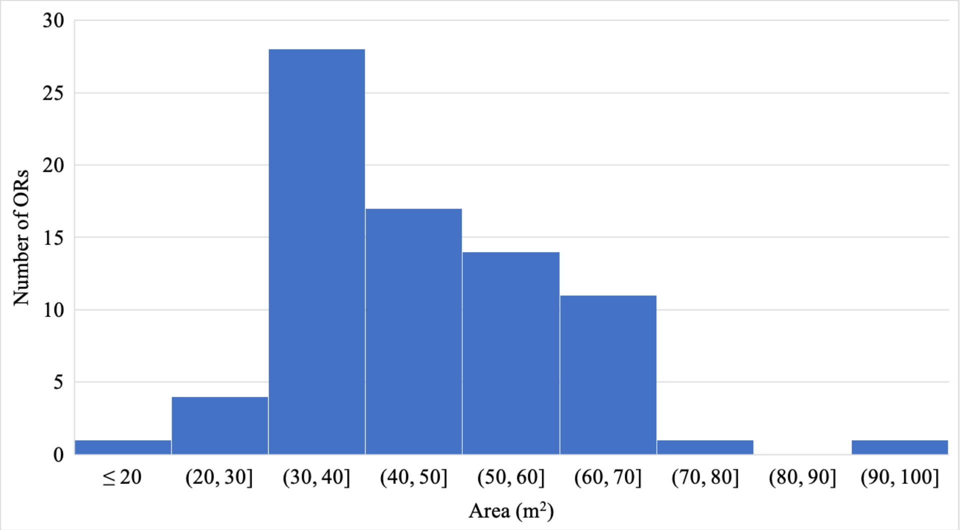

**Figure S3.** Distribution of OR floor areas collected from publications captured by this systematic review. The dimensions of real ORs and those used in modeling studies are both included.

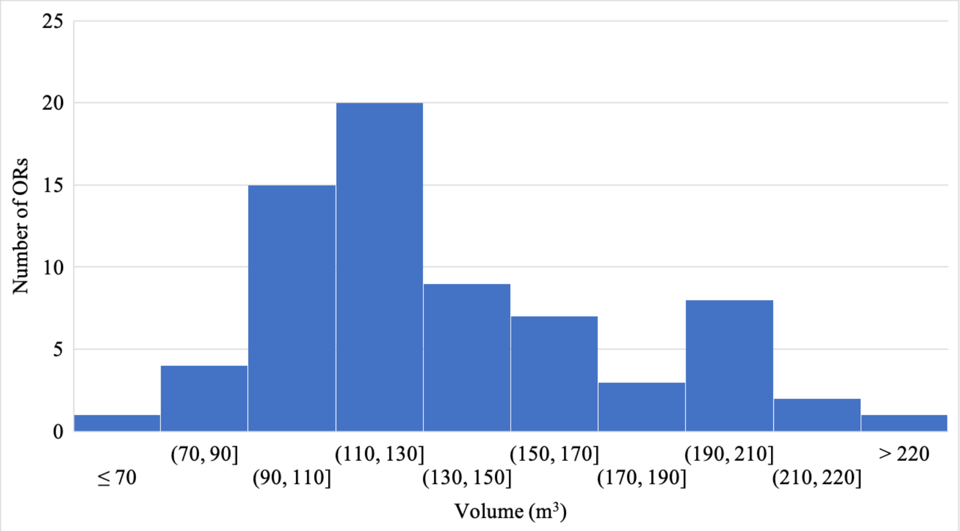

**Figure S4.** Distribution of OR volumes collected from the publications captured in this systematic review. The dimensions of real ORs and those used in modeling studies are both included.

### APPENDIX S3. LITERATURE REVIEWS AND META-ANALYSES

According to the fifth exclusion criteria used in this systematic literature review, "the article did not report any primary data," literature reviews and meta-analyses were excluded from the main body of this report. Given that these existing reviews help summarize the current literature surrounding the OR built environment, they have been included here as supplementary information. **Error! Reference source not found.** incudes a brief summary of each review article identified, separated by built environment component. These articles are further discussed in the pages that follow the table below.

**Table S1.** Literature reviews and meta-analyses not included in this systematic literature review.

| Reference | Study design | Location | Sample size | OR characteristic(s) | Aspect of built environment component | Outcome | Major Takeaway | Conclusion / Recommendation |
|---|---|---|---|---|---|---|---|---|
| **Disinfection systems** | | | | | | | | |
| 1 | Literature review | | | Arthroplasty surgeries | HEPA/UV system; UV-C light and air purification | Bacterial counts; infection rates | Particle count was reduced by 53.4% using the combined HEPA/UV system. The combination of UV-C light and air purification led infection rates per 1,000 patient days to drop from 17.5 to 12.5. | Bacteria count was reduced using a combination of HEPA filtration and UV light. While effective, more studies are recommended before using this system. |
| 2 | Systematic literature review | | | | Use of UV decontamination | Presence of infection-causing microorganisms | Application of UV lights in the OR is proven to effectively reduce airborne bacteria, although it is not currently advised due to potential risks associated with OR personnel. | Use of UV light for terminal cleaning of empty ORs may reduce risk of subsequent SSIs and prevent accidental staff exposure. |
| 3 | Systematic literature review | USA | | | Use of UVL/UVGI | SSI rates | Some previous studies suggest that UVL/UVGI systems lower SSI rates and may reduce CFUs. Conversely, other studies advise against the use of UVL/UVGI due to exposure risks to personnel. | Use of UVL/UVGI systems may provide reduction in surgical site infections, and if implemented, UV exposure should be closely monitored to protect surgical staff. |
| **Doors** | | | | | | | | |
| 4 | Literature review | UK | 29 studies of variable size | | Frequency and length of time for door openings | Air particle counts | There is a strong positive correlation between CFU/$m^3$ and number of door openings. Authors also note that results of different studies cannot be directly compared, as there is no standardized method for quantifying airborne bacterial contamination. | Behavioral changes including fewer door openings and less OR personnel traffic are recommended. Use of non-visible door counters for research purposes is preferred, as this reduces the Hawthorne effect. |

| Reference | Study design | Location | Sample size | OR characteristic(s) | Aspect of built environment component | Outcome | Major Takeaway | Conclusion / Recommendation |
|---|---|---|---|---|---|---|---|---|
| 5 | Literature review | | Variable | Variable | Frequency of door openings | Effect on positive pressure in the OR | One study found door-opening events caused loss of positive pressure in the OR in 77 out of 191 hip and knee arthroplasties. An experimental model of a mock OR and adjacent hallway demonstrated no air mixing between the positive-pressure OR and the lower-pressure adjacent space during door openings if the door was open for less than 15 seconds when positive pressure was at 2.5 Pa. The length of time the door could be open without contamination from the adjacent space was proportional to the area the door occupied and the magnitude of the pressure difference between the two spaces. A study of empty ORs similarly found that positive pressure was not defeated when a single door was open, even for up to 30 seconds, but simultaneous opening of >1 door defeated OR positive pressure and allowed ingress of contaminated air. | Association of OR door opening with the loss of positive pressure and increased bacterial CFUs is controversial. |
| 6 | Literature review | | Variable | Variable | Frequency of door openings | CFU | Door openings increased the number of CFUs by 69.3%. Every individual door opening was found to increase CFUs by 3%. | The study recommends reducing the number of times OR doors are opened during procedure. |
| 7 | Literature review | UK | 8 studies cited, ranging from 450 to 450,000 cases | | Frequency of door openings | Particle counts | There was a direct correlation between frequency of door openings and particle counts. | Door openings should be minimized once the surgical procedure has started. |
| 8 | Literature review | Singapore and USA | Andersson: 91 samples; not the only study cited | | Frequency of door openings | CFU/$m^3$ and SSI | There is significant evidence in the literature that frequent door openings have a negative effect on OR air quality, and are correlated with increased airborne bacterial contamination in multiple studies. Additionally, there is mounting evidence that frequent door openings are correlated with higher SSI rates, although this relationship is less well-established. | Given that door openings commonly occur for nonessential reasons, reducing them is a simple measure that has the potential to protect patients. |
| 9 | Literature review | France | A total of 27 articles were included in this review, of | Orthopedic surgeries rooms; general surgery rooms; n = 17 university | Frequency of door openings, length of time | Airborne particle and bacterial counts | Study results were variable, with average number of door openings per hour ranging from five to over | Given the inconsistency across studies, the results lack |

| Reference | Study design | Location | Sample size | OR characteristic(s) | Aspect of built environment component | Outcome | Major Takeaway | Conclusion / Recommendation |
|---|---|---|---|---|---|---|---|---|
| | | | which 11 were related to door openings | hospitals; n = 12 public hospitals; n = 2 private hospitals | | | 100. Door openings were positively correlated with airborne bacterial counts and correlates of infection risk. Multiple studies found that the reason for many or most door openings was either unknown or unnecessary. | clear evidence for recommendations. |
| **Ventilation systems** | | | | | | | | |
| 10 | Meta-analysis | Germany | 17 studies with approximately 500,000 cases total (8 studies on total hip arthroplasty, 6 on total knee arthroplasty, and 3 on abdominal and open vascular surgeries) | | Use of LAF ventilation in the OR | SSI rates (deep SSIs for arthroplasty cases and overall SSIs for abdominal/vascular cases) | Meta-analyses of the 17 cohort studies, grouped by type of surgical procedure, revealed no significant difference in the risk of developing an SSI. | Laminar airflow should not be regarded as a preventative measure against SSIs, and therefore should not be newly installed in ORs. |
| 11 | Literature review | United Kingdom | Four studies totaling around 100,000 cases | Specialty: joint replacement surgeries | Use of LAF ventilation system | Incidence rate of SSIs following TJA | Use of LAF does not seem to have a significant effect on SSI rate. | Given the extensive conflicting evidence surrounding the use of LAF, it is not appropriate to recommend this ventilation system for joint replacement surgeries. Further research is needed into the effect of heat sources including surgical lights on LAF. |
| 2 | Literature review | | | | Use of LAF during orthopedic surgery | Incidence of SSIs following orthopedic surgical procedures (namely PJIs) | Based on the studies reviewed, there is little agreement in the literature. Several early studies showed a benefit of LAF ventilation, but others conducted since have shown no benefit and a few have even found this type of ventilation to be harmful. | Given the large disagreement in the literature, hospitals should not spend the time and funds required to transition their orthopedic ORs to LAF without more extensive and conclusive evidence. |
| 12 | Literature review | Italy | 60 papers total; 17 papers related to ventilation | | Different ventilation systems | SSI rates | Some studies have shown that fewer infections arise when orthopedic surgery is performed in ORs with ultra-clean air ventilation systems (LAF systems). Two papers mention that improper positioning of OR personnel is more impactful on SSI increase than the type of ventilation. Overall, the current data does not agree on the existence of a | More studies must be conducted to determine if LAF is beneficial in reducing SSI rates and/or SSI risk. The prevention of SSIs requires a continued multidisciplinary approach, including infection surveillance and the coordination of those |

| Reference | Study design | Location | Sample size | OR characteristic(s) | Aspect of built environment component | Outcome | Major Takeaway | Conclusion / Recommendation |
|---|---|---|---|---|---|---|---|---|
| | | | | | | | correlation between type of ventilation and SSI rates. | involved in ventilation design and OR layout. |
| 13 | Literature review | Ireland | Unknown number of total studies reviewed, results included findings from 6 clinical studies/documents | | Use of LAF ventilation systems | Airborne bacterial contamination | The majority of laboratory-based studies assessing LAF systems conclude that there is decreased airborne bacterial contamination when LAF is in use, but only under the LAF ceiling or canopy. | The placement of surgical instruments is important and should be under the LAF canopy or, if necessary, placed outside of the canopy but should remain wrapped until used. The turbulent flow present outside the canopy can result in increased bacterial contamination rates on sterile trays. |
| 14 | Literature Review | Various Countries | 119 references | | Use of ultra-clean LAF | Risk of SSIs | While several studies, including a multi-center randomized control trial, have indicated that operating under LAF can reduce the risk of infection, multiple investigations conducted since have either showed no benefit or a negative association between this ventilation system and SSIs. The effectiveness of LAF is dependent on the ability of its air supply to travel undisturbed, so it is plausible that other interventions meant to further reduce SSIs interfere with its proper functioning. | LAF is not currently recommended by the World Health Organization and further studies into its placement and functioning are needed. |
| 15 | Literature review | United Kingdom | 62 surgery registry studies and randomized controlled trials (RCTs) | | UCV ORs versus CV ORs | Microbiological performance and infection rates | The reviewed studies show that UCV ORs have better microbiological performance than CV ORs. Because of the improved microbiological performance in UCV ORs, there is some reduction in infection rates in those ORs. But, if UCV ORs are used incorrectly and HCWs are not using proper techniques, infection can and does occur. | It is best to operate in a UCV OR over a CV OR, but since UCV constitutes a complex system, minor shortcoming in surgical technique can result in detrimental effects to air quality and therefore infection rates. |
| 8 | Literature review | | | | Type of ventilation system: LAF versus standard | Incidence of SSIs | LAF is not necessarily associated with decreased SSIs, given the contradictory information provided by the literature. | A large randomized controlled trial with SSIs as the end point is necessary before a firm conclusion can be made as to the role of LAF ventilation. In the meantime, national surveillance systems and joint registries should |

| Reference | Study design | Location | Sample size | OR characteristic(s) | Aspect of built environment component | Outcome | Major Takeaway | Conclusion / Recommendation |
|---|---|---|---|---|---|---|---|---|
| | | | | | | | | continue to monitor SSIs in order to identify correlations with type of OR ventilation. |
| 3 | Systematic literature review | USA | 48 papers are cited in the systematic review | | Use of LAF as described in the scientific literature published through June 2010 | Rates of PJI and some potentially associated variables, including bacteria measured in CFUs/$m^3$ and particle counts measured in particles/$m^3$ | Multiple early studies found that rates of PJI are reduced under LAF as compared to conventional HVAC systems, but improved patient care practices in recent decades may have eliminated this effect. The only randomized controlled trial identified found a reduction in rates of PJI under LAF, but also found that use of prophylactic antibiotics was a much more important factor. Other studies suggest that LAF is beneficial only when the OR conditions are carefully controlled, and in some scenarios can paradoxically be associated with increased infection rates. | A large, randomized, and standardized controlled study of PJI rates under LAF versus CV would be required to determine if LAF is beneficial and cost-effective. |
| 7 | Literature review | UK | | | Use of LAF in the OR | Risk of SSIs | For arthroplasty surgeries, only 2 out of the 8 studies statistically support use of LAF, while 5 out of the 8 do not, and one study finds contradictory results: a reduction in SSIs when LAF is used in hip replacements but not in knee replacements. For other types of surgeries, results are similarly inconsistent, and most studies fail to control for other infection risks and variables. | When using LAF, care should be taken to avoid disrupting the airflow with heat sources and physical barriers. |
| 16 | Systematic literature review | Studies in the meta-analysis came from Germany, New Zealand, the UK, and the US. | 20 papers published between January 2000 and September 2011 are cited; 5 papers were appropriate for meta-analysis | | Use of LAF in the OR | Risk of severe SSIs following hip and knee replacements | No study shows a benefit of LAF following knee prosthesis, and one study showed LAF significantly associated with higher SSI rates. One small study of hip prosthetics found LAF to be beneficial and three found it to be harmful. A meta-analysis using pooled data from the studies found increased relative risk of severe SSI following both knee prosthesis (RR 1.36) and hip prosthesis (RR 1.71). | LAF may be a risk factor for severe SSI, and the installation of LAF systems into new ORs should be discontinued. Further review is needed before determining whether existing LAF systems should be removed. |

| Reference | Study design | Location | Sample size | OR characteristic(s) | Aspect of built environment component | Outcome | Major Takeaway | Conclusion / Recommendation |
|---|---|---|---|---|---|---|---|---|
| 6 | Literature review | | 101 papers are cited | | Use of LAF or UCV in the OR during implant-based breast reconstruction | SSI rates | No evidence is found in the literature for the use of LAF or UCV in breast implant surgery, so studies of orthopedic implant surgery are reviewed. Effect on SSI rates is unclear, but the reduction in airborne and surgical field contamination with use of LAF is well-established. | Use of LAF or UCV is suggested. |
| 17 | Systematic literature review | USA | Papers published between January 1950 and April 2016 | | Functioning of the HVAC LAF system | Temperature, airflow, and SSI risk | High-velocity airflow can dilute and remove particles in the OR, but may also cause increased turbulence, humidity, and patient hypothermia, which is a risk factor for development of SSIs. | Changes to guidelines and ambient OR environments should be made with careful consideration of the safety of surgical patients and OR personnel. |
| 18 | Systematic literature review and meta-analysis | China | 14 studies: 10 studies on THA, 7 studies on TKA, and 3 studies on abdominal and open vascular surgery. 590,121 surgeries, 328,183 under LAF and 261,938 under CV | | Use of LAF versus CV in the OR | Risk of SSI | No significant differences in risk were found in the meta-analysis for any surgery type, suggesting that the type of ventilation may not have an independent effect on the risk of SSI. | No recommendation about the use of LAF can be made. |
| 5 | Literature review | USA | | | Use of LAF ventilation | SSI rates, air velocity, and particles in the operative site | Literature on the benefits of LAF is mixed: some studies show a reduction in SSIs and airborne CFUs, while others suggest LAF may be harmful or has no effect. Increasing the velocity of airflow above the recommended level of 25-50 feet per minute may in fact increase the number of particles directed toward the operative site. | LAF functions unpredictably when not used correctly. |
| 19 | Literature Review | USA | 211 articles | | Use of LAF ventilation | SSI rates | Ventilation system design and air quality are critical factors impacting patient and staff safety in the OR and many studies have focused on LAF ventilation systems with inconsistent impacts on bacterial counts and SSI. LAF ventilation systems have been shown to help reduce SSI in some studies while other studies showed no difference between LAF | Many of the studies that investigate aspects of the OR such as LAF are conducted by clinicians rather than OR design researchers, and the design implications of the findings are not always made clear. |

| Reference | Study design | Location | Sample size | OR characteristic(s) | Aspect of built environment component | Outcome | Major Takeaway | Conclusion / Recommendation |
|---|---|---|---|---|---|---|---|---|
| | | | | | | | systems and traditional ventilation systems. | |
| 20 | Systematic literature review and meta-analysis | Australia and the UK | 77,321 patients and 9 different treatment strategies | Orthopedic ORs | CV versus LAF | Risk of deep infection following THA | An infection prevention strategy that included CV significantly reduced the risk of SSI compared to a reference strategy. An otherwise identical strategy that was implemented had an SSI odds ratio of 1.96, indicating increased risk of SSI under LAF. | LAF may actually increase the risk of SSI following a THA, and should not be used. |

#### Disinfection systems

Overall, three literature reviews were identified that discussed the use of disinfection systems in ORs. The first of these reported on two disinfection systems.[1] In testing the first of these systems, bacterial counts were measured in an OR before and after using a system that combined both HEPA filtration and UV light for decontamination.[1] An air sampling impactor and agar media plates were placed around the OR to measure CFU per cubic meter of air. Biological contaminants passed through the C-UVC chamber where they are inactivated. These and other non-viable particles are then removed through the air exhaust HEPA filters. From the samples collected, there was a reported 53.4% reduction in overall CFU count from when the device was used to when it was not used.[5] The second system, which aims to destroy viable airborne contaminants by drawing air into a chamber equipped with UV-C light, was studied in terms of resultant infection rates. After the deactivation, the cleaned air is returned to the room through baffles. The use of this system decreased the number of infections per 1,000 patient days from 17.5 to 12.5, which is a statistically significant result.

The last two literature reviews discussed the use of UV light and germicidal irradiation systems to disinfect ORs.[2,3] Overall, the authors of both reviews concluded that:

- There is substantial evidence to suggest that UVL effectively reduces airborne bacteria in ORs
- Widespread installation is not yet advised due to the potential risks to OR personnel associated with prolonged exposure.

#### Operating room doors

A total of six literature reviews that investigated the influence of doors on the operating room environment were identified by this systematic literature review. Four of these only investigated frequency of door openings[5–8] and two investigated frequency and length of time of door openings.[4,9] Five of the six articles agreed on a positive correlation between door opening frequency and bacterial contamination in the operating room, all five of them having cited several empirical studies as evidence of this relationship.[4,6–9] Two of these further stated that door openings events are associated with higher incidents of SSIs in surgical patients.[8,9] The remaining review article concluded that the association between door openings and OR contamination is controversial, citing examples of both empirical and modeling studies, some of which failed to find a correlation.[5] Only two of the literature reviews explored the effect of door openings on ambient pressure inside an OR.[4,5] These reviews ultimately concluded that the relationship between loss of positive pressure due to door openings and increased CFU is controversial, as some empirical data from real surgeries showed positive pressure loss while some modeling studies showed no pressure loss during brief door openings.

[5] This review article provided specific data related to the use of C-UVC systems, but since this was not primary data, the article was excluded from the systematic review.

Overall, the literature reviews about OR doors collected in this review agree that door openings lead to increased contamination. Consequently, these reviews recommended a reduction in door opening frequency, mainly through training HCWs to be more cognizant of their movements while in an OR. Before these recommendations are implemented, however, additional research into the influence of door openings on the OR environment should be conducted. Of the aforementioned reviews, only one addressed the influence of door openings on OR pressure, suggesting that there is a gap in the current literature. While most articles agree that door openings lead to increased contamination, the research surrounding why this relationship exists has been limited. Without knowing how door openings increase contamination, recommendations as to whether HCW behavior or OR design should be modified are not well informed. Additionally, more research should be done to address whether door openings directly influence the incidence of SSIs. Finally, there is significant opportunity to investigate what danger, if any, door openings pose to OR personnel. The vast majority of the literature surrounding door openings in the OR discusses them in the context of patient safety, rarely addressing what impact may exist on HCWs. The aforementioned literature reviews are important summaries of the work that has already been conducted, but these also highlight the fact that there is still research to be done on the topic. Overall, the following observations can be made from these reviews:

- The existence of a positive correlation between door opening frequency and bacterial contamination in the OR is well-established but why it occurs is not well understood
- Only two review articles addressed the influence of door openings on OR pressure, and both concluded that the available information is inconclusive and warrants more study
- Since the mechanism through which doors openings increase OR contamination is not well understood, existing recommendations as to whether HCW behavior or OR design should be modified are not well informed.

#### Operating room ventilation system

Fourteen literature reviews and three meta-analyses were identified that investigated the influence of ventilation systems on airflow, contamination, and infection in the OR. Sixteen of these articles reviewed the literature surrounding the use of laminar airflow (LAF) ventilation in operating rooms,[2,3,14,16–20,5–8,10–13] while one article reviewed the literature for comparisons between ultra-clean ventilation (UCV) systems and conventional ventilation (CV).[15] It should be mentioned, however, that there is some inconsistency in the definitions of these types of ventilation. In one review article, LAF is used interchangeably with UCV,[12] whereas another explicitly compares and contrasts the two systems.[6] However, given that both of these articles discuss LAF, they were grouped accordingly. The literature review comparing UCV and CV systems investigated "microbiological performance" and infection rates as outcomes, ultimately finding that there is a small reduction in infection rates in ORs equipped with UCV.[15] Although "microbiological performance" was not defined in the review, the discussion surrounding it suggests that it refers to the ventilation system's ability to clear the OR air of biological contaminants. While the overall recommendation of this article is that UCV should be used over CV, the authors caution that UCV systems are complex and their misuse can result in detrimental effects to air quality in the operating room.[15] Of the 16 reviews that address the use of LAF, 15 include either surgical site infections (SSIs) or periprosthetic joint infections (PJIs) among the outcomes investigated.[2,3,16–20,5–8,10–12,14] Nine of these 15 literature reviews concluded that the currently available information on the ability of LAF to reduce SSIs provides inconsistent and contradictory results, thus preventing a definitive recommendation for or against its use.[2,3,5–8,12,14,19] Of the three meta-analyses that were collected, and two found that there was no significant difference in the risk of developing an SSI when surgeries were conducted in an LAF OR versus a CV OR.[10,18] The third meta-analysis found that the use of LAF might actually increase the risk of developing a SSI following THA, and ultimately recommended against the use of LAF, favoring CV instead.[20] Another literature review found that the use of laminar airflow does not have a significant effect on SSI rates in TJA cases.[11] Lastly, one literature review correlated temperature and velocity settings on LAF canopies with SSI rates. Ultimately, the authors found that while the high-velocity airflow provided by LAF can dilute and even remove particles that are generally thought to be harmful, such airflow can lead to increased turbulence, undesirable changes to air humidity, and decreased air temperature which can increase the incidence of hypothermia in patients.[17] The latter is a well-known risk factor for developing SSIs. The remaining article found that the majority of the reviewed laboratory-based studies on LAF concluded that LAF systems decrease airborne bacterial contamination but only in samples taken directly under LAF ceilings or canopies.[13] Overall, the following observations can be made from these review articles:

- Currently available information on the ability of LAF to reduce SSIs provides inconsistent and contradictory results, leading all currently available information to be inconclusive
- The only definitive conclusion reached by any review article on the topic is that type of ventilation system is not an independent risk factor for the development of SSIs.

### APPENDIX S4. SUMMARY OF INCLUDED ARTICLES

#### Operating room layout

**Table S2.** Summary of the two articles included that investigated the impact of the operating room layout on airflow, contamination, and infections.

| Reference | Study design | Location | Sample size | OR characteristic(s) | Aspect of Floor | Outcome | Major Takeaway | Conclusion / Recommendation |
|---|---|---|---|---|---|---|---|---|
| 21 | Empirical | Italy | 35 ORs in 30 hospitals | Orthopedic | Floor layout geometry (simple vs. complex).<br>Simple: regular geometries in plan and room height, symmetrical layout of the extraction points of the HVAC system. Complex: considerable geometrical complexity in the floorplan or room height, with the presence of beams, pillars, and cavities in various positions inside the room | TVC measured in CFU/$m^3$ | Floor layout geometry does not significantly affect microbiological air quality of the OR. | |
| 22 | Empirical | Netherlands | 829 surgeries, 4 ORs, 8 interviews | Eye surgery | Introducing floor markings.<br>Red tape (width 2.5 cm) was placed on the contours of the LAF area of the OR floor. Stop positions of the surgical tables were indicated by white tape dots | Cases of ophthalmic infections | There is evidence suggesting that floor markings aid personnel in being more responsible about making sure their devices are in the correct positions during the surgery. However, there is not enough conclusive data to assert that this leads to fewer ophthalmic infections (endophthalmitis). | OR floor markings are recommended to help guide where surgical devices need to be properly placed and may help reduce cognitive overload. |

#### Disinfection systems

**Table S3.** Summary of the several articles included that investigated the impact of disinfection systems on airflow, contaminations, and infections.

| Reference | Study design | Location | Sample size | OR characteristic(s) | Aspect of disinfection system | Outcome | Major Takeaway | Conclusion / Recommendation |
|---|---|---|---|---|---|---|---|---|
| 23 | Empirical | USA | 2 ORs, 3 air samples, and 14 surface samples pre | BICU | Use of PPX-UVD | Bacterial growth and mean colony count taken from surface and air microbial | PPX-UVD in a BICU reduced overall environmental bioburden, without a | The use of PPX-UVD has demonstrated reduction of environmental bioburden in BICUs; however, its effect on HAI and |

| Reference | Study design | Location | Sample size | OR characteristic(s) | Aspect of disinfection system | Outcome | Major Takeaway | Conclusion / Recommendation |
|---|---|---|---|---|---|---|---|---|
| | | | and post use of PPX-UVD | | | samples, HAI rates, and presence of MDROs or GNRs | statistically significant impact on HAI rates or MDRO, although no MDRO or GNR responsible for HAI were isolated from the environment, even before PPX-UVD. | MDRO is inconclusive and warrants further investigation. Therefore more data is required before advocating for the implementation of this portable cleaning device. |
| 24 | Simulation | Germany | 6 control and 6 plasma runs per configuration | | Use of CAP module | CFUs of various *E. coli* strains | Application of CAP plasma module as an air disinfectant effectively reduced CFUs for several strains of *E. coli.* Antimicrobial efficiency was improved with increased intensity and duration of treatment. | Based on the results of this study, use of CAP modules may improve air quality by inactivating strains of MDROs. However more research is required to improve antimicrobial efficiency and determine efficacy against viruses. |
| 25 | Empirical | China | 426 patients, including 213 controls | Neurosurgery | Use of UV lamp with minimum intensity of 70 $\mu W/cm^2$ | Bacterial strains, SSI rates, and microbial contamination measured in $CFU/m^3$ | Use of UV lamp with minimum 70 $\mu W/cm^2$ in a multi-step infection control pathway effectively reduced $CFU/cm^3$ in OR throughout surgery and ultimately resulted in significantly fewer SSIs in comparison to control group. | Based on the results of this study UV lamps of sufficient intensity may be implemented to assist in reduction of SSI rate in neurosurgery ORs, but warrants further exploration as an isolated variable. |
| 26 | Empirical | USA | 30 experiments | | Use of high energy UVL and distance of system from door | TPCs | UVL may be applied to significantly reduce TPCs within the OR, regardless of placement; however researchers suggest placing UVL closer to potential sources of contamination for maximum efficiency. | Use of UVL as an OR disinfectant should be considered, although personnel exposure was not considered in this study and requires additional analysis. |
| 27 | Empirical | USA | 3 ORs, 4518 surgeries | | Visible light CED system equipped with 405 nm LED visible light | CFUs and SSI rates | CED systems remove bacteria at a higher rate than manually cleaning an OR. SSI rates were reduced | Visible light CED systems enhanced disinfection beyond standard cleaning. |

| Reference | Study design | Location | Sample size | OR characteristic(s) | Aspect of disinfection system | Outcome | Major Takeaway | Conclusion / Recommendation |
|---|---|---|---|---|---|---|---|---|
| | | | | | | | significantly after installation, from 1.4% to 0.4%, but also decreased similarly in a control OR with no CED system. | |
| 28 | Empirical | USA | 1 OR, 50 surgeries | | UVL filtration system | Particle counts and viability | Particle counts decreased in both viability and total count; however, the reduction was not found to be significant. | Particle count was reduced using the UVL filtration system. |
| 29 | Empirical | USA | 496 procedures | | UVC air decontamination units | Postoperative infection rates | ORs with standard HEPA filter systems had more post-operative infections than ORs equipped with UV-C systems. | To reduce the risk of infection, enhancing of air purification of HVAC systems should be considered. |

### Operating room lights

**Table S4.** Summary of the fifteen articles included that investigated the impact of lights in the operating room on airflow, contamination, and infections.

| Reference | Study design | Location | Sample size | OR characteristic(s) | Aspect of lights | Outcome | Major Takeaway | Conclusion / Recommendation |
|---|---|---|---|---|---|---|---|---|
| 30 | Empirical | United States | 5 ORs | | Cleanliness on the surface of the lights | Presence of bacteria (*Streptococcus viridans, S. epidermidis, Staphylococcus epidermidis, Neisseria mucosa*) | Bacteria were isolated from surgical lights. This may be due to cross-contamination during cleaning, as the same wipes were used for multiple surfaces. | Prevent cross-contamination when disinfecting the built environment by using new materials for different components such as the lights, surfaces, and floors. |
| 31 | CFD modeling | Netherlands | | | Light/lamp shape and configurations | Airflow velocity and relative infection risk | Magnitude of mean peak airflow velocity did not change with different light shapes and configurations. Semi-open lights were related to the highest relative infection risk. | Surgical light/lamp shape and configuration may influence OR airflow and relative infection risk and should be incorporated into OR design planning. |

| Reference | Study design | Location | Sample size | OR characteristic(s) | Aspect of lights | Outcome | Major Takeaway | Conclusion / Recommendation |
|---|---|---|---|---|---|---|---|---|
| 32 | CFD modeling | Oman | Five model options with varying parameters that lead to 50 cases | | Type of light and light heat generation | Airflow and temperature impacted by lights | From a ventilation standpoint, a smaller size light with low generated heat may be the optimum selection. | Supply an UCV system capable of supplying at least 63 ACH and a surgical light with minimal face area to interrupt flow. |
| 33 | CFD modeling | Iran | | | Heat generation and placement of lights | Airflow patterns and particle deposition | Both the placement of the surgical lights and their thermal output can affect airflow patterns and particle deposition. The temperature of the lights is more important than their location. | Light placement and temperature should be carefully considered in OR design. |
| 34 | Empirical | Germany | | | Placement and position ing of lights | LAF | The position of lights in the OR affects the direction of airflow. | Smaller lights are potentially more desirable in the OR, and it may be important to position the lights out of the LAF if possible. |
| 35 | Empirical | United States | Two ORs, airflow was measured at 504 points each and cleanliness was measured at 8 points | | Placement and position ing of lights | Airflow, LAF | There was evidence of more uniform airflow in a room with lights in a double axis configuration and more variable airflow in a room with lights in a single axis configuration. In the single axis room, more rapid airflow was observed in the center of the room. In the double axis room, vertical LAF with uniform velocity was observed throughout. The double axis room also cleared out particles faster than the single axis room. | A double axis light configuration was suggested to be better for airflow and particle clearing in the OR as compared to a single axis light system. |
| 36 | Simulation | Netherlands | Five different light s ystems were examined under various circumstances. Each measurement was repeated 4 times | | Lamp shape configuration (open, semi-open, closed) | Particle concentration at the operating table | The different operating lamp shapes did not affect the particle concentration on the operating table in a significant way. The lamps were placed according to the VDI procedure (published particle dissemination test), which is behind the head of the surgeon. | The VDI 2167 particle test method to evaluate the ventilation system can be used in combination with infection incidence to better understand the influence of the ventilation system and infection risk. |
| 37 | Simulation | Norway | One subject in one OR. Total number of cases: 8 (heat | The floor was made of wood and raised from the standard concrete | Positioning of lights and light heat generation | Particles | Medical equipment significantly affected the $N_2O$ (ppm) levels in the sterile field, depending on the | Design equipment such that impact to UDF is minimized and/or design airflow systems that are not as readily affected by |

| Reference | Study design | Location | Sample size | OR characteristic(s) | Aspect of lights | Outcome | Major Takeaway | Conclusion / Recommendation |
|---|---|---|---|---|---|---|---|---|
| | | | load, C-arm, surgical lights); All measurements repeated twice with 18 successive samples for each case | industrial hall by use of wooden beams. The walls and ceiling were insulated on the outside. | | | location of the contaminant source and the amount of equipment positioned above the patient in the sterile field. The highest levels of the tracer gas concentration were recorded under the largest surgical light. | equipment positioned above the sterile field. |
| 38 | Simulation | Norway | 2 ORs, 2 scenarios with 4 different cases | HEPA-equipped UDF diffuser (3.4 x 3.4 $m^2$) in ceiling and enclosed by 0.5-m high partial glass walls | Placement and position ing of lights | Airflow, LAF | The effect of the lamps on airflow patterns is likely related to the heat generated by the lamps rather than the shape of the lamps. | LAF systems should be calibrated for all surgical facilities using simulated patients and surgical staff to ensure that air velocity fulfills the national standards for operating zone requirements. |
| 39 | Simulation | Norway | 2 ORs and 4 mock surgeries | Orthopedic ORs (~7-8 years old) | Placement and positioning of lights | Air velocity, bacteria measured in CFUs, and TI | Surgical lights significantly decrease air velocity. Slightly higher bacterial counts were observed when the lights were on versus off. The mean TI with lights was 3.05%. and the mean TI without lights was 2.92%. | Surgical lights should be positioned as far as possible from operating table but still close enough to illuminate the wound area. LEDs or methods to prevent heat dissipation may help with temperature control of the OR. Flatter shaped lights disrupt airflow more than rounder lights with the same surface area. |
| 40 | Simulation | Norway | | Orthopedic OR | Placement and positioning of lights | Airflow velocity | The airflow velocity was lower above the operating table when the surgical lamps were placed directly above the operating table. | Lateral airflow systems should be calibrated with all surgical facilities with the simulated patient and surgical staff to ensure that the air velocity fulfills the requirements in the operating zone. |
| 41 | Simulation | Netherlands | 1 OR, 264 samples | | Comparing lights with and without skirts | Degree of protection (DP), which is defined as the logarithm of the quotient (ratio) of the number of particles with a size of 0.5 µm or larger in the protected area compared to the number of particles of this size in the periphery. | A skirt can have a positive effect on the DP at the center of the protected area and seems to have a minor effect on the size of the protected area. Solid Y-shaped lamps demonstrated a more negative effect on the DP at the center of the protected area, potentially due to its semi-open shape. | Suppliers need to design lights to minimize interference with room airflow. Skirts on lights, if at proper height, help to protect the patient. A hub and spoke light with smaller surface area was better for maintaining airflow. |

| Reference | Study design | Location | Sample size | OR characteristic(s) | Aspect of lights | Outcome | Major Takeaway | Conclusion / Recommendation |
|---|---|---|---|---|---|---|---|---|
| 42 | CFD modeling | China | | | LED surgical light versus typical lamp type | Turbulent airflow eddies | Fewer eddies formed near the surgical OR table when using LEDs as compared to using typical lamps. | LED-based surgical lamps may offer a more sterile environment in the OR and reduce turbulent airflow. |
| 43 | Simulation | United Kingdom | Five different lighting configurations were assessed: no lights (at the extreme of the canopy); one light directly above the surgical field; two lights touching, directly above the surgical field; two lights 50 cm apart; two lights 160 cm apart. Each experiment was repeated five times | Experiment was designed to represent the typical setup for a TKA | Placement and position of lights | LAF disruption | The use of surgical lights had a significantly disruptive effect on LAF, potentially substantially reducing its effectiveness. | Flow disruption can be mitigated in part by repositioning the lights into a more favorable position for LAF. In order to maximize the effect of LAF, lights should be as far from the operating field as practicable. If possible, avoid suspended lighting within the LAF hood altogether. |
| 44 | Modeling | Sweden | Using one OR's dimensions, 2 types of ventilation were modeled (mixing and UDF), and 2 lamp orientations tested | | Lamp shapes, Innovative fan-mounted versus conventional closed-shape | Contamination near the wound | Innovative fan-mounted surgical lamp designs helped to reduce the contamination near the wound as compared to the conventional closed-shape lamp. | Mounting a fan in a surgical lamp can help eliminate low-velocity stagnant areas, which can help reduce contamination during surgery. |

#### Operating room doors

**Table S5.** Summary of the 33 articles included that investigated the impact of doors in the operating room on airflow, contamination, and infections.

| Reference | Study design | Location | Sample size | OR characteristic(s) | Aspect of doors | Outcome | Major Takeaway | Conclusion / Recommendation |
|---|---|---|---|---|---|---|---|---|
| 45 | Empirical | Italy | 14 hospitals; 28 OTs: 16 vertical unidirectional, 6 turbulent, 6 mixed airflow ventilation; 1228 elective procedures | Volumes varied from 90 $m^3$ to 180 $m^3$ (mean 116 $m^3$, with SD 20.4). HVAC systems were equipped with HEPA filters with efficiency >99.97% for particles size >0.3mm.; mean number of air changes per hour was 18 (SD 4.5) | Frequency of door openings | Microbial air contamination values | There is positive correlation between microbial air contamination values and the number of door openings. The number of door openings in an OT during surgical activity can be regarded as a key factor in increasing bacterial counts. | One cannot assume that a UDF system will always provide acceptable bacterial counts, even when engineered and monitored properly, and functioning correctly. It is essential to increase HCWs' awareness. |
| 46 | Empirical | Sweden | 6 ORs total, 3 with LAF, 3 with displacement ventilation (DV). 30 surgeries in DV ORs and 33 surgeries in LAF ORs are included | Orthopedic, LAF room areas were 46 $m^2$, while DV rooms were 39 $m^2$. Temperature was 20ºC (plus or minus 1ºC in LAF, 2ºC in DV). Airflow: 9,160 $m^3$/h in LAF, 2,430 $m^3$/h in DV; air velocity: 0.25 m/sec in LAF (SD 0.02) and 0.09 to 0.15 m/sec in DV | Frequency of door openings | Contamination measured as CFU/$m^3$ | In rooms with DV, every door opening instance increases CFUs by 3%, while no significant change is observed in the rooms with LAF systems. | The technical ventilation solutions are important, but they do not guarantee clean air. The organization of work and the behavior of the staff will influence outcomes. Continuous maintenance and risk assessments of LAF and other technical systems are crucial. |
| 47 | Empirical | Pakistan | Two hospitals, three ORs at each hospital, monitored for 24 hours | GOT: general operation theatre; OOT: orthopedic operation theatre; EOT: emergency operation theatre<br>Two of each OR were included; OOT2 has LAF<br><br>Floor areas, all in $m^2$: GOT1, 29.42 $m^2$; GOT2, 29.09 $m^2$; OOT1, 30.73 $m^2$; OOT2, 30.56 $m^2$; EOT1, 39.03 $m^2$; EOT2, 25.46 $m^2$ | Door openings | PM | There was a comparative reduction in the PM during surgery hours with the door closed as compared to an open door. Additionally, higher concentrations of PM were observed during non-activity periods with open door scenarios. | Open doors cause maximum infiltration from outside. |
| 48 | Empirical | Taiwan | Four class 10,000 ORs (defined to have a maximum concentration of 10,000 particles/$ft^3$ for PM≥0.5), two trauma ORs and two colorectal ORs, from a medical | Each OR has dimensions of 7.68 m (length) x 5.19 m (width) x 3.0 m (height) for a total volume of 120 $m^3$ | Frequency of door openings | TVOC concentrations | Increasing the door opening time was found to increase both TVOC and PM 10 concentrations. In trauma ORs, door openings were positively correlated with both TVOC and PM 2.5 concentrations. | The door opening time impacts the suspended particle concentration. Door opening time and number of staff in the OR should be monitored in order to maintain optimal air quality. |

| Reference | Study design | Location | Sample size | OR characteristic(s) | Aspect of doors | Outcome | Major Takeaway | Conclusion / Recommendation |
|---|---|---|---|---|---|---|---|---|
| | | | center in Northern Taiwan. Air sampling lasted 10 h in each OR at each ventilation rate and the sampling was triplicated during the study period | | | | | |
| 49 | Empirical | Egypt | 8 ORs in one hospital were studied, initially with two hour-long samples per OR, then repeated two weeks later | Temperature averaged 24.48ºC (SD 1.39ºC); humidity averaged 58.31 % (SD 6.54%) | | PM 2.5 | PM 2.5 had a statistically significant relationship with the status of the door (open vs. closed). | |
| 50 | Simulation | USA | 1 OR with and without LAF turned on | OR in an arthroplasty unit equipped with vertical LAF system with HEPA filters and two 12 in x 18 in exhaust grills. Area is 534 $ft^2$ and height is 9.2 ft | Frequency of door openings and length of time door is opened | Aerosolized particles | A positive trend was observed between the number of door openings and the amount of aerosolized particles, but did not reach statistical significance. | |
| 51 | Empirical | UK | 2 ORs, 46 consecutive operations | 2 cardiac ORs at Leeds General Infirmary | Frequency of door openings and length of time door is opened | Contamination and SSIs | Door opening disturbs airflow and results in increased air and wound contamination. The sample size was too small to statistically link the door openings to SSIs. | Education and training combined with robust audit processes are most likely to improve current practices. |
| 52 | Empirical | Switzerland | 2 ORs, 688 patients | LAF ORs at University Hospital Basel. "Internal door" led to clean instrument prep rooms attached to ORs; "external door" led to outside the operating area | Frequency of door openings and type of door being opened | SSI risk | Increased mean door opening frequency was associated with SSI risk. Further analysis shows that the increased risk for SSIs was driven by internal door openings and that external door openings had no correlation with SSI risk. | Better coordination in the instrument prep room beforehand could reduce the number of internal door openings and therefore SSI risk. |
| 53 | Empirical | Sweden | 3 ORs, 30 orthopedic surgeries, 116 air samples collected | ORs are 39 $m^2$ with a single door opening inwards. Pressure difference is 3 kPa. Upward air-displacement system supplies cool air (2-3ºC below room temp) above the floor in each of the 4 corners of the room. By thermal convection, air is evacuated via four exhaust fans in the ceiling | Frequency of door openings | $CFU/m^3$ | High rate of door openings was associated with higher $CFU/m^3$. | Better logistics and planning including enhanced knowledge would give OR staff the necessary tools to minimize door openings. |
| 54 | Empirical | Taiwan | 28 ORs, 250 surgical procedures | | Frequency of door openings | $CFU/m^3$ | Found a significant positive correlation between the | Doors should be kept closed in surgical |

| Reference | Study design | Location | Sample size | OR characteristic(s) | Aspect of doors | Outcome | Major Takeaway | Conclusion / Recommendation |
|---|---|---|---|---|---|---|---|---|
| | | | | | | | frequency door opening and bacterial counts. | procedures except as needed. ORs doing complex surgeries with more surgical personnel present should increase the frequency of air exchanges. |
| 55 | CFD modeling | Sweden | 1 OR | OR modeled is 8.6 m x 7.7 m x 3.2 m. Temperature in OR is modeled at 20ºC and the corridor is modeled at 18ºC in winter and 21ºC in summer. Turbulent airflow of 2.0 $m^3/s$. Positive pressure of 5 Pa relative to corridor | Frequency of door openings | BCP concentration before, during, and after the door is closed and opened | A single door cycle increases the overall airborne BCP concentration in the OR by approximately 2.1 $CFU/m^3$. The maximum contamination level found in the OR was similar under the summer and winter conditions. Contamination decreases following door closure, and after 5 minutes drops to about 0.1 $CFU/m^3$, or about 5% percent of the peak level of contamination. | Decreasing the exhaust rate while the door is open can reduce the influx of contaminated air from the corridor. |
| 56 | Empirical | USA | 21 procedures in 4 ORs (12 orthopedic procedures in one OR, 9 pediatric procedures in 3 ORs), two seasons (March and September) | | Number of door openings per procedure | $CFU/m^3$ in air | The number of door openings did not significantly impact microbial load, regardless of the location of sampling or the season. | |
| 57 | Empirical | Ghana | 3 ORs, 124 cases | Floor area is 36 $m^2$ | Frequency of door openings | Contamination measured as $CFU/m^3$ | There was a significant positive correlation between the frequency of door openings and OR contamination in $CFU/m^3$. | Many door openings were categorized as ‘unnecessary door-openings,’ suggesting that the door opening rate could be reduced without negatively impacting surgical performance. |
| 58 | Empirical | USA | August and October of 2011, 81 orthopedic cases, mainly TJA in 3 ORs. Total of 642 samples | | Frequency of door openings | Number of contaminated agar plates | The number of contaminated plates increases by almost 70% if any door opening event occurs in the OR. | Installing door locks, educating the staff, and having all the required equipment inside the OR at the time of incision are all interventions that could theoretically decrease the potential for infection by decreasing the number of door openings. |

| Reference | Study design | Location | Sample size | OR characteristic(s) | Aspect of doors | Outcome | Major Takeaway | Conclusion / Recommendation |
|---|---|---|---|---|---|---|---|---|
| 59 | Empirical | USA | 36 total hip arthroplasty cases, 2 ORs | | Frequency of door openings | $CFU/m^3$ at the surgical site | The number of door openings was not significantly related to $CFU/m^3$ at the surgical site. | |
| 60 | CFD modeling | USA | One simulated OR, number of trials not specified | Dimensions: 8.5 m × 7.7 m × 3.2 m. The OR was maintained under a positive pressure of 15 Pa relative to the adjacent corridor. The inlet air was introduced to the room at a total airflow rate of 2.5 $m^3/s$ and a temperature of 20ºC. The ceiling diffusers were recessed from the false ceiling and covered a projected area of 8.9 $m^2$. The temperature of the adjacent corridor air leading to the OR during door-opening was 24ºC | Type of door (both sliding and hinged), length of time door is open (16 s total: 3 s to open, 10 s open, and 3 s to close) | Airborne contamination ($CFU/m^3$) | Door openings can allow contaminated air to enter the OR, increasing the volumetric CFU concentration. When door openings were modeled to occur every 2.5 minutes, contaminant levels were observed to increase by about 7 $CFU/m^3$. | Door openings are a contamination risk and should be avoided. More research is needed to understand the relationship between contamination and SSI risk. |
| 61 | Simulation | Spain | Three types of tests were carried out: a) door opening-closing, b) door opening, a person entering the OR and the door closing, and c) door opening and a person leaving the OR and door closing.18 ORs at the University of Valladolid Hospital | The OR temperature was 22ºC with a positive pressure of 20 Pa. The OR was supplied with 18 ACH. Return air grilles near the ground measured 38 cm x 10.5 cm and 18 cm x 10.5 cm near the ceiling. Sliding door measures L=1.48 m and H = 2.10 m. Full door opening time is 14 s with a door motion speed is 0.27 m/s | Frequency of door openings and closing, door opening and closing with foot traffic | Airflow | Results show that, even with a sliding door and an initial overpressure of 20 Pa, a small volume of air enters the OR with every door opening, even without personnel traffic. Furthermore, if a person walks through the door the volume of air entering the OR is higher, especially if the person enters the OR. | |
| 62 | CFD modeling | Italy | The study investigated 3 case scenarios: A) opening and closing of the sliding door; B) opening of the sliding door, one person crossing, closing of the sliding door; C) opening of the sliding door, two people with a stretcher crossing, closing of the sliding door | Floor dimensions are 6.3 m x 6.3 m with a total volume of 119.07 $m^3$. The OR was modeled with typical a standard ISO 5 class layout with ultra clean air filters system. Internal air temperature of the OT was modeled as 20ºC and that of the surrounding zones as 26ºC | Opening and closing of sliding door | Airflow and velocity; pressure | The results obtained from all three cases indicate a strong modification of the air velocity field inside the OT due to opening/closing of the door and staff movements. The fluctuation of positive pressure inside the modelled OR during door openings was evaluated, highlighting that pressure reaches values very close to the lower threshold limits recommended by international standards. | |
| 63 | Empirical | USA | 7 cases in one OR | Building built in 1997, OR used for plastic surgery/reconstruction | Frequency of door openings | Air particle counts and particle size | Overall air particulate counts of all sizes increased by 13% | Make use of intercom or other technologies that allow HCWs within the |

| Reference | Study design | Location | Sample size | OR characteristic(s) | Aspect of doors | Outcome | Major Takeaway | Conclusion / Recommendation |
|---|---|---|---|---|---|---|---|---|
| | | | | | | when doors are open versus closed | when at least one door to the OR was open. | OR to communicate with those outside in order to reduce door openings. |
| 64 | Empirical | USA | 191 cases | | Frequency of door openings from cut to close, length of time the door is open | Pressure | For 77 of the 191 cases, the doors were open long enough for positive room pressure to be defeated, causing air to flow into the OR. Total door-open time significantly affected the minimum pressure recorded in the room, but did not significantly affect the average room pressure, suggesting that the loss of positive pressure was a transient event from which the room recovered. | |
| 65 | Empirical | USA | 48 orthopedic and general surgery procedures | Temperature kept at 65ºF. OR was equipped with a vertical LAF system that uses a combination of outside and return air through a HEPA filter that is replaced every 3 years | Frequency of door openings; length of time door is open | CFUs (no pathogen specified) | There was a statistically significant positive correlation between door openings and overall CFUs, as well as for the subset of cultures taken outside of LAF. The relationship was not statistically significant with cultures taken within the LAF. | In ORs not equipped with LAF, door openings should be limited as much as possible. |
| 66 | Empirical | France | ORs (n = 13); hospitals (n = 10); Procedures (n = 59); Orthopedic procedures (n = 34); Cardiac procedures (n = 25) | 2 specialties: cardiac and orthopedic. Some located in university hospitals, some located in private hospitals | Frequency of door openings | Particle count, wound culture, microbial air counts, CFUs | There was a significant positive correlation between particle counts and the number of door openings per 5-minute period. An increased number of door openings per 5-minute period was also associated with increased air microbial count. | The authors concluded that the intraoperative discipline of staff is important and they suggested that restricting staff movements and door openings in the OR would potentially prevent airborne contamination and therefore the associated SSI risk. |
| 67 | Empirical | Ghana | 358 patients in 7 ORs | Temperature varied between 15-25ºC | Number of door openings per operation | SSI rates | Operations with greater than 100 door openings more than doubled the risk of developing an SSI. Researchers found 25 patients developed SSIs in 234 procedures with less than 100 door openings and 33 patients developed SSIs in 124 procedures with greater than 100 door openings. | Unnecessary social visits as well as staff walking in and out of the OR without a clear purpose during an operation should be actively discouraged. Proper planning by staff and a central logistic distributing area within |

| Reference | Study design | Location | Sample size | OR characteristic(s) | Aspect of doors | Outcome | Major Takeaway | Conclusion / Recommendation |
|---|---|---|---|---|---|---|---|---|
| | | | | | | | | the theatre complex may reduce the need for staff to enter functioning ORs for logistics for other ongoing operations. |
| 68 | Simulation | USA | 5 trials per day, 2 days in phase 1 and 2 days in phase 2 (20 trials overall, 10 in each phase). The two phases were conducted in different hospitals and in different seasons | | 1) Frequency of door openings, 2) door size width of opening (45 vs 90 degrees), and 3) length of time door is open (6 vs 12 secs) | CFUs on settle plates, fungi and bacteria (based on type of agar used) | 1) no significant impact on fungi or bacteria; 2) wider door opening led to increased total CFUs (bacteria and fungi combined); 3) longer door openings led to a significant increase in number of bacterial CFUs and combined fungi plus bacteria, but not for solely fungi. | |
| 69 | Empirical | Netherlands | 70 procedures total in 2 ORs. 1 OR in Nijmegen with 59 procedures, and 1 OR in Delft with 11 procedures | In Nijmegen, the OR is under positive pressure in relation to the adjacent rooms (5 Pa) and air temperature and humidity are set at 18°C and 56%–60%, respectively. In Delft the OR is under positive pressure in relation to the adjacent rooms (15 Pa) and air temperature is set at 18°C and humidity at 50%–65%. Both were orthopedic ORs equipped with TV systems with HEPA filters | Frequency of door openings, reason for openings when available | Number of CFUs | After adjusting for the number of people in the OR and duration of the procedure, the number of door openings per surgery was associated with increased CFU/m$^3$. Additionally, every door opening increased the odds of unacceptable CFU/m$^3$ values by 5 %, with "unacceptable" being defined as > 20 CFU/m$^3$. | |
| 70 | Empirical | USA | 30 orthopedic cases (fractures) | | Frequency of door openings | Amount of time elapsed prior to contamination at the C-arm with aerobic and anaerobic bacteria | Time to C-arm contamination was not significantly related to number of door openings. The most common bacteria identified were *Staphylococcus* (62%), *Corynebacterium* spp. (31%), or *Micrococcus* spp. (5%). | |
| 71 | Modeling | Italy | 1 OR modeled (empirical data used to validate model) | OR was modeled with a volume of 120m$^3$ and sliding doors measuring 2.2 m x 1.4 m | Door opening event | Pressure and airflow | A total volume of 16.3 m$^3$ of OR air outflows toward the corridor during the door opening, person entering, and door closing event (around 11 seconds). A significant drop in pressure relative to the corridor is observed when the door opens, with the original +32.6 Pa differential dropping to +1.2 Pa. | |

| Reference | Study design | Location | Sample size | OR characteristic(s) | Aspect of doors | Outcome | Major Takeaway | Conclusion / Recommendation |
|---|---|---|---|---|---|---|---|---|
| 72 | Simulation | USA | 6 ORs within one hospital but at 2 locations. Each OR was equipped with a main door and a door connected to a substerile hallway | ORs were used for TJA. Temperatures varied from 61-67°F and relative humidity varied from 20-33% | Door opening event | Recovery time following depressurization | The average time to room recovery from a depressurization event was 14.22 seconds for personnel entering through the main door, 14.11 seconds for passing equipment through the main door, and 14.93 seconds for personnel entering through the inner door. Notably, the opening of the main door generated a negative pressure gradient at the closed inner substerile door between the pressure of the OR and the pressure of the inner substerile hallway. This finding suggests that if the inner door were to be opened while the main door was open, contaminated air from the inner substerile hallway would enter the OR. | OR door usage should be limited to a single door, both during room preparation activities and during the procedure itself. The entry of nonessential personnel during the surgical procedure should continue to be discouraged. |
| 73 | Modeling | China | Two sets of temperature conditions (lower in the OR versus lower in the anteroom), 5 cases each which vary both the ambient temperature of the OR and the temperature difference between the two rooms | The dimensions of the OR and the anteroom are 7.80 m (L) × 5.80 m (W) × 3.00 m (H) and 3.00 m (L) × 6.83 m (W) × 2.60 m (H), respectively. The air gap around the door was modeled as 0.006 m and +6 Pa can be maintained between the OR and the anteroom. The particle quantity generated by the floor and the surfaces of enclosure structure were modelled as $1.25 \times 10^4$ CFU/(min·m$^2$). The total quantity of particles generated by the anteroom environment is 18,100 CFU/s, modelled based on one person in activity | Door opening | Airflow, particles, air infiltration rate | Higher airflow velocity occurs when the sliding door opens and closes. Contaminants begin to accumulate in the upper part of the OR when the door undergoes the process of opening and closing. Air infiltration rate and air volume both peak right around the fully opened and closing phases of the sliding door. The rate also increases when the temperature difference between the OR and adjacent room is higher. | Temperature difference between the inside and outside of the OR should be minimal to prevent any large accumulation of contaminants in the OR while the door is opened and subsequently closed. |
| 25 | Empirical | China | 426 patients (213 per group - control and experimental) | LAF-equipped | Door openings: as part of an infection control protocol including multiple interventions, | Bacterial contamination as CFU/cm$^3$, SSI rates | The experimental group had significantly better air quality at all time points in terms of CFU/cm$^3$, and had significantly fewer SSIs than the control group. | |

| Reference | Study design | Location | Sample size | OR characteristic(s) | Aspect of doors | Outcome | Major Takeaway | Conclusion / Recommendation |
|---|---|---|---|---|---|---|---|---|
| | | | | | doors were kept closed throughout surgery for experimental group | | | |
| 26 | Simulation | USA | 30 experiments | | Occurrence of door openings (4 door openings over 20-30 min) | TPC and VPC | Largest increases in TPC and VPC occurred approximately 90 seconds after each walkthrough event. The C-UVC device significantly reduced TPC when placed 8 or 4 m from the opened door, and also significantly reduced VPC at 4 m. | Placing a C-UVC unit near the OR door can help mitigate the bacterial contamination introduced by OR traffic. |
| 74 | Empirical | Sweden | Three ORs (different ventilation systems LAF, TMA, TcAF), 15 cases in each OR (same exact number and type of procedures in each OR) | Orthopedic ORs | Frequency of door openings (2.1-5.6 openings per surgery per hour) | CFUs at the surgical wound | Door openings had no significant correlation with the number of CFUs found at the wound. | |
| 75 | CFD modeling | UK | 24 different cases where there is (1) excess or equal pressure (n=2), (2) various inlet velocities (n=3), (3) door inflow velocities (n=4) | OR model dimensions 6.4 m x 5.6 m x 2.9 m with a set of double-hinged doors measuring 1.6 m x 2 m. Operating table centered under the main air inlet and enclosed by a clean zone measuring 3m x 3m x 2.9m. Temperature 25°C. Pressure set to 0 and then to 20 Pa. Unidirectional filtered air 0.3–0.4 m/s with inlet air velocities of 0.2m/s, 0.3m/s, and 0.4 m/s and turbulent intensities from 8% -12% | Frequency of door openings | Door inflow velocities | Positive pressure reduces the risk of contaminated air entering the OR via door openings, especially in conjunction with the use of UCV. | ORs should be designed to function despite the challenge of door openings, through the use of positive pressure, an ultra-clean lobby, and by establishing the clean zone sufficiently far away from the door(s). |

### Operating room ventilation

**Table S6.** Summary of the 86 articles included that investigated the impact of ventilation in the operating room on airflow, contamination, and infections.

| Reference | Study design | Location | Sample size | OR characteristic(s) | Aspect of HVAC | Outcome | Major Takeaway | Conclusion / Recommendation |
|---|---|---|---|---|---|---|---|---|
| 76 | Empirical | Germany | 6 ORs, 277 surgeries | | Types of ventilation: window-based, supported air nozzle canopy, low-turbulence displacement airflow supply air canopy, or low-turbulence displacement airflow supply air canopy with flow stabilizer | Bacterial contamination, measured as CFU counts collected on settle plates | Highest bacterial burden was in the window-based ventilation OR, and lowest in the OR with the low-turbulence displacement airflow with a flow stabilizer. | An OR with a low-turbulence displacement airflow system with a flow stabilizer was the most effective ventilation system in reducing intraoperative bacterial burden. |
| 77 | CFD modeling | Malaysia | 1 OR | Specialty: Cardiovascular surgeries | Overall and relative velocity of air supplied from the HVAC inlets in a single OR case study | Temperature, humidity, and concentration of gaseous contaminant ($CO_2$) | The rate of airflow can be safely reduced by 15%, to an average of 0.3 m/s, without allowing humidity to exceed 60% or temperature to exceed 20°C. Uneven rates of airflow from the HVAC inlets are more effective than uniform rates in removing gaseous contaminants from the surgical area. | It may not be necessary to maximize airflow velocity, depending on the layout of the HVAC inlets. |
| 78 | CFD modeling | Malaysia | Modeled according to 1 OR | OR with non-standardized HVAC setup and internal storage area | Comparing inlet velocities of 0.2 m/s and 0.4 m/s | Contamination modeled with $CO_2$ gas | The concentration profile for the contaminant was virtually identical between the two inlet velocities. | Doubling the inlet velocity was not successful in compensating for the shortcomings of this OR design. |
| 79 | CFD modeling | Sweden | 1 model | The OR was modeled with dimensions 8.5 m (L) x 7.7 m (W) x 3.2 m (H) and a temperature of 20°C. Ventilating air is introduced through 24 diffusers placed evenly over the ceiling (downflow) or on a side wall (lateral-flow, cases 1 and 2) | Direction of airflow (horizontal LAF versus vertical LAF) and variation in air renewal rate measured in ACH | Particle counts | Horizontal LAF with an increased air renewal rate proved to be the best at reducing particle counts as compared to the other scenarios tested. Vertical LAF with an increased air renewal rate resulted in reduced particle counts, however this was not as good as that provided by horizontal LAF. | Horizontal LAF is a better choice than vertical LAF in an OR, but it must not be obstructed and needs to maintain the correct air renewal rate. |
| 80 | Empirical | France | 10 healthcare facilities; 13 ORs (orthopedic and | The OR dimensions are as follows: Orthopedic | Use of LAF versus TV systems | Particle count and size; air microbiologic sampling; wound culture | TV systems were associated with significantly increased air microbial contamination in | The authors recommend continued surveillance of proper functioning of OR |

| Reference | Study design | Location | Sample size | OR characteristic(s) | Aspect of HVAC | Outcome | Major Takeaway | Conclusion / Recommendation |
|---|---|---|---|---|---|---|---|---|
| | | | cardiac specialties); 60 procedures studied | ORs had an average volume of 105 $m^3$ (range 102-136 $m^3$). Cardiac ORs had an average volume of 124 $m^3$ (range 10 1-134 $m^3$). Orthopedic ORs had an average surface area of 39.4 $m^2$ (range 34 -46 $m^2$) while cardiac ORs had an average surface area of 39 $m^2$ (range 36-46.9$m^2$). All ORs were kept at positive pressure, with orthopedic ORs at 20 Pa (range 19-42 Pa) and cardiac ORs at 19 Pa (range 12-33 Pa) | | | comparison with LAF systems. There was no statistically significant relationship between air microbial contamination and wound contamination at closing. | air ventilation systems to ensure SSI prevention and that more empirical studies must be conducted to confirm that LAF is an ideal choice for clean surgery settings. |
| 49 | Empirical | Egypt | 8 ORs in one hospital | All ORs were equipped with a ceiling-mounted LAF system with an air renewal rate of 15 ACH. Some of the ORs were equipped with an additional conventional HVAC system | Ventilation function: working versus not working | Bacterial concentration, fungal concentration, and PM2.5 concentration | Ventilation function had no statistically significant effect on the bacterial and PM2.5 concentrations. Fungal concentration was higher when the ventilation system was not working, and this result was statistically significant. | Given the degree of contamination observed in the study, cleaning and maintenance guidelines for the ventilation system in OTs should be adjusted to be in compliance with published guidelines. Additionally, further studies should be conducted in order to assess the effect that hospital equipment and procedures have on indoor air quality. |
| 81 | Empirical | Austria | 80 surgeries; 40 with forced air warming devices; 40 with electric blankets; 20 surgeries of each scenario had LAF in their ORs | Specialty: orthopedic surgery | Presence of unidirectional, turbulent-free, LAF ventilation system | Airborne bacteria counts | ORs with a unidirectional, turbulent-free LAF system had decreased viable airborne bacterial counts compared to ORs not equipped with an LAF system. | The present study shows that, in the setting of a minor orthopedic surgery, the presence of a LAF ventilation system was associated with decreased airborne sedimentation. |

| Reference | Study design | Location | Sample size | OR characteristic(s) | Aspect of HVAC | Outcome | Major Takeaway | Conclusion / Recommendation |
|---|---|---|---|---|---|---|---|---|
| 82 | Empirical | England | 803,065 surgeries | Specialty: trauma surgery | LAF versus plenum ventilation systems | Incidence rate of SSIs within 90 days after surgery | The incidence rate of SSIs within 90 days following surgery was very similar for the two ventilation types. | The installation of LAF systems may not be a necessary expense, as it did not show a benefit in the present study. |
| 83 | Empirical | Spain | 8 hospitals were observed over one year, for a total of 18,910 patients | All ORs were equipped with LAF ventilation | Air renewal rate, humidity, temperature, and differential pressure | SSI rates | Decreased air renewal rate, increased humidity, and increased temperature were found to be significant risk factors for the occurrence of superficial SSIs. Differential pressure was determined to not be a significant risk factor. | The results suggest the implementation of environmental and surface contamination control protocols in an attempt to prevent SSIs. |
| 84 | CFD modeling | Turkey | Three diffuser sizes were tested in one OR, the number of trials was not specified | LAF ventilation with 30 ACH and air supplied at 0.29 m/s. OR dimensions: 6.5 m (L), 9.43 m (W), 2.43 m (H) | LAF diffuser size: 1.8 m x 2.4 m, 2.4 m x 2.4 m, 3.2 m x 3.2 m | Amount of particles deposited on the OR table | Increasing the LAF diffuser size was shown to decrease the amount of particles deposited on the OR table by 73% for smallest particles (5 μm) and 32% for largest particles (20 μm). | Increased LAF diffuser size significantly reduces particle deposition over the patient, which should be taken into account in future OR design plans. |
| 85 | Empirical | Taiwan | Three colon and rectal ORs, number of procedures not specified | OR dimensions: 7.68 m (L), 5.19 m (W), 3.0 m (H), totaling 120 $m^3$. Mean temperature of 18°C and mean relative humidity of 75% | Number of air exchanges per hour (ACH) in an OR at rest | Particulate matter concentration and airborne bacterial concentration | Varying air renewal rate between 6 ACH and 30 ACH did not have a significant influence on the particulate matter and airborne bacterial concentrations. When the HVAC system was shut down completely, however, both concentrations were significantly above the recommended limit. | When no surgeries are being performed, OR HVAC systems should maintain an air renewal rate of at least 6 ACH instead of being shut down. This will allow the air to maintain sufficiently low levels of particulate matter while still offering low energy costs during inactivity. |
| 48 | Empirical | Taiwan | Two trauma and two colorectal ORs, number of procedures not specified | OR dimensions: 7.68 m (L), 5.19 m (W), 3.0 m (H), totaling 120 $m^3$. Mean temperature of 22°C and mean relative humidity of 55 %. Before the start of the study, HVAC ran all day at 30 ACH | Number of ACHs in an active OR | Airborne particle (PM>0.5 - PM10) and bacterial concentrations | In the trauma ORs, increasing the air renewal rate from 20 ACH to 30 ACH significantly decreased the concentrations of particulate matter for all sizes. This relationship was reversed in the colorectal ORs. The concentration of airborne bacteria (CFU/$m^3$) was significantly decreased in the colorectal ORs, whereas the bacterial settle rate (CFU/h) was significantly decreased in the trauma ORs. | Recommend minimum air renewal rates of 20 ACH and 25 ACH in trauma and colorectal ORs, respectively. |

| Reference | Study design | Location | Sample size | OR characteristic(s) | Aspect of HVAC | Outcome | Major Takeaway | Conclusion / Recommendation |
|---|---|---|---|---|---|---|---|---|
| 32 | CFD modeling | Oman | 5 ventilation options with 10 trials each | Modeled based on a floor area of 56 $m^2$ and a ceiling height of 3 m. Temperature simulated as 18-24°C | 1) Conventional operating theater with HEPA filters sized at 1.2 m x0.6 m; 2) orthopedic or transplant OR with ultra-clean ventilation (UCV) system and HEPA filters sized at 2.4 m x 2.4 m (2a) and 3 m x 3 m (2b); 3) adapting option 1 to mimic the UCV system without the added cost, HEPA filters sized at (3a) 1.5 m x 0.9 m and (3b) 1.9 m x 1.1 m | Bacterial concentrations measured as BCP/$m^3$ and air speed at the wound | Ventilation options 2a, 2b, and 3a were able to achieve the class 5 cleanliness requirement of <29 BCP/$m^3$ within 0.5 m of the wound. The UCV system tended to offer better clean air distribution, but at the cost of higher air speed at the wound, which could dry it out. | A sufficiently clean environment can be achieved with a conventionally designed array of HEPA filters without the need for a costly commercial UCV. If a UCV system is used, it should be capable of producing at least 63 ACH and its supply air outlets should be fitted with 3 m x 3 m HEPA filters for use in an orthopedic or transplant OR. |
| 86 | CFD modeling | Malaysia | 12 samplings points for each of three scenarios | OR dimensions were modeled as 6.0 m (L) × 6.9 m (W) × 3.0 m (H), with exhaust grilles 0.22 m (W) × 0.46 m (H), surgical lamp 0.45 m (d) × 0.15 m (H), patient entrance 2.0 m (W) × 2.1 m (H), and personnel entrance 0.9 m (W) × 2.1 m (H). The temperature was modeled at 19°C | Three scenarios: 1) original ventilation system, 2) addition of one extra exhaust grill, and 3) addition of one extra exhaust grill and installation of a larger ceiling air supply diffuser | Air velocity measured in m/s and counts of 5 µm airborne particles settling onto the operating table | In the case where the surgical staff was modeled to be standing at a 45 degree angle over the operating table, the addition of an extra exhaust grill did not influence the particles settling onto the table. The installation of a larger air supply diffuser with the additional exhaust grill, however, reduced the number of airborne particles settling onto the operating table, as more were transported towards the exhaust grills. | |
| 87 | Empirical | Italy | 35 ORs (18 with TV and 17 with mixed airflow). 280 total air samples (140 at rest and 140 during procedures) | The median volume of the ORs was 114 $m^3$ and 140 $m^3$ for mixed and TV, respectively. The median air renewal rate was 15 ACH and 18 ACH for mixed and TV, respectively | Mixed versus TV systems | Bacterial counts measured as CFU/$m^3$ by both active and passive collection methods; particle counts | Regardless of type of ventilation and sampling method, there was a statistically significant increase in bacterial counts when the ORs were in use compared to at rest. There was no statistically significant difference in either the bacterial counts or the number of particles of >0.5 µm collected under either ventilation type. | Additional studies with more detailed protocols and repeated measurements over time are needed to further assess aspects of this study. |
| 88 | Empirical | United Kingdom | 2 orthopedic ORs (one with conventional airflow, one with LAF) | Mean temperature: 21-23°C; mean relative humidity: 44-51% | Conventional versus LAF ventilation systems | Airborne bacterial levels measured as CFU/$m^3$ in the morning and in the evening | Morning levels were considerably lower in the LAF than in the conventional flow OR, whereas evening levels were very similar. Virtually no gram-negative bacteria were detected | The results of the study draw attention to the role that regular air sampling can play in infection control surveillance plans in hospitals. |

| Reference | Study design | Location | Sample size | OR characteristic(s) | Aspect of HVAC | Outcome | Major Takeaway | Conclusion / Recommendation |
|---|---|---|---|---|---|---|---|---|
| | | | | | | | in either OR at either time of the day. | |
| 89 | Simulation | Germany | 6 scenarios with 5 repetitions each | OR equipped with low-turbulence, vertical airflow directed from ceiling to the floor (LAF) | LAF system engaged versus turned off | Aerosol propagation velocity and direction | The LAF was able to slow down the propagation of the aerosols and reverse their direction, such that the aerosol components reaching the surgeons were diluted by a factor of 10 compared to the scenario in which LAF was absent. | Tracheotomies should be performed under the presence of LAF systems in an attempt to reduce the infection risk to OR personnel. |
| 90 | Simulation | Brazil | One OR with five different scenarios (no filters, 65 %, 65 % + 85 %, 65 % + 95 %, and 65 % + 85 % + HEPA) | OR with an area of approximately 19 $m^2$ and a self-contained air conditioning system with 8 ACH of 100% outdoor air | Layout and efficiency of filters; air renewal rate measured as ACH | Particle count collected by active air sampling | The scenario that resulted in the lowest concentration of indoor particles was the combination of a 65% efficiency filter on the HVAC unit and both a HEPA filter and an 85% efficiency filter in the ventilation box. According to a simulation performed, increasing the air renewal rate from 8 ACH to 15 ACH decreased particle counts by 71%. | The use of high efficiency filtration is necessary in order to eliminate small particle size contaminants. |
| 91 | Simulation | Norway, China | 2 different air renewal rate scenarios, 3 cases each (4/6/8 exhausts), 2 different physical configurations each (upper/lower), and 4 designs of exhaust airflow. Number of trials not specified | The OR had an area of 56 $m^2$ and a height of 3.8 m. The OR was equipped with mixing ventilation, and air was supplied at 22.4°C through a prefilter in a ceiling diffuser with two air handling units (2600 $m^3$/h and 2700 $m^3$/h) | Different air renewal rates and number of exhausts | Airborne contaminants, air velocity at the wound | In the scenarios with 4 and 6 exhausts, more clean air was able to reach the wound with more ease than in the case with 8 exhausts. Increasing the air renewal rate from 18 ACH to 20 ACH did not have a significant impact on the air velocity near the wound, nor on the overall velocity distribution in the room. | In an OR with MV, supply diffusers should be placed on the ceiling and exhausts on the walls. In an OR with four supply diffusers, the highest contaminant removal efficiency can be achieved with six exhausts and 18 ACH or four exhausts and 20 ACH. |
| 92 | Empirical | Italy | 1,285 surgeries across 14 hospitals. 12 ORs had unidirectional airflow (U-OT), 6 had turbulent (T-OT), and 6 had mixed (M-OT). 61.1 % of procedures were THAs and 38.9 % were TKAs | The ORs had areas ranging from 30 to 60 $m^2$ (with a mean of 39 $m^2$) and volumes ranging from 90 to 180 $m^3$ (with a mean of 116 $m^3$). The mean air renewal rate amongst all ORs was 18 ACH | Type of ventilation: unidirectional airflow (U-OT), turbulent airflow (T-OT), and mixed airflow (M-OT) | IMA, SSI incidence, and airborne bacterial contamination measured in CFU/$m^3$ | The mean IMA was lower in the U-OT than in T-OT and M-OT, but nevertheless exceeded the recommended threshold value. No statistically significant differences were found between the three types of ventilation and the incidence of SSI. The mean airborne microbial contamination was lower in the U-OT than in T-OT and M-OT, | It is premature to discontinue the use of UDF in arthroplasty surgeries, but further research and evaluation is needed before its installation can be recommended. |

| Reference | Study design | Location | Sample size | OR characteristic(s) | Aspect of HVAC | Outcome | Major Takeaway | Conclusion / Recommendation |
|---|---|---|---|---|---|---|---|---|
| | | | | | | | but nevertheless exceeded the recommended threshold value. | |
| 93 | Simulation | Japan | Two ORs, unspecified number of trials | ISO-5 OR: 48 $m^2$ of floor space, height of 3 m, air supply of 33,131 $m^3$/h with an air renewal rate of 228 ACH. ISO-6 OR: 34 $m^2$ of floor space, height of 3 m, air supply of 10,102 $m^3$/h with an air renewal rate of 98 ACH | Comparing the performance of ventilation systems installed in ISO-5 versus ISO-6 ORs. The ISO-5 OR had a higher air supply, air renewal rate, more floor space, and an air-conditioner outlet layout (ACOL) of a larger footprint. The ACOL in the ISO-5 OR did not completely cover the patient table, whereas that of the ISO-6 OR did | Number of artificial particles shifting from the patient head into the surgical field | The number of particles shifted from the point of release at the manikin patient head into the surgical field was significantly higher in the ISO-5 OR than in the ISO-6 OR. | When designing OR ventilation systems, it is important to consider the relative positions of the ACOL and the operating table. |
| 94 | Empirical | Austria, Switzerland, Germany | 80 orthopedic surgeries: 19 surgeries performed in ORs with a large (518 cm x 380 cm) LAF system, 21 procedures in ORs with a small (380 cm x 120 cm) LAF system, and 40 procedures in ORs without an LAF system | LAF size: large = 19.83 $m^2$, small = 4.56 $m^2$. OR floor area: large LAF ORs = 49.66 $m^2$, small LAF ORs = 48.66 $m^2$, and no LAF = 33.42 $m^2$. Ceiling heights were 3 m across all three types of OR. The ORs were all 17 years old but had undergone updates | Comparing ORs without an LAF system to ORs equipped with small or large LAF systems. ORs without an LAF system received a fresh air volume of 1170 $m^3$/h and an air renewal rate of 12 ACH. ORs with a small LAF system received a fresh air volume of 3880 $m^3$/h and an air renewal rate of 26 ACH. ORs with a large LAF system received a fresh air volume of 1300 $m^3$/h and an air renewal rate of 178 ACH with 20% recycled air | Airborne bacterial contamination measured as CFU/$m^3$ | For all five sampling locations, the airborne bacterial contamination was the lowest in the large LAF ORs, followed by the small LAF ORs, then the non-LAF ORs. This relationship was only statistically significant for the sampling located on the instrument table, however. | The results suggest that the presence of an LAF system alone is insufficient to ensure a particle- and bacteria-free instrument table. |
| 95 | Empirical | Italy | 71,655 surgical procedures in 15 ORs over 12 years. The study had two phases: 2001 to 2003, ORs equipped with low-turbulence displacement air supply systems (21,000 procedures), and 2004-2013, ORs equipped with LAF supply systems (50,655 procedures) | | Low-turbulence displacement air supply system versus LAF supply systems | Airborne particles measured as particles/$m^3$ and airborne bacterial contamination measured as UFC/$m^3$ | Use of a LAF supply system decreased the mean number of particles from 189.96 particles/$m^3$ to 3.92 particles/$m^3$ compared to low-turbulence supply. Additionally, use of an LAF supply system decreased microbial air contamination from 40.02 UFC/$m^3$ to 1.46 UFC/$m^3$ compared to low-turbulence supply. Both of these results were statistically significant. | The results show that LAF was effective for both clean and contaminated/dirty procedures, thus adding to the conflicting literature. |

| Reference | Study design | Location | Sample size | OR characteristic(s) | Aspect of HVAC | Outcome | Major Takeaway | Conclusion / Recommendation |
|---|---|---|---|---|---|---|---|---|
| 96 | CFD modeling | Turkey | 1 OR and 4 different exhaust configurations | | Configuration of exhaust outlets | Distribution of particles in the OR | Use of both floor and ceiling exhaust outlets decreases particle counts near the operating table. | Recommend use of both floor- and ceiling-level exhaust outlets. |
| 33 | CFD modeling | Iran | 1 OR and 9 different air velocities | | Rate of airflow | Deposition of particles over the wound site | There is an optimal airflow velocity around 0.10-0.12 m/s that minimizes particle deposition at the wound. | The rate of airflow should be balanced with lights and temperature to avoid disrupting the thermal plume over the wound tissue. |
| 97 | Modeling | USA | 3 ORs in different hospitals | | Air renewal rates of 15 ACH, 20 ACH, or 25 ACH | Air velocity, particle counts, and airborne bacteria measured as CFU | Although increased air renewal rate was shown to decrease airborne bacterial counts in some cases, this observation was not consistent across ORs or even across sampling locations within the same room. Similarly, no consistent relationship was observed between increased air renewal rate and particle counts. | The results suggest that environmental quality indicators (EQIs) such as particle counts and air velocity at relevant locations in the OR might be better indicators of air cleanliness than air renewal rates. These provide more accurate measures of air quality and offer greater potential in minimizing the risk of SSIs. |
| 98 | Empirical | The Netherlands | 101 ORs across 22 hospitals | | Air supply canopy size, shape, and rate of airflow | EPR is defined as the negative log of the ratio of particles in the clean/protected area to the particles outside of the clean/protected area | Canopy shape and size, air speed, presence and height of a screen, and the type of air exchange system were all significant predictors of the EPR. | A rectangular canopy with two angled corners (irregular hexagonal shape) led to the best positive effect of EPR, and this information should be used in designing future ventilation systems. |
| 99 | Simulation | USA | 3 tests in 3 different ORs | | Vent placement, functioning, and type of diffuser: single large diffuser, multi diffuser array, or 4-way throw diffuser | Rate of airflow, temperature, particle counts, and clearance of microbes and $CO_2$ | The OR with a single large diffuser cleared microbes and $CO_2$ most effectively; the multi diffuser array best minimized total particles. | Recommend UDF of clean air with low-wall return grilles. |
| 100 | Empirical | United Kingdom | 252patients total; 95 in the LAF group and 157 in the TV group | Specialty: orthopedic surgery | Use of LAF versus TV | Incidence rate of SSIs | The rate of early SSI was slightly higher under turbulent flow, but this finding was not statistically significant. | The sample size of this study was small, and the results are inconsistent across other studies. Therefore, more research is needed to determine if LAF is protective against SSIs. |

| Reference | Study design | Location | Sample size | OR characteristic(s) | Aspect of HVAC | Outcome | Major Takeaway | Conclusion / Recommendation |
|---|---|---|---|---|---|---|---|---|
| 101 | CFD modeling | Brazil | Used 57 measurements of temperature and velocity at 5 points in the room to inform 5 model ORs with variable inlet/outlet locations | Dimensions: 5.7 m (L) x 5.4 m (W) x 3.14 m (H), surface area of 30.78 $m^2$. Doors: main door (1.5 m x 2.1 m, double door); secondary (1.0 m x 2.1 m, simple door). HVAC: Inlet and outlet dimensions (0.9 m x 0.3 m) | Various inlet and outlet layouts being compared to "Case 0," the real-life OR on which the models are based. Two inlet scenarios: A) sidewall (conventional high supply) inlet systems and B) ceiling (four-way supply, 0.4 m x 0.4 m). Two outlet (exhaust) scenarios: 1) outlet on the same wall as in Case 0 and 2) outlet on the opposite wall as in Case 0 | Air motion and distribution | Compared with Case 0 and type B cases, type A cases had lower magnitude velocity vectors in the center of the room, making it more difficult for this type of ventilation to move particles from the sterile field to the outskirts of the OR. In the XZ plane, none of the cases had flow vectors indicating the downward motion of clean air from the ceiling onto the operating table. In the YZ plane, however, the case 2B model clearly showed the movement of clean air from the ceiling down onto the operating table and off to the side. | Use of conventional high supply promotes a more adequate environment for an OR. The four-way suppliers presented a greater velocity magnitude close to the floor, which could entail the lifting of settled particles into the surgical field, thus risking contamination. |
| 102 | CFD modeling | USA | Unspecified number of trials | | Air renewal rate: 15 ACH, 23 ACH, and 31 ACH | Airborne particulates and airflow patterns | Higher air renewal rates may reduce the recirculation of particles, but they do not alter airflow patterns. | Configuration of the HVAC system is likely an important factor in the movements of airborne contaminants. |
| 103 | CFD modeling | USA | Unspecified number of trials | All configurations were modeled with UDF, an air renewal rate of 23 ACH, and air supplied at 21°C. The legacy and ceiling exhaust designs had an average airflow discharge velocity of 153 L/s/$m^2$, while the distributed supply had a discharge velocity of 114 L/s/$m^2$ | HVAC configuration: legacy design, ceiling exhaust, and distributed supply | Movement of airborne contaminants modeled as originating from two surgeons and a nurse inside the sterile field | In all three configurations, particles originating inside the sterile zone are swept out and away from such zone and no entrainment is observed. In the ceiling exhaust and distributed supply conditions, however, the particles are forced upward. | When attempting to reduce airborne contamination, modifications of legacy HVAC configurations at a low air renewal rate should be considered prior to increases in air renewal rate, which can lead to increased energy costs. |
| 104 | Empirical | South Asia | 363 neurosurgical cases between 2012 and 2013, plus 623 neurosurgical cases between 2013 and 2016 | | Replacement of the OR air filtration system after its first ever routine servicing found it to be defective | Rate of positive CSF cultures and PCNSIs | In the year before the air system replacement, there were seven positive CSF cultures and 71 PCNSIs out of 363 cases (1.9% and 19.6%, respectively). During the following three years, there were zero positive CSF cultures and four PCNSIs out of | Based on their findings, the authors advise that medical professionals acknowledge the role that the OR environment can play in the incidence of postoperative infections. They recommend that, when postoperative |

| Reference | Study design | Location | Sample size | OR characteristic(s) | Aspect of HVAC | Outcome | Major Takeaway | Conclusion / Recommendation |
|---|---|---|---|---|---|---|---|---|
| | | | | | | | 623 cases (0% and 0.6%, respectively). | infections are inexplicably high, the environment be surveyed for defects. |
| 105 | Modeling | Italy | 19 ORs (14 with TV and 5 with LAF) were surveyed a total of 59 times under operating conditions | OR volumes ranged from 74 $m^3$ to 164 $m^3$ | LAF versus TV systems; overall air renewal rates | Airborne bacterial contamination measured as CFU/$m^3$ | LAF was associated with lower contamination as compared to TV (medians of 12 CFU/$m^3$ and 71 CFU/$m^3$, respectively). Overall, contamination decreased with increasing air renewal rates. | The reference air renewal rate value of 15 ACH should be adjusted according to the ventilation system in use. In ORs with LAF ventilation, the reference value should be increased to ensure that airborne microbial contamination remains below the 20 CFU/$m^3$ limit. Alternatively, for ORs with TV, the reference value of 15 ACH could be lowered, so long as the 180 CFU/$m^3$ limit is still met. |
| 47 | Empirical | Pakistan | Two orthopedic ORs, one in each hospital (Services hospital and Shalimar Hospital) | The orthopedic OR at the Services hospital (OOT1, 30.73 $m^2$) was equipped with "natural" ventilation system, whereas that at the Shalimar Hospital (OOT2, 30.56 $m^2$) was equipped with LAF | Presence of LAF system in an orthopedic OR | Particulate matter of various sizes and $CO_2$ levels | The concentration of particulate matter of all sizes was lower in the orthopedic OR equipped with LAF than that with natural ventilation, and this result was statistically significant. Additionally, the $CO_2$ concentration under LAF conditions was less than one third that of the natural ventilation condition. | The presence of vertical LAF ventilation was found to be effective at refining the air quality in orthopedic ORs. |
| 106 | Empirical | United Kingdom | Number of patients: 74 (mixed-used emergency theatre ), 85 (LAF theatre); chosen from an original sample size of 1370 | | Review of patient records to determine risk of SSI (additionally correlated to reported length of stay and mortality rate) when patients were operated on while in the presence of LAF versus when not (in a mixed-use emergency theatre) | Risk of SSIs | The risk of SSIs among patients who underwent hip fracture surgery in mixed-use emergency theatres (without LAF) was no different than that for surgeries conducted under LAF conditions. | This was a retrospective cohort study with no control for confounders; therefore, further research that controls for confounding variables should be conducted before direct conclusions can be made regarding the impact of LAF on SSI risk. |
| 107 | Simulation | USA | Four ceiling air delivery layouts tested, number of trials not specified | OR dimensions: 6.2 m (W) x 6.3 m (L) x 3.0 m (H). Pressure | Comparing ventilation systems equipped with an air curtain (AC), multi-diffuser array (MDA; supply | Microbial counts at the patient table reported as CFU/$m^3$ | Zero CFU/$m^3$ were found for all SLD scenarios, with the exception of the 57 $ft^2$ option with 30 fpm, for which 2 | The results show that concentrating uniform LAF over the operating table using a single large |

| Reference | Study design | Location | Sample size | OR characteristic(s) | Aspect of HVAC | Outcome | Major Takeaway | Conclusion / Recommendation |
|---|---|---|---|---|---|---|---|---|
| | | | | kept at 2.5 Pa and 17°C | velocities 30 and 50 fpm), or a single large diffuser (SLD; layout options 57 vs 86 $ft^2$, supply velocities 30 and 50 fpm) | | $CFU/m^3$ were identified. The MDA cases had 11 and 24 $CFU/m^3$ for air supply velocities of 30 and 50 fpm, respectively. In the AC scenario, 87 $CFU/m^3$ were found. | diffuser can move airborne contaminants away from the patient in a predictable manner, thus assisting in the maintenance of a sterile field. |
| 108 | Simulation | USA | 3 ORs; 15 mock surgical procedures | Two modern ORs (built in 2017) with an area of 55 $m^2$ and one older OR (built in 1992) with an area of 44.3 $m^2$ | Comparing two modern ceilin g-mounted airflow systems (single large diffuser and multiple diffuser array designs) to an antiquated four -way throw diffuser system. Both modern systems were tested at air renewal rates of 20 ACH and 26 ACH, while the old design was only tested at 26 ACH | Relative particle concentration measured at the instrument table and at a low-wall return grille | The concentration for all particle sizes was significantly greater at the return grill than at the instrument table for the single large diffuser at 26 ACH, and the multiple diffuser arrays at both 20 ACH and 26 ACH. Alternatively, the single large diffuser at 20 ACH and the older room's four-way throw diffuser systems only had statistically significant greater concentrations at the return grille for particles greater than or equal to 5.0 microns. | Recommend personnel awareness that in an OR it may be a counter-productive strategy to move away from the patient toward a wall, which is where air return grills are located. |
| 109 | Empirical | Norway | 51,292 primary THAs assessed across 40 hospitals | Unidirectional ventilation split into high- and low-volume based on volume flow rates above and below 10,000 $m^3/h$, respectively. These were further split into small and large based on canopy sizes below and above 10 $m^2$, respectively | Comparing four different ventilation systems: large, high-volume, unidirectional vertical airflow/vertical LAF (hvUDVF); small, low-volume, unidirectional vertical airflow/vertical LAF systems (lvUDVF); unidirectional horizontal flow/horizontal LAF systems (UDHF); and conventional turbulent airflow | Number of surgeries requiring revision due to infection | THAs performed in ORs with hvUDVF systems had lower risk of revision due to infection compared to THAs performed in ORs with conventional turbulent airflow systems. THAs performed in ORs with lvUDVF or UDF had a risk of revision due to infection similar to that of THA performed in conventional turbulent airflow. | The authors recommend the use of hvUDVF systems for all ultraclean surgery settings due to the reduction in particle count and microbial loads compared to conventional turbulent airflow systems, therefore leading to a lowered infection risk. |
| 110 | Empirical | Italy | 175 ORs across 31 hospitals | ORs had an average surface area of 38.2 $m^2$ and an average volume of 114.1 $m^3$. The differential pressure between the OR and adjacent areas was always | Four ventilation systems were compared: A) Partial Unidirectional Airflow designed for ISO 5 cleanliness; B) Partial Unidirectional Airflow designed for ISO 7 cleanliness; C) Mixing Airflow designed for ISO 7 | Airborne bacterial contamination measured as $CFU/m^3$ | Under at rest conditions, ventilation system A had the lowest average contamination at 0.9 $CFU/m^3$ and type D had the highest at 11.8 $CFU/m^3$. Under operating conditions, ventilation system A had the lowest average at 5.5 $CFU/m^3$ and type C had the highest at 72.8 $CFU/m^3$. | Regardless of the condition (at rest or under operation), the partial unidirectional airflow system designed to meet ISO-5 air cleanliness conditions was shown to be the most effective at cleaning the OR air. |

| Reference | Study design | Location | Sample size | OR characteristic(s) | Aspect of HVAC | Outcome | Major Takeaway | Conclusion / Recommendation |
|---|---|---|---|---|---|---|---|---|
| | | | | positive, averaging 10 Pa | and adopting high-wall supply grills; and D) Mixing Airflow designed for ISO 7 and adopting ceiling air diffusers. Average air renewal rates were 51.2 ACH, 19 ACH, 15.1 ACH, and 18.4 ACH for systems A through D, respectively | | | |
| 111 | CFD modeling | Iran | Four scenarios were investigated, but the number of trials was not specified | The OR was modeled with a square floor plan of dimensions 6 m x 6 m and a ceiling height of 3.15 m. The air renewal rate was modeled as 25 ACH. The LAF diffuser velocity was held at 0.25 m/s in all scenarios | No air curtain compared to three different air curtain velocities: 1.5 m/s, 2.5 m/s, and 3.5 m/s | Contamination at the surgical bed ("contamination" was not defined by the authors) | Of the four scenarios, the lowest level of contamination was observed with the air curtain set at 1.5 m/s, followed by the scenario with the air curtain at 2.5 m/s. The level of contamination was essentially the same between the no air curtain and 3.5 m/s scenarios. | Air curtains serving as boundaries surrounding LAF systems help further prevent the entrance of contaminants into the sterile zone, further directing the clean laminar air to move down and outwards. The velocity of this air curtain, however, is essential to its proper functioning, and it must be optimized based on the velocity of the laminar supply it surrounds. |
| 112 | Simulation | India | Five patients | Ophthalmic clinic outpatient OR | Ceiling-mounted LAF turned on versus off | Pathway of exhaled air from patients | When the LAF ventilation was turned off, the exhaled air from patients flowed freely towards the surgeons' breathing area. When the LAF was turned on, however, it successfully directed the exhaled air away from the surgeons. | The use of LAF is helpful in significantly dampening and dissipating air exhaled from the patient in its travel towards the surgeon, thus offering added protection to HCWs against possible infections. |
| 113 | Empirical | Germany | 33,463 elective hip prosthesis procedures due to arthrosis (HIP-A) from 48 hospitals, 7,749 urgent hip prosthesis procedures due to fracture (HIP-F) from 41 hospitals, and 20,554 knee prosthesis (KPRO) procedures from 38 hospitals | Ventilation systems installed between 1990 and 2004 | Presence and size of LAF system | Severe SSI rates | For all three prosthesis procedure types, neither the presence nor the size of an LAF ceiling was associated with lower infection risk. Comparing hospitals with large LAF ceilings (at least 3.2 m x 3.2 m) with those with no LAF also failed to demonstrate lowered SSI rates. | LAF ceilings may not be needed for orthopedic surgeries, at least for the hip and knee procedures studied. |

| Reference | Study design | Location | Sample size | OR characteristic(s) | Aspect of HVAC | Outcome | Major Takeaway | Conclusion / Recommendation |
|---|---|---|---|---|---|---|---|---|
| 114 | Empirical | Taiwan | Three ORs, each sampled 10 times for 4 consecutive hours over an 11-month period | The two trauma ORs measured 6.6m (L) × 5.43 m (W) × 3.0 m (H), volume 107.59 $m^3$. The ENT OR measured 6.42 m (L) × 5.43 m (W) × 3.0 m (H), volume 119.12 $m^3$ | Comparison of RDAC (ENT OR), conventional HVAC (trauma OR), and LDAC (trauma OR) ventilation systems | Suspended particulate matter concentration, $CO^2$ concentrations, and airborne bacterial concentration | The PM≥0.5concentration of the OR using the conventional HVAC system (101.53 particles/L) and that of the OR using the LDAC system (88.08 particles/L) were significantly higher than that of the OR using the RDAC system (21.33 particles/L). Similar results were found for PM10, PM2.5, and PM1 in the three ORs. The $CO^2$ concentration of the OR using the LDAC system (421.20 ppm) was significantly higher than that of the OR using the conventional HVAC system (357.51 ppm) and the one using the RDAC system (408.88 ppm). There was no statistically significant difference in airborne bacteria between the three ORs. | The PM≥0.5 and airborne bacterial concentrations achieved by all three systems when at rest met both the ISO and Chinese air quality standards. The RDAC system offered the lowest total energy consumption. |
| 46 | Empirical | Sweden | Six ORs total, 3 with LAF and 3 with DV. 30 surgeries in DV ORs and 33 surgeries in LAF ORs | Dimensions: LAF and DV rooms had areas of 46 $m^2$ and 39 $m^2$, respectively. All ORs were around 20°C | LAF (9,160 $m^3$/h, 0.25 m/s) versus DV (2,430 $m^3$/h, 0.09-0.15 m/s) | Airborne bacterial contamination measured as CFU/$m^3$ | Use of an LAF system reduces bacterial contamination by 89% as compared with the DV system. In LAF ORs, 2 out of 164 samples had unacceptable bacterial growth exceeding 10 CFU/$m^3$. In DV ORs, 52 out of 91 samples exceeded 10 CFU/$m^3$. | As with all ventilation systems, regular maintenance of LAF systems is crucial in ensuring the safety of patients. In order to prevent future problems, knowledge needs to be shared between the technical and medical worlds regarding ventilation systems. |
| 115 | CFD modeling | Iraq | One trial in one OR | Equipment and dimensions: OR (7.10 m x 7.10 m x 3 m); plenum box (2 m x 2 m); upper grill (x4) (18 m x 18m) each; lower grill (x4) (48 m x 25 m) each; patient bed (2 m x 0.75 m) | Two cases with varied placement/location of vents: 1) four large exhaust grills in lower corners and four small exhaust grills in upper corners and 2) only four large exhaust grills in four lower corners | Airflow, pressure, temperature, and humidity | No clear impact was observed with the addition of four small grills in the upper corners of the room for case 1. | Achieving positive pressure inside the OR is possible and can be easily achievable when exhausting 85% of the air generated from the plenum box. Careful choice of the location of the plenum box is important in achieving this. |
| 116 | Empirical | Italy | 1,425 air samples from 29 OTs over 9 years | | TV vs LAF systems | TBCs measured as CFU/$m^3$ | A statistically significant difference was identified between LAF and TV systems, with | Environmental monitoring in the OR is important in order to ensure that |

| Reference | Study design | Location | Sample size | OR characteristic(s) | Aspect of HVAC | Outcome | Major Takeaway | Conclusion / Recommendation |
|---|---|---|---|---|---|---|---|---|
| | | | | | | | higher bacterial contamination rates found under the latter. | acceptable levels of air quality are constantly being met. To achieve this, the engineering and hygiene departments need to cooperate in the creation of a monitoring program. |
| 117 | CFD modeling | Iran | Number of trials not specified | The OR was modeled with the dimensions 6.3 m (L) x 6.3 m (W) x 3 m (H) and an area of 37 $m^2$ | Inlet air velocity in an UCV system | Particle deposition on the wound across various particles sizes (measured as percent of total particles released). | The optimum inlet air velocity was found to be about 0.1 m/s and is independent of particle size. A lower inlet air velocity is incapable of pushing particles away from the wound, and a higher velocity can cause turbulence above the wound that increases the risk of contamination. | The use of fixed partitions in the UCV system decreases particle deposition on the found without changing the optimum inlet air velocity, but removable partitions have shown negative effects on air quality. Thus, removable partitions should be used cautiously, although further study is needed. |
| 118 | CFD modeling | Sweden | Number of trials not specified | The OR was modeled as orthopedic with dimensions 6.4 m (L) x 6.3 m (W) x 3 m (H) and an ambient temperature of 20ºC | Three different ventilation schemes: conventional (turbulent) airflow, LAF and TAF. | BCPs measured as CFU/$m^3$ and CFU/$m^2$/h. | LAF and TAF both showed BCP counts at the operating table that were below 0.1 CFU/$m^3$, well below the standard threshold of 10 CFU/$m^3$ for airborne contamination, while turbulent flow resulted in about 30 CFU/$m^3$. The turbulent flow simulations found that increasing the air renewal rate from 26 to 40 ACH improved air quality, and further increasing to 60 ACH reduced contamination to 3.6 CFU/$m^3$ at the operating table and surface contamination dropped to 34 CFU/$m^2$/h. | While both TAF and LAF are better than conventional airflow at reducing contamination, TAF is more energy-efficient. Future work should not consider the dilution model as an appropriate way to predict OR contamination. |
| 21 | Empirical | Italy | 35 orthopedic ORs in 30 hospitals. 17 OTs had mixed flow (M-OT) and 18 with turbulent flow (T-OT) ventilation systems | | Comparing mixed flow and turbulent flow ventilation systems. Also compared symmetric versus asymmetric HVAC extraction arrangement | TVC measured as CFU/$m^3$ | The mean TVC in M-OTs was 38.9 CFU/$m^3$, while in the T-OTs this was 49.6 CFU/$m^3$. The mean TVC in OTs with asymmetric arrangement was 49.4 CFU/$m^3$, while in those with symmetric this was 41.8 CFU/$m^3$. Neither one of these differences was statistically significant, however. | No statistically significant differences between type of ventilation or HVAC layout were identified. Further studies are needed before recommendations can be made. |

| Reference | Study design | Location | Sample size | OR characteristic(s) | Aspect of HVAC | Outcome | Major Takeaway | Conclusion / Recommendation |
|---|---|---|---|---|---|---|---|---|
| 119 | Empirical | Germany | 12 TKAs, 6 in a LAF theater and 6 without LAF venti lation | The LAF system was 3.2 m x 3.2 m with an air volume flow of 11,400 $m^3/h$ and a vertical flow of 0.22 m/s | Use of LAF ventilation in the OR | Particle load in various locations inside the OR | The particle load was significantly reduced with LAF, regardless of particle size, even outside the boundary of the LAF area. This result was observed throughout the entire length of the surgery. | It is worthwhile to note that the reduction in respirable particles under LAF could benefit the health of the surgical team even if the rate of SSIs is unaffected. |
| 120 | Empirical | Singapore | 1,028 total knee replacements (TKRs) performed by the same surgeon, 453 of which were conducted under LAF | | Use of LAF ventilation in the OR | Incidence of PJIs | The overall incidence of PJI was 0.6%, with three infections occurring in the LAF condition and three in the non-LAF condition. There was no statistically significant difference. | Performing TKRs in conventionally ventilated ORs remains safe and cost-effective. |
| 121 | Empirical | New Zealand | 51,485 total hip replacement (THR) surgeries and 36,826 TKR surgeries, 35.5% o f which were performed in LAF ORs | | Use of LAF ventilation in the OR | Percentage of patients requiring early revision for deep infection | Operations performed under LAF had more deep infections requiring revision: hip replacements had an infection rate of 0.148% under LAF and 0.061% under CV; knee replacements had an infection rate of 0.193% under LAF and 0.100% under CV. Both of these results were statistically significant. | Use of LAF during joint replacement surgeries does not protect against deep infection and may actually increase risk of revision. |
| 122 | CFD modeling | Turkey | Two ORs, each modeled with a different outlet grille configuration | Inlet air is provided through 3.2 m x 3.2 m LAF unit. Outlet air is provided through two 0.525 m x 0.525 m outlet ports near the floor and two 0.425 m x 0.325 m outlet ports near the ceiling | Two outlet exhaust configurations: 1) four outlet grilles located in only two corners and 2) eight outlet grilles located in all four corners of the room | Temperature, humidity, and air velocity distributions at multiple points in the room | No differences in temperature or humidity were observed between the two configurations, regardless of location of measurement in the room. The air velocity distribution proved best with configuration 2. The air curtain effect was successfully modeled with the eight outlet grille configuration, but not with the four outlet grille configuration. | Installing outlet grilles in all four corners of the room is most appropriate, since this configuration provides the proper air distribution. Additionally, simply increasing the air inlet velocity in the two-corner configuration is not an appropriate solution. |
| 123 | CFD modeling | Italy | Number of trials not specified | Modeled as an orthopedic OR equipped with UDF and an air renewal rate of 25 ACH | Comparing two ventilation schemes: V1) 80% supply air sourced from a diffuser at 0.60 m/s and 20% from an air curtain at 2.02 m/s, and V2) 100% supply air sourced from a diffuser at 0.74 m/s | Air temperature distribution and indoor air quality (IAQ) | The ventilation scheme equipped with vertical UDF provided an ultra-clean environment and better comfort conditions for all OR zones, proven by the IAQ measurement data. The ventilation scheme with the UDF can be used as an alternative only | The combination of a high surface area air supply diffuser and an air curtain to control the direction of LAF was identified as essential in ensuring proper airflow to reduce |

| Reference | Study design | Location | Sample size | OR characteristic(s) | Aspect of HVAC | Outcome | Major Takeaway | Conclusion / Recommendation |
|---|---|---|---|---|---|---|---|---|
| | | | | | | | if the supply and return diffusers are positioned properly in relation to the location of various objects in the OR. | contamination of the breathing zones in the OR. |
| 124 | Empirical | Italy | 22 TV ORs and 5 mixed flow ORs | | Comparing TV and mixed flow ventilation systems in OTs at rest | Particle counts greater than 0.5 µm in size, total microbial count measured as CFU/$m^3$, and fungi concentration measured as CFU/$m^3$ | The median particle counts for the turbulent and mixed ventilation ORs were 372.5 and 491 particles, respectively. The median microbial and fungi concentrations were zero for both ventilation systems. The statistical significance of these results was not specified. | To control air contamination, the authors recommend consistent air quality monitoring programs, the development of tools for staff training, and consistent maintenance and testing of HVAC systems. |
| 74 | CFD modeling | Sweden | Three ORs, 15 orthopedic case s in each OR | | Comparing three types of ventilation: LAF (12,000 $m^3$/h), TMA (3,200 $m^3$/h), and TCAF (5,600 $m^3$/h) | Bacterial contamination measured as CFU/$m^3$ at three different locations: 40 cm from the wound, at the instrument table, and in the periphery of the room | ORs equipped with TMA ventilation had median CFU counts (in all three measured locations) greater than or equal to the recommended level to prevent SSIs. The median values for LAF and TcAF ORs were below the threshold at all three locations. | TMA ventilation should not be used in ORs where infection-prone surgeries take place, as this type of ventilation cannot offer the recommended limit of less than or equal to 10 CFUs/$m^3$. |
| 125 | CFD modeling | USA | Five cases run in a commercial CFD program for a total of 5,000 iterations | The OR was modeled with dimensions 6.1 m (L) x 5.8 m (W) x 2.9 m (H), ambient temperature of 20ºC and a pressure of 2.5 Pa. The air renewal rate was modeled as 31.6 ACH with a supply air temperature of 18.3ºC | Comparing five different ventilation configurations: cases 1-3 modeled air curtains with air supplies of 120 $ft^3$/min, 80 $ft^3$/min, and 100 $ft^3$/min, respectively; case 4 modeled an air curtain system that is smaller than traditional systems, encompassing only the operating table and therefore falling between the patient and the surgical staff; and case 5 modeled a larger LAF diffuser, with an area of 7.46 $m^2$ as opposed to the 7.06 $m^2$ in the base case | Contamination levels at patient wound site and back table site | Cases 1-3 had the highest contamination levels of all cases at both modeled sampling sites. The large LAF diffuser case had virtually no contamination at either measurement site. | The use of air curtains as specified by manufacturers may not provide better indoor environmental results compared to baseline models. Maximizing the area of the LAF diffusers remedies the issues of being unable to achieve UDF pattern and therefore adequately achieving optimal air asepsis. |
| 126 | Empirical | Germany | 1,286 surgeries performe d by the same surgical team across five O Rs, three equipped with TMV and tw o with UDF | The 3 TMV ORs had volumes of 103 $m^3$; one UDF OR had a volume of 94 $m^3$ and the other two had volumes of 112$m^3$. The TMV supply air | Comparing TMV versus UDF | Intraoperative airborne bacterial burden, measured as total CFU and CFU/h | The average bacterial burden was 94 % lower in ORs equipped with UDF as opposed to those equipped with TMV. The average and maximum values of both total CFU and CFU/h were lower in UDF ORs compared to TMV ORs. | UDF is extremely effective at decreasing airborne bacterial burden, much more so than TMV. |

| Reference | Study design | Location | Sample size | OR characteristic(s) | Aspect of HVAC | Outcome | Major Takeaway | Conclusion / Recommendation |
|---|---|---|---|---|---|---|---|---|
| | | | | volume was 2,200 $m^3$/h in two ORs and 1,600 $m^3$/h in the third. The UDF ceiling size was 3.2 m x 3.2 m with a supply air volume of 9,000 $m^3$/h | | | | |
| 127 | Empirical | Nigeria | 127 surgical cases across two OTs | | Quality of ventilation (this was not further specified by the authors) | Odds and rates of SSI incidence | Surgeries conducted in an OR that was "moderately ventilated" had 34 times the odds of SSI as compared to surgeries conducted in "well-ventilated" ORs. The rate of SSI was the same between the two types of ventilated ORs. | Hospital management should employ prevention strategies and modify risk by improving ventilation. |
| 128 | CFD modeling | China | Number of trials was not specified | Conducted in an ISO-5 cleanroom for experimental validation. Dimensions were 7 m (L) x 6 m (W) x 3 m (H) | Four types of ventilation schemes: vertical LAF, horizontal LAF, DVAF, and TAF ventilation. These were also compared at five different air renewal rates | BCP concentration recorded as CFU/$m^3$ | Under the DVAF and TAF conditions, BCP concentrations were below the recommended limit of 10 CFU/$m^3$ at every sampling point on the operating table and instrument table 1. The samples at instrument table 2, however, had similar contamination across all four scenarios, with averages around 10 CFU/$m^3$. The vertical LAF and TAF systems reached maximum BCP removal efficiency at 60 ACH, while horizontal LAF and DVAF reached theirs at 70 ACH. | The degree of air cleanliness depends not only on the flow rate of the air supply, but also on the airflow distribution across the room. TAF ventilation is recommended over the other three ventilation systems. |
| 129 | CFD modeling | Taiwan | Number of trials was not specified, but all measurements were taken in an unoccupied OR | The OR was modeled as an ISO-7 environment with dimensions 6.5 m x 5 m x 3 m, temperature 22ºC, relative humidity 55%, pressurization 6 Pa, and an air renewal rate of 39 ACH | Air renewal rates of 20 ACH, 10 ACH, and 5 ACH, as compared to the usual 39 ACH recommended for the ventilation system being studied | Particle concentration and microbial counts at thirteen sampling locations, recorded as particles/$m^3$ and CFU/$m^3$, respectively | The level of contamination (in both particles and microbial counts) at all sampling locations was extremely consistent across all four air renewal rate conditions. Most importantly, the contamination at all sampling locations was kept below the recommended values of 350,000 particles/$m^3$ and 100 CFU/$m^3$ regardless of air renewal rate. | Lowering the air renewal rate in an unoccupied OR during non-operational hours is acceptable, and can be conducted with levels as low as 5 ACH without sacrificing air quality or violating recommended guidelines. |
| 45 | Empirical | Italy | 1,228 elective procedures (hip or knee surgery) performed across | The OR volumes varied from 90 $m^3$ to 180 $m^3$, with an average of 116 $m^3$. | Type of ventilation system: vertical UDF (U-OT), turbulent airflow (T-OT), and mixed airflow (M-OT). | Air microbial contamination recorded as IMA and CFU/$m^3$. | The percentage of ORs within the threshold values for IMA and CFU/$m^3$ was highest in U-OTs, but this was still less than half. | A significant number of OTs were found be out of compliance with recommended thresholds |

| Reference | Study design | Location | Sample size | OR characteristic(s) | Aspect of HVAC | Outcome | Major Takeaway | Conclusion / Recommendation |
|---|---|---|---|---|---|---|---|---|
| | | | 14 hospitals and 28 OTs (16 with vertical UDF, 6 with TV, and 6 with mixed airflow) | The mean air renewal rate across all ORs was 18 ACH | | | The mean IMA and CFU/m$^3$ recorded were lowest in U-OTs, followed by T-OT. | for air microbial contamination. Given this observation, it is essential to increase awareness amongst healthcare professionals regarding the risks associated with their behaviors in the OR. |
| 58 | Empirical | USA | 81 orthopedic cases across three ORs; most procedures were TJAs | The average OR temperature was 65.1°F (range = 62°F - 68°F) and the average humidity was 57.4 mmHg (range = 39 mmHg - 64 mmHg). All ORs were equipped with partial ceiling supply, vertical LAF systems | Sample plates located outside versus inside of the LAF | Contamination rate and presence of *Staphylococcus aureus*, *Bacillus* species, *Micrococcus* species, diphtheroids, mold, and gram-negative rods on sample plates | The contamination rate was consistently lower inside rather than outside the LAF, and this result was statistically significant. All organisms had similar distributions among the contaminated plates, regardless of position relative to the LAF. It was not able to be determined, however, that LAF offers added protection in the absence of people and door openings. | The results indicate that operating under LAF is protective for at least the first 90 minutes of surgery, as the sample size for longer surgeries was too small to allow a determination. However, larger prospective studies that independently examine the influence of LAF on contamination should be conducted. |
| 130 | Empirical | New Zealand | 91,585 THAs performed between 2000 and 2014 | | Presence of conventional (55,391 cases) versus LAF (35,373 cases) ventilation | Incidence of revisions at 6 and 12 months due to PJIs | The incidence of PJI after 6 months was 0.11% for conventional airflow and 0.20% for LAF, a statistically significant result. Similarly, after 12 months, the incidence for conventional airflow was 0.17% and that of LAF was 0.25%. The presence of LAF was found to increase the incidence of PJIs by up to 1.9 times. | While the theoretical benefits of LAF are well established, joint registry analyses such as this one demonstrate a reality in which the application of this type of ventilation is not as simple. The nature of LAF makes it sensitive to the positioning and movement of equipment and personnel, such that its theoretical benefits are negated by improper functioning. |
| 131 | Empirical | Sweden | 13 surgeries performed across two ORs | The OR with UWD measured 36 m$^2$ in area and 108 m$^3$ in volume, while that with UDF plus mixing ventilation measured 100 m$^2$ in area and 270 m$^3$ in volume | Comparing two types of ventilation: UWD (19 each) and downward UDF combined with MV (57 ACH) | UFP concentration and airborne microbial contamination measured as particles/m$^3$ and CFU/m$^3$, respectively | UFP concentration was lower on average under UDF with MV, but was also affected by surgery type. Microbial contamination under UWD was higher, averaging 20 CFU/m$^3$, versus less than 1 CFU/m$^3$ on average under UDF with mixing ventilation. | The OR with a hybrid ventilation system had better air quality than the OR with UWD ventilation alone. |
| 132 | Empirical | UK | 40 surgeries (29 TKAs and 11 | | LAF versus non-LAF OTs | Bacterial contamination on sterile surgical | Based on the direct inoculation assessment, the swabs of sterile | Touching the sterile surgical helmet system |

| Reference | Study design | Location | Sample size | OR characteristic(s) | Aspect of HVAC | Outcome | Major Takeaway | Conclusion / Recommendation |
|---|---|---|---|---|---|---|---|---|
| | | | THAs) performed by three surgeons across two ORs | | | helmets, determined by swabbing helmets every 30 minutes | surgical helmet systems taken from LAF OTs had no contamination, which was significantly less than those from the non-LAF theaters. After an incubation period, however, the degree of bacterial contamination was not significantly different between the LAF and non-LAF swabs. *Staphylococcus aureus* was the most commonly identified organism. | should be avoided during surgery. |
| 133 | CFD modeling | India | Number of trials not specified | One supply diffuser and four exhaust vents, inlet airflow velocity fixed at 0.4 m/s | Angle of distribution of the supply diffuser in an AAD system: 45, 60, or 90 degrees | Air velocity in the critical zone and formation of turbulence, as determined by camera images of smoke flow tests | For each angle tested, the air velocity in the critical zone varies from 0.2 to 0.3 m/s, which is within the acceptable range. However, the flat (90 degree) angle can cause turbulence in the OR. | To avoid turbulence and drafts, increase the inlet surface area of the flat diffuser, or provide angular flow over the critical zone by implementing the AAD system. |
| 134 | CFD modeling | USA | Number of trials not specified | Floor area of the OR is 15.6m$^2$ and ceiling height is 3 m; air is supplied at 18.3°C | LAF diffusers with air supply velocities of 0.13, 0.25, 0.38, and 0.51 m/s, supplied to ORs operating at isothermal or non-isothermal conditions | Airflow patterns and temperature in different regions of the OR | Under isothermal conditions, the supply velocity has little effect on airflow patterns. When heat sources are present, calculating the Archimedes number can be informative during the design stage; sustained unidirectional flow is most likely to be maintained if this number is less than one. | Recommend calculating the Archimedes number during design of the OR and HVAC layout in order to avoid LAF disruption and turbulent flow. |
| 135 | CFD modeling | Turkey | Number of trials not specified | The OR had dimensions 9.35 m (L) x 5.54 m (W) x 3 m (H) with an LAF unit measuring 3 m x 3 m | LAF function with average inlet air velocities of 0.1 m/s and 0.2 m/s at 19ºC, and initial OR temperatures of 19, 20, 21, or 22°C | Airflow velocity and average number of airborne particles | An inlet velocity of 0.1 m/s was too low to keep particle counts to an acceptable minimum. While variations in particle counts were observed between the four temperature conditions, no specific pattern was observed and the statistical significance of these differences was not described. | The temperature of inlet air from the LAF system must be a few degrees cooler than the temperature of the OR for unidirectional airflow to be maintained at an acceptable velocity. Future studies should also consider the effect of personnel presence and movement in the OR. |
| 136 | Empirical | Republic of Korea | 2,091 patients of gastric surgeries across 10 hospitals | | Use of LAF in the OR | Incidence of SSIs in the month following gastric surgery | A total of 71 SSIs were recorded. 63.4% of the surgeries that resulted in a SSI were conducted in LAF ORs, whereas | Use of LAF during gastric surgery is encouraged to lower SSI rates in Korea. |

| Reference | Study design | Location | Sample size | OR characteristic(s) | Aspect of HVAC | Outcome | Major Takeaway | Conclusion / Recommendation |
|---|---|---|---|---|---|---|---|---|
| | | | | | | | 92.8% of surgeries that did not result in an SSI were conducted in LAF ORs. This result was statistically significant. | |
| 36 | Simulation | The Netherlands | Each measurement was repeated four times | The test chamber had dimensions 6 m (L) x 6 m (W) x 3 m (H). Air was supplied through four separate plenums in the ceiling and extracted through 12 grilles in the corners of the room | Location of exhaust grills; comparing various LAF conditions against the conventional MV system at the same ventilation rate | VDI 2167 (published particle dissemination test) guidelines including protection factor, which is calculated by dividing the measured concentration of particles by the concentration in a fully MV case | The location of the exhaust grills do not influence the measured protection factor. Additionally, every LAF scenario tested offered a higher level of protection than the MV case. | Regardless of the ventilation systems, the temperature distribution in the room can affect its performance. Future studies can use the VDI method to better understand the correlation between ventilation systems and infection incidence. |
| 137 | Empirical | The Netherlands | Three ORs | All ORs were equipped with unidirectional LAF with supply air velocities of 0.25-0.28 m/s and plenum heights of 3 m | Comparing three different OTs: Theater A (built in 2010, sized at 45.6 $m^2$, plenum size 8.12 m2), Theater B (built in 2012, sized at 39.9 $m^2$, plenum size 8.29 $m^2$), and Theater C (built in 1993, sized at 37.4 $m^2$, plenum size 3.51$m^2$) | Recovery time, defined as the amount of time needed to lower the detected number of particles sized greater than 0.5 μm by a factor of 100 | Regardless of OT, an acceptable air quality level was reached within the recommended 30-minute safety margin of having initiated the ventilation system. Based on the number of measures taken in each OR, the 95% upper confidence limit was determined to be 13 minutes for Theater A, 23 minutes for Theater B, and 20 minutes for Theater C. | OR ventilation systems can be safely shut down overnight in an attempt to save on electricity, provided that the appropriate recovery time is allowed on the following morning. It is worth noting, however, that further studies are needed in order to determine the generalizability of these results to other ventilation systems and the shutdown's effect on particle sedimentation onto OR surfaces. |
| 138 | Simulation | USA | Three ORs at three different hospitals | ORs A and B are associated with medical schools, opened in 2013 and 2011 respectively; OR C is in a private community hospital opened in 2004 | ORs A and B are equipped with HEPA filters while OR C has minimum efficiency reporting value 14 filters | Air velocity, temperature, humidity, pressure, microbial counts measured in CFU/$m^3$ and particle counts for various sizes | Air velocity varied at multiple locations, between ORs A and B (equipped with HEPA filters) versus OR C, but temperature, humidity, and pressurization were similar. Microbial counts in the sterile field were significantly lower in OR A compared to ORs B and C, while OR C had significantly lower microbial counts than A and B at the back table. Counts of small particles of 0.3 to 0.5 μm were significantly higher in OR C. | Study is aimed at developing a consistent performance metric that is more informative than ventilation rates or other metrics. Results suggest the environmental quality indicators (EQI) approach is effective. |

| Reference | Study design | Location | Sample size | OR characteristic(s) | Aspect of HVAC | Outcome | Major Takeaway | Conclusion / Recommendation |
|---|---|---|---|---|---|---|---|---|
| 139 | Empirical | Sweden | Ten surgeries (including six liver resections) performed in five ORs (four equipped with UWD systems and one with UDF) | UWD ORs had an area of 37 $m^2$ and a volume of 100 m3. UDF ORs had an area of 100 $m^2$ and a volume of 270 $m^3$. The air renewal rates were 20 ACH and 57 ACH for UWD and UDF ORs, respectively. All ORs were held at a relative pressure of 10 Pa, 20°C, and a relative humidity of 55% | UWD versus UDF systems | HCW exposure to surgical smoke and UFP concentration in the air | In ORs equipped with UWD ventilation systems, the exposure to surgical smoke is 13x higher than in ORs equipped with the UDF system. During the liver resections, the concentrations of UFPs was 18.3 particles/$cm^3$ with the UWD ventilation system, whereas with the UDF system this concentration was 1.38 particles/$cm^3$. | ORs equipped with UDF ventilation systems evacuate surgical smoke from the surgical site faster and more efficiently than ORs with the UWD system. The UDF system offers a greater airflow volume and more defined airflow patterns which, coupled with well-positioned extraction grills, prevent the entrainment and recirculation of particles that has been observed with UWD systems. Large air supply volumes with sufficiently defined airflow pathways are necessary in order to adequately protect HCWs from the health risks associated with surgical smoke inhalation. |
| 140 | CFD modeling | Iran | Number of trials not specified | The OR was modeled with dimensions 7 m (W) x 7 m (L) x 3.66 m (H), giving a total volume of 179 $m^3$. The air renewal rate is modeled as 25 ACH, and the laminar diffuser velocity was 0.38 m/s in all conditions | Presence of an air curtain at different velocities (1.37 m/s, 2.37 m/s, 3.37 m/s, and 4.37 m/s) surrounding the vertical LAF system | Contamination on the operating table (units were not specified) | The lowest level of contamination at the operating table was observed when the air curtain was operating at 2.37 m/s, while the highest was observed at 1.37 m/s. | The presence of an air curtain with an optimized exhaust airflow velocity can drastically decrease contamination. Given that LAF can sometimes demonstrate unexpected behavior that leads to a disturbance of optimal sterile conditions, it can be useful to have an air curtain that helps ensure a "one pass, then exit" airflow pattern. |
| 141 | Empirical | Brazil | Two ORs, 5 surgeries per measurement period | The OR had dimensions 5 m (L) x 5.6 m (W) x 2.7 m (H) and was specialized for orthopedic surgery | Effect of changing the 16-month-old air filter on an LAF HVAC in one OR and installation of new LAF HVAC system in another | TPCs, VPCs, ACH, and $CO_2$ concentration | Changing the filter increased ACH from about 9.3 to 25.8 and decreased particle concentration by 97%, reaching levels similar to the new HVAC system, which had 27.1 ACH. All cases still remained below the recommended Brazilian | HVAC maintenance, especially regular filter replacement, is essential for maintaining acceptable airflow levels and minimizing contamination in the OR. |

| Reference | Study design | Location | Sample size | OR characteristic(s) | Aspect of HVAC | Outcome | Major Takeaway | Conclusion / Recommendation |
|---|---|---|---|---|---|---|---|---|
| | | | | | | | standard for ACH. Average VPC was decreased from 57 to 24 CFU/$m^3$ by changing the filter, but remained much higher than in the new OR at15 CFU/$m^3$. Temperature, relative humidity, and $CO_2$ concentration were not significantly affected. | |
| 142 | CFD modeling | Iran | 27 simulation cases | The half of the OR modeled measures 6.1 m (L) x 2.15 m (W) x 3 m (H) | System design of LAF | Particle deposition in the occupancy zone | To minimize mean contaminant concentration, an air supply velocity of about 0.4 m/s and an air barrier velocity of about 2 m/s is optimal. A larger LAF system is not necessarily an improvement. | Recommend an air supply velocity of 0.4 m/s and an air barrier velocity of 2 m/s for average sized LAF systems. |
| 143 | CFD modeling | Sweden | 1 modeled OR, number of trials not given | The OR was modeled with dimensions 8.5 m (L) × 7.7 m (W) × 3.2 m (H), a temperature of 20°C, and an air renewal rate of 47 ACH | Comparing two different LAF distribution strategies: horizontal and vertical | Particle sedimentation and OR recovery time | The vertical system performed worse than a horizontal configuration. Fewer particles were detected in the sedimentation areas under the horizontal LAF condition. The recovery time for the horizontal LAF system was 8.1 min, while this time increased to11.9 min in the vertical LAF scenario. | Horizontal LAF performed better than vertical. |
| 54 | Empirical | Taiwan | 250 surgical procedures performed across 28 ORs | Climate: ~19-23ºC, ~30-70% relative humidity<br>HVAC: Minimum of 40 ACH; LAF set to 0.3-0.5 µm/s<br>Particle counts: The overall mean number of bacterial colonies in the ORs was 78 ± 47 CFU/$m^3$. Before incision = 99.9 ± 55.7 CFUs/$m^3$; during = 66.9 ± 38.3 CFUs/$m^3$; after = 110 ± 46.6 CFUs/$m^3$ | Ambient temperature inside the OR | Colony count measured as CFU/$m^3$ | The colony count collected inside the OR increased by 9.4 CFU/$m^3$ with each additional 1°C, a statistically significant result. | We suggest that ORs doing complex surgeries with more surgical personnel present should increase the frequency of air exchanges. |
| 144 | Modeling | China | | The OR was modeled with dimensions 8.73 m | Comparing four different air renewal rates: 10 ACH, 15 ACH, 20 ACH, and 26 ACH | Contamination modeled using $N_2O$ gas | Increasing the air renewal rate led to a decrease in contaminant concentration. | The current commonly recommended 20 ACH should be improved given that a higher air renewal |

| Reference | Study design | Location | Sample size | OR characteristic(s) | Aspect of HVAC | Outcome | Major Takeaway | Conclusion / Recommendation |
|---|---|---|---|---|---|---|---|---|
| | | | | (L) x 7.05 m (W) x 3.25 m (H) | | | | rate could further improve OR air quality. |
| 56 | Empirical | USA | 21 procedures performed in 4 ORs (12 orthopedic procedures in one OR and 9 pediatric procedures across 3 ORs), two seasons (March and September) | | Ambient temperature and humidity inside the OR | Microbial load at various sampling locations | Ambient temperature and humidity did not have a significant impact on microbial load, regardless of the location of sampling or the season. | |
| 145 | Empirical | UK | 21 open vascular cases and 24 orthopedic major joint replacement cases | | Conventional airflow used in vascular cases compared to LAF in orthopedic cases | Bacterial colony counts collected on agar settle plates | Bacterial counts were 15-fold greater in the vascular cases using CV than in the orthopedic cases using LAF ventilation. | Consider performing vascular surgeries under LAF rather than CV. |
| 38 | Empirical | Norway | 2 ORs and 2 scenarios with 4 different cases | OR with LAF ventilation has an area of 56 $m^2$ and an LAF zone of 11 $m^2$ and is surrounded by partial walls of 1.1 m in length. OR with MV had 4 ceiling-mounted diffusers, and an area of 59.7 $m^2$ and a height of 2.90 m | To examine the disruption of airflow distribution (LAF and MV systems) while under real operating conditions | Airflow distribution, airflow velocity, airflow pattern/direction, TI | Due to disruption of the LAF, airflow velocity above the patient was significantly lower under LAF than MV. TI of the supply airflow from LAF was much lower than that of MV. | LAF systems should be calibrated for specific surgical facilities to ensure that the minimum standard for airflow velocity near the patient is achieved. |
| 146 | Empirical | Egypt | 6 ORs, 186 collected samples | | Presence of HEPA filters | Number of samples exceeding the acceptable limit of microbial contamination | While 2.1% of samples taken in ORs equipped with HEPA filters exceeded the acceptable limit, 11.3% of those taken in rooms without HEPA filters exceeded such limit. | Use of a HEPA filter improves the air quality in ORs. |
| 147 | Empirical | United Kingdom | A total of 170 vascular operations were performed during the study period: 114 in a non-LAF OT and 56 in an LAF OT; SSIs occurred in 23 patients, or 13.5% of cases; 14 | Vascular surgery; mean operation time 120-126.4 min | Comparing LAF vs non-LAF OTs: A one-year retrospective analysis was carried out on a prospectively collected database of consecutive patients undergoing open vascular procedures, both venous and arterial, performed by a single vascular surgeon | Rate of SSI | LAF may play an important role in reducing the incidences of SSIs subsequent to vascular surgery. Rates of SSI following vascular surgery were significantly higher when surgeries were not performed in ORs equipped with LAF. | Vascular surgeries should be performed in OTs equipped with LAF ventilation. |

| Reference | Study design | Location | Sample size | OR characteristic(s) | Aspect of HVAC | Outcome | Major Takeaway | Conclusion / Recommendation |
|---|---|---|---|---|---|---|---|---|
| | | | superficial infections and 9 deep infections | | | | | |

#### Portable airflow systems

**Table S7.** Summary of the 12 articles included that investigated the impact of portable airflow systems in the operating room on airflow, contaminations, and infections.

| Reference | Study design | Location | Sample size | OR characteristic(s) | Device | Outcome | Major Takeaway | Conclusion / Recommendation |
|---|---|---|---|---|---|---|---|---|
| 148 | Empirical | USA | 43 total shoulder arthroplasty cases: 21 intervention, 22 control | CV with 12-15 ACH | Air barrier system (ABS) providing localized, HEPA-filtered LAF | Number of CFUs in air sampled directly above the wound | The use of an ABS significantly reduced the number of CFUs in the air directly above the wound. | Further studies should investigate whether the ABS device can ultimately reduce rates of PJI following surgery. |
| 149 | Empirical | Sweden | Two TKAs | Dimensions: 8 m x 6 m x 3 m. OR equipped with a main air supply from ceiling of 0.3 m/s and a full-sized vertical LAF unit. Air outlets located at floor level | Use of a horizontal LAF device attached to instrument table | Particle counts and CFUs on the instrument table. These counts were taken during surgery with an extra nurse challenging the LAF systems by conducting extra movements to stimulate particle production | Addition of the horizontal LAF device to the instrument table dramatically reduced particle counts over the table and reduced CFU/m$^2$/h by more than 50%. | The addition of a horizontal LAF device to the instrument table reduces airborne contamination over the table, and can be used safely in conjunction with the main LAF unit. |
| 150 | CFD modeling | Sweden | 500 simulations | Modeled OR size: 8.5 m (L) × 7.7 m (W) × 3.2 m (H) | MLAF device used to complement TV in the OR | Simulated BCP deposition and concentration on surfaces | MLAF significantly reduced contamination as long as the rate of airflow was sufficiently fast, at about 0.4 m/s. | Surgeries with high risk of infection can safely be carried out in ORs with TV if MLAF units are also used. |
| 151 | Empirical | Italy | 34 orthopedic surgeries: 17 intervention, 17 control | The OR temperature and relative humidity were 20.6°C and 44.6%, respectively. The OR was equipped with conventional turbulent air ventilation providing 12.5 ACH | MLAF screen device positioned near the operating table, with airflow directed toward the surgical site | Bacterial counts measured in CFU/m$^3$ and counts of 0.5 µm particles, both near the wound and near the instrument table | The MLAF unit significantly decreased particle counts and also reduced mean bacterial count near the wound from 23.5 to 3.5 CFU/m$^3$, which is below the suggested limit for an OR with UCV. Bacterial counts near the instrument table were not significantly different with the use of the mobile unit. | The addition of a MLAF screen in an OR with CV could be a safe and cost-effective alternative to the installation of LAF ventilation systems. |

| Reference | Study design | Location | Sample size | OR characteristic(s) | Device | Outcome | Major Takeaway | Conclusion / Recommendation |
|---|---|---|---|---|---|---|---|---|
| 152 | Simulation | The Netherlands | 10 background samples, four samples from instrument table and surgical site at rest, and 10 samples during simulation | The mechanical ventilation system inside the OR was switched off during the simulation | Use of a mobile UDF screen | The degree of protection provided by the device, calculated from the airborne particle concentration in the operative area versus the background level | The mobile UDF device significantly reduced the concentration of larger, potentially BCPs. The protection factor at the instrument table was up to 5.0, and the protection factor in the ocular area was between 2.64 and 5.0. | Use of a mobile UDF device alone may be sufficient protection from airborne infection during intravitreal injections. |
| 153 | CFD modeling | Sweden | One OR | OR dimensions: 8.5 m (L) × 7.7 m (W) × 3.2 m (H). The supply air volume, temperature, and turbulent intensity at the ceiling inlet were taken as 2000 L/s, 20°C, and 5%, respectively. The exhausts were set with pressure outlets at 10 Pa and TI of 5% | Use of a MLAF unit in addition to the main TMV system | Particle counts above the operating and instrument tables, and sedimentation rate of BCPs measured as $CFU/m^2/h$ | The simulation showed an increase in particle counts in the absence of MLAF units during surgery. The use of MLAF units significantly decreased BCP sedimentation rates, regardless of the type of surgical clothing worn by HCWs. The sedimentation rate was decreased by 70% with the mobile units for the clothing that released the most particles. | MLAF units offer an advantage for surgeries with a high risk of infection, especially in ORs equipped with TMV. These units were successful in reducing particle counts and sedimentation rates, and have the added benefit of installation without renovation and with immediate use. |
| 59 | Empirical | USA | 36 total hip arthroplasties in two ORs | Orthopedic ORs | Use of a portable HEPA LAF device, compared to two controls: no portable device or portable device present but turned off | Airborne particle concentration and bacterial concentration, measured in $CFU/m^3$, over the surgical site | On average, particle counts decreased by at least 66%, and particles larger than 10 µm decreased by 80%. $CFUs/m^3$ were significantly reduced compared to the controls, and more than 60% of samples with the portable device had no detectable CFUs at all. | Recommend a larger and ongoing study of 1000+ patients, in order to understand the effect of the portable device on SSI rates. |
| 154 | CFD modeling | Sweden | One OR with turbulent airflow, modeled under six different MLAF conditions | OR is equipped with conventional turbulent flow | Use of an MLAF screen with air velocity at 0.0, 0.2, 0.4, 0.6, 0.8, or 1.0 m/s | Concentration of BCPs from simulated active air sampling, in $CFU/m^3$, and simulated passive air sampling, in $CFU/m^2/h$ | The concentration of BCPs decreases as air velocity from the mobile LAF increases, with the exception of settling particles in the periphery of the OR. Reductions in BCPs are modest as velocity increases beyond 0.4 m/s. | Use of MLAF can be very effective in reducing BCPs in an OR with turbulent flow, especially within about 1 m of the mobile device. |
| 155 | CFD modeling | Italy | One OR with six simulation conditions | OR is equipped with LAF diffuser in the ceiling | Addition of a mobile LAF device at 15, 42, or 69 ACH | Simulated particle deposition rate, measured in $BCP/m^3$ | Use of MLAF in this design significantly reduces BCPs above the instrument table, but by disrupting the airflow in the OR, the device | MLAF devices should not be placed in the OR without careful assessment of their impact on airflow patterns. |

| Reference | Study design | Location | Sample size | OR characteristic(s) | Device | Outcome | Major Takeaway | Conclusion / Recommendation |
|---|---|---|---|---|---|---|---|---|
| | | | | | | | actually increases BCPs above the surgical table. | |
| 156 | Empirical | Sweden | 45 neurosurgical procedures performed in three ORs. 26 surgeries were done without MLAF and 19 surgeries were done with MLAF units | OR 1: 153 $m^3$; OR 2: 175 $m^3$; OR 3: 173 $m^3$. All ORs equipped with conventional turbulent ventilation | Use of a horizontally placed MLAF unit | Airborne bacterial contamination in the surgical site area and above the instrument table, measured in CFU/$m^3$ | MLAF units significantly reduced airborne bacterial concentrations both in the surgical site area and above the instrument table. | MLAF may be a useful addition during neurosurgeries that take place in ORs equipped with conventional turbulent airflow. Further studies should evaluate whether the MLAF units have an effect on SSI rates following surgery. |
| 157 | CFD modeling | USA and China | Four alternative ventilation setups, with three variations in supply and exhaust angle and three alternative exhaust flow rates. All scenarios had conventional ceiling-mounted unidirectional ventilation (CA) | Model OR dimensions are 6.1 m x 5.8 m x 2.9 m. Modeled climate is 20°C, with an air supply temperature of 18.3°C and a room pressurization of +2.5 Pa. The HVAC system diffuser dimensions are 2.44 m x 3.05 m, for a diffuser coverage area of 7.06 $m^2$ supplying 31.6 ACH | The study compared three localized ventilation approaches used in conjunction with CA. The local ventilation approaches are ceiling air supply with two jets (CAJ), ceiling air supply with two suction units (CAS), and ceiling air supply with jet and suction (CAJS). In each case, the jet and/or suction devices are placed opposite one another across the wound area. For the CAJS system, they also test alternative device angles of 0, 45, and 90 degrees, and exhaust rates of 0.0231, 0.0462, and 0.0693 $m^3$/s | The CFD model generates 1000 or 5000 particles smaller than 0.5 µm in the wound area, depending on the simulation. The study reports predicted particle counts on the patient near the wound, on the medical personnel, and trapped in the air within the OR | The CAJS system was the most effective in preventing particles from settling on medical personnel. Particles on other surfaces and in the air are somewhat varied between treatments. Under the CAJS system, it was found that 90% of particles can be removed from the OR if the jet and exhaust are at a 90 degree angle, and that the number of particles deposited near the wound is minimized at an exhaust flow rate of 0.0463 $m^3$/s. | Local ventilation used appropriately in coordination with conventional air supply can reduce particle deposition and the number of particles trapped in the air. |
| 158 | CFD Modeling | The Netherlands | 30 particle counter measurements for each corner of the clean area for 20 minutes. Two alternative configurations of the local ventilation system | OR dimensions: 6 m x 7 m x 3 m; UDF ventilation system | Use of a ventilating blanket design during surgery, under two alternative configurations: configuration 1 with horizontal ventilation and configuration 2 with vertical ventilation | Airflow patterns and particle concentrations near the surgical wound | Particle concentrations were significantly lower for configuration 2, with vertical ventilation, compared to configuration 1, with horizontal ventilation. Neither configuration is sufficiently effective in minimizing particles in the absence of OR ventilation. | The local ventilation system could be useful in emergency or resource-limited situations. More research and improved prototypes are needed. |

### APPENDIX S5. PRISMA CHECKLIST

**Table S8.** PRISMA 2020 Checklist[159]

| Section and Topic | Item # | Checklist item | Location where item is reported |
|---|---|---|---|
| **TITLE** | | | |
| Title | 1 | Identify the report as a systematic review. | Title page |
| **ABSTRACT** | | | |
| Abstract | 2 | See the PRISMA 2020 for Abstracts checklist. | Page i |
| **INTRODUCTION** | | | |
| Rationale | 3 | Describe the rationale for the review in the context of existing knowledge. | Pages 1-3 |
| Objectives | 4 | Provide an explicit statement of the objective(s) or question(s) the review addresses. | Page 3 |
| **METHODS** | | | |
| Eligibility criteria | 5 | Specify the inclusion and exclusion criteria for the review and how studies were grouped for the syntheses. | Pages 4-6 |
| Information sources | 6 | Specify all databases, registers, websites, organisations, reference lists and other sources searched or consulted to identify studies. Specify the date when each source was last searched or consulted. | Page 4 |
| Search strategy | 7 | Present the full search strategies for all databases, registers and websites, including any filters and limits used. | Page 4 |
| Selection process | 8 | Specify the methods used to decide whether a study met the inclusion criteria of the review, including how many reviewers screened each record and each report retrieved, whether they worked independently, and if applicable, details of automation tools used in the process. | Pages 4-6 |
| Data collection process | 9 | Specify the methods used to collect data from reports, including how many reviewers collected data from each report, whether they worked independently, any processes for obtaining or confirming data from study investigators, and if applicable, details of automation tools used in the process. | Pages 5-6 |
| Data items | 10a | List and define all outcomes for which data were sought. Specify whether all results that were compatible with each outcome domain in each study were sought (e.g., for all measures, time points, analyses), and if not, the methods used to decide which results to collect. | Page 4-6 |
| | 10b | List and define all other variables for which data were sought (e.g., participant and intervention characteristics, funding sources). Describe any assumptions made about any missing or unclear information. | N/A |
| Study risk of bias assessment | 11 | Specify the methods used to assess risk of bias in the included studies, including details of the tool(s) used, how many reviewers assessed each study and whether they worked independently, and if applicable, details of automation tools used in the process. | N/A |
| Effect measures | 12 | Specify for each outcome the effect measure(s) (e.g., risk ratio, mean difference) used in the synthesis or presentation of results. | N/A |
| Synthesis methods | 13a | Describe the processes used to decide which studies were eligible for each synthesis (e.g., tabulating the study intervention characteristics and comparing against the planned groups for each synthesis (item #5)). | N/A |
| | 13b | Describe any methods required to prepare the data for presentation or synthesis, such as handling of missing summary statistics, | N/A |

| Section and Topic | Item # | Checklist item | Location where item is reported |
|---|---|---|---|
| | | or data conversions. | |
| | 13c | Describe any methods used to tabulate or visually display results of individual studies and syntheses. | Appendix S4 |
| | 13d | Describe any methods used to synthesize results and provide a rationale for the choice(s). If meta-analysis was performed, describe the model(s), method(s) to identify the presence and extent of statistical heterogeneity, and software package(s) used. | N/A |
| | 13e | Describe any methods used to explore possible causes of heterogeneity among study results (e.g., subgroup analysis, meta-regression). | N/A |
| | 13f | Describe any sensitivity analyses conducted to assess robustness of the synthesized results. | N/A |
| Reporting bias assessment | 14 | Describe any methods used to assess risk of bias due to missing results in a synthesis (arising from reporting biases). | N/A |
| Certainty assessment | 15 | Describe any methods used to assess certainty (or confidence) in the body of evidence for an outcome. | N/A |
| **RESULTS** | | | |
| Study selection | 16a | Describe the results of the search and selection process, from the number of records identified in the search to the number of studies included in the review, ideally using a flow diagram. | Figure 1 |
| | 16b | Cite studies that might appear to meet the inclusion criteria, but which were excluded, and explain why they were excluded. | Figure 1 |
| Study characteristics | 17 | Cite each included study and present its characteristics. | Appendix S4 |
| Risk of bias in studies | 18 | Present assessments of risk of bias for each included study. | N/A |
| Results of individual studies | 19 | For all outcomes, present, for each study: (a) summary statistics for each group (where appropriate) and (b) an effect estimate and its precision (e.g., confidence/credible interval), ideally using structured tables or plots. | N/A |
| Results of syntheses | 20a | For each synthesis, briefly summarise the characteristics and risk of bias among contributing studies. | N/A |
| | 20b | Present results of all statistical syntheses conducted. If meta-analysis was done, present for each the summary estimate and its precision (e.g., confidence/credible interval) and measures of statistical heterogeneity. If comparing groups, describe the direction of the effect. | N/A |
| | 20c | Present results of all investigations of possible causes of heterogeneity among study results. | N/A |
| | 20d | Present results of all sensitivity analyses conducted to assess the robustness of the synthesized results. | N/A |
| Reporting biases | 21 | Present assessments of risk of bias due to missing results (arising from reporting biases) for each synthesis assessed. | N/A |
| Certainty of evidence | 22 | Present assessments of certainty (or confidence) in the body of evidence for each outcome assessed. | N/A |
| **DISCUSSION** | | | |
| Discussion | 23a | Provide a general interpretation of the results in the context of other evidence. | Pages 7-26 |

| Section and Topic | Item # | Checklist item | Location where item is reported |
|---|---|---|---|
| | 23b | Discuss any limitations of the evidence included in the review. | Pages 25-26 |
| | 23c | Discuss any limitations of the review processes used. | Page 4 |
| | 23d | Discuss implications of the results for practice, policy, and future research. | Pages 24-26 |
| **OTHER INFORMATION** | | | |
| Registration and protocol | 24a | Provide registration information for the review, including register name and registration number, or state that the review was not registered. | The review is not registered |
| | 24b | Indicate where the review protocol can be accessed, or state that a protocol was not prepared. | A protocol was not prepared |
| | 24c | Describe and explain any amendments to information provided at registration or in the protocol. | N/A |
| Support | 25 | Describe sources of financial or non-financial support for the review, and the role of the funders or sponsors in the review. | Omitted per PLOS Medicine submission guidelines |
| Competing interests | 26 | Declare any competing interests of review authors. | None |
| Availability of data, code and other materials | 27 | Report which of the following are publicly available and where they can be found: template data collection forms; data extracted from included studies; data used for all analyses; analytic code; any other materials used in the review. | None |

**References**

1. Parvizi J, Barnes S, Shohat N, Edmiston CE. Environment of care: Is it time to reassess microbial contamination of the operating room air as a risk factor for surgical site infection in total joint arthroplasty? *Am J Infect Control*. 2017;45(11):1267-1272. doi:10.1016/j.ajic.2017.06.027
2. Aalirezaie A, Akkaya M, Barnes CL, et al. General Assembly, Prevention, Operating Room Environment: Proceedings of International Consensus on Orthopedic Infections. *J Arthroplasty*. 2019;34(2, S):S105-S115. doi:10.1016/j.arth.2018.09.060
3. Evans RP. Current concepts for clean air and total joint arthroplasty: Laminar airflow and ultraviolet radiation: A systematic review. *Clin Orthop Relat Res*. 2011;469(4):945-953. doi:10.1007/s11999-010-1688-7
4. Brown C, Owen S. An exploration on the relationship between traffic flow and the rate of surgical site infections: A literature review. *J Perioper Pr*. 2019;29(5):135-139. doi:10.1177/1750458918815550
5. Weiser MC, Moucha CS. Operating-Room Airflow Technology and Infection Prevention. *J Bone Jt Surg*. 2018;100(9):795-804. doi:10.2106/JBJS.17.00852
6. Barr SP, Topps AR, Barnes NLP, et al. Infection prevention in breast implant surgery - A review of the surgical evidence, guidelines and a checklist. *Eur J Surg Oncol*. 2016;42(5):591-603. doi:10.1016/j.ejso.2016.02.240
7. Jain S, Reed M. Laminar Air Flow Handling Systems in the Operating Room. *Surg Infect (Larchmt)*. 2019;20(2, SI):151-158. doi:10.1089/sur.2018.258
8. Pada S, TM P. Operating room myths: what is the evidence for common practices. *Curr Opin Infect Dis*. 2015;28(4):369-374. doi:10.1097/QCO.0000000000000177
9. Birgand G, Saliou P, Lucet JC. Influence of staff behavior on infectious risk in operating rooms: What is the evidence? *Infect Control Hosp Epidemiol*. 2015;36(1):93-106. doi:10.1017/ice.2014.9
10. Bischoff P, Kubilay NZ, Allegranzi B, Egger M, Gastmeier P. Effect of laminar airflow ventilation on surgical site infections: a systematic review and meta-analysis. *Lancet Infect Dis*. 2017;17(5):553-561. doi:10.1016/S1473-3099(17)30059-2
11. James M, Khan WS, Nannaparaju MR, Bhamra JS, Morgan-Jones R. Current Evidence for the Use of Laminar Flow in Reducing Infection Rates in Total Joint Arthroplasty. *Open Orthop J*. 2015;9(S2:M7):495-498. doi:10.2174/1874325001509010495
12. Spagnolo AM, Ottria G, Amicizia D, Perdelli F, Cristina ML. Operating theatre quality and prevention of surgical site infections. *J Prev Med Hyg*. 2013;54(3):131-137.
13. McHugh SM, Hill ADK, Humphreys H. Laminar airflow and the prevention of surgical site infection. More harm than good? *Surg*. 2015;13(1):52-58. doi:10.1016/j.surge.2014.10.003
14. Dobson PF, Reed MR. Prevention of infection in primary THA and TKA. *EFORT Open Rev*. 2020;5(10):604-613. doi:10.1302/2058-5241.5.200004
15. Thomas AM, Simmons MJ. The effectiveness of ultra-clean air operating theatres in the prevention of deep infection in joint arthroplasty surgery. *Bone Joint J*. 2018;100-B(10):1264-1269. doi:10.1302/0301-620X.100B10.BJJ-2018-0400.R1
16. Gastmeier P, AC B, Brandt C. Influence of laminar airflow on prosthetic joint infections: a systematic review. *J Hosp Infect*. 2012;81(2):73-78. doi:10.1016/j.jhin.2012.04.008
17. Katz JD. Control of the Environment in the Operating Room. *Anesth Analg*. 2017;125(4):1214-1218. doi:10.1213/ANE.0000000000001626
18. Bao J, Li J. The effect of type of ventilation used in the operating room and surgical site infection: A meta-analysis. *Infect Control Hosp Epidemiol*. Published online 2020:1-6. doi:10.1017/ice.2020.1316
19. Joseph A, Bayramzadeh S, Zamani Z, Rostenberg B. Safety, Performance, and Satisfaction Outcomes in the Operating Room: A Literature Review. *Heal Environ Res*

*Des J*. 2018;11(2):137-150. doi:10.1177/1937586717705107
20. Graves N, Wloch C, Wilson J, et al. A cost-effectiveness modelling study of strategies to reduce risk of infection following primary hip replacement based on a systematic review. *Heal Technol Assess*. 2016;20(54). doi:10.3310/hta20540
21. D'Amico A, Montagna MT, Caggiano G, et al. Observational study on hospital building heritage and microbiological air quality in the orthopedic operating theater: The IM.PA.C.T. Project. *Ann di Ig*. 2019;31(5):482-495. doi:10.7416/ai.2019.2309
22. De Korne DF, Van Wijngaarden JDH, Van Rooij J, Wauben LSGL, Hiddema UF, Klazinga NS. Safety by design: effects of operating room floor marking on the position of surgical devices to promote clean air flow compliance and minimise infection risks. *BMJ Qual Saf*. 2012;21(9):746-752. doi:10.1136/bmjqs-2011-000138
23. Green C, Pamplin JC, Chafin KN, Murray CK, Yun HC. Pulsed-xenon ultraviolet light disinfection in a burn unit: Impact on environmental bioburden, multidrug-resistant organism acquisition and healthcare associated infections. *Burns*. 2017;43(2):388-396. doi:10.1016/j.burns.2016.08.027
24. Prehn F, Timmermann E, Kettlitz M, Schaufler K, Günther S, Hahn V. Inactivation of airborne bacteria by plasma treatment and ionic wind for indoor air cleaning. *Plasma Process Polym*. 2020;17(9):e2000027. doi:10.1002/ppap.202000027
25. Dong C, Yuan H, Xu R, et al. Efficacy of infection control pathway in reducing postoperative infections in patients undergoing neurosurgery. *J Infect Dev Ctries*. 2020;14(1):74-79. doi:10.3855/jidc.11747
26. Curtis GL, Faour M, Jawad M, Klika AK, Barsoum WK, Higuera CA. Reduction of Particles in the Operating Room Using Ultraviolet Air Disinfection and Recirculation Units. *J Arthroplasty*. 2018;33(7):S196-S200. doi:10.1016/j.arth.2017.11.052
27. Murrell LJ, Hamilton EK, Johnson HB, Spencer M. Influence of a visible-light continuous environmental disinfection system on microbial contamination and surgical site infections in an orthopedic operating room. *Am J Infect Control*. 2019;47(7):804-810. doi:10.1016/j.ajic.2018.12.002
28. Anis HK, Curtis GL, Klika AK, et al. In-Room Ultraviolet Air Filtration Units Reduce Airborne Particles During Total Joint Arthroplasty. *J Orthop Res*. 2020;38(2):431-437. doi:10.1002/jor.24453
29. Cook TM, Piatt CJ, Barnes S, Edmiston CE. The Impact of Supplemental Intraoperative Air Decontamination on the Outcome of Total Joint Arthroplasty: A Pilot Analysis. *J Arthroplasty*. 2019;34(3):549-553. doi:10.1016/j.arth.2018.11.041
30. Kalava A, Midha M, Kurnutala LN, Schianodicola J, Yarmush JM. How clean are the overhead lights in operating rooms? *Am J Infect Control*. 2013;41(4):387-388. doi:10.1016/j.ajic.2012.04.335
31. Zoon WAC, van der Heijden MGM, Loomans MGLC, Hensen JLM. On the applicability of the laminar flow index when selecting surgical lighting. *Build Environ*. 2010;45(9):1976-1983. doi:10.1016/j.buildenv.2010.02.011
32. Al-Waked R. Effect of Ventilation Strategies on Infection Control Inside Operating Theatres. *Eng Appl Comput Fluid Mech*. 2010;4(1):1-16. doi:10.1080/19942060.2010.11015295
33. Sajadi B, Saidi MH, Ahmadi G. Computer modeling of the operating room ventilation performance in connection with surgical site infection. *Sci Iran*. 2020;27(2):704-714. doi:10.24200/sci.2018.5514.1359
34. Külpmann R, Christiansen B, Kramer A, et al. Hygiene guideline for the planning, installation, and operation of ventilation and air-conditioning systems in health-care settings - Guideline of the German Society for Hospital Hygiene (DGKH). *GMS Hyg Infect Control*. 2016;11:Doc03-Doc03. doi:10.3205/dgkh000263
35. Kai T, Ayagaki N, Setoguchi H. Influence of the Arrangement of Surgical Light Axes on

the Air Environment in Operating Rooms. *J Healthc Eng*. 2019;2019:1-8. doi:10.1155/2019/4861273
36. Zoon WAC, Loomans MGLC, Hensen JLM. Testing the effectiveness of operating room ventilation with regard to removal of airborne bacteria. *Build Environ*. 2011;46(12):2570-2577. doi:10.1016/j.buildenv.2011.06.015
37. Aganovic A, Cao G, Stenstad L-I, Skogas JG. An experimental study on the effects of positioning medical equipment on contaminant exposure of a patient in an operating room with unidirectional downflow. *Build Environ*. 2019;165. doi:10.1016/j.buildenv.2019.04.032
38. Cao G, Nilssen AM, Cheng Z, Stenstad LI, Radtke A, Skogås JG. Laminar airflow and mixing ventilation: Which is better for operating room airflow distribution near an orthopedic surgical patient? *Am J Infect Control*. 2019;47(7):737-743. doi:10.1016/j.ajic.2018.11.023
39. Aganovic A, Cao G, Stenstad LI, Skogås JG. Impact of surgical lights on the velocity distribution and airborne contamination level in an operating room with laminar airflow system. *Build Environ*. 2017;126:42-53. doi:10.1016/j.buildenv.2017.09.024
40. Cao G, Storås MCA, Aganovic A, Stenstad LI, Skogås JG. Do surgeons and surgical facilities disturb the clean air distribution close to a surgical patient in an orthopedic operating room with laminar airflow? *Am J Infect Control*. 2018;46(10):1115-1122. doi:10.1016/j.ajic.2018.03.019
41. Traversari AAL, Bottenheft C, Louman R, van Heumen SPM, Böggemann J. The effect of operating lamps on the protected area of a unidirectional down flow (UDF) system. *Heal Environ Res Des J*. 2017;10(3):40-50. doi:10.1177/1937586716671292
42. Liu P, Zhang Y, Zheng Z, Li H, Liu X. LED surgical lighting system with multiple free-form surfaces for highly sterile operating theater application. *Appl Opt*. 2014;53(16):3427-3437. doi:10.1364/AO.53.003427
43. Refaie R, Rushton P, McGovern P, et al. The effect of operating lights on laminar flow: an experimental study using neutrally buoyant helium bubbles. *Bone Joint J*. 2017;99-B(8):1061-1066. doi:10.1302/0301-620X.99B8.BJJ-2016-0581.R2
44. Sadeghian P, Wang C, Duwig C, Sadrizadeh S. Impact of surgical lamp design on the risk of surgical site infections in operating rooms with mixing and unidirectional airflow ventilation: A numerical study. *J Build Eng*. 2020;31. doi:10.1016/j.jobe.2020.101423
45. Agodi A, Auxilia F, Barchitta M, et al. Operating theatre ventilation systems and microbial air contamination in total joint replacement surgery: results of the GISIO-ISChIA study. *J Hosp Infect*. 2015;90(3):213-219. doi:10.1016/j.jhin.2015.02.014
46. Erichsen Andersson A, Petzold M, Bergh I, Karlsson J, Eriksson BI, Nilsson K. Comparison between mixed and laminar airflow systems in operating rooms and the influence of human factors: Experiences from a Swedish orthopedic center. *Am J Infect Control*. 2014;42(6):665-669. doi:10.1016/j.ajic.2014.02.001
47. Nimra A, Ali Z, Khan MN, et al. COMPARATIVE AMBIENT AND INDOOR PARTICULATE MATTER ANALYSIS OF OPERATION THEATRES OF GOVERNMENT AND PRIVATE (TRUST) HOSPITALS OF LAHORE, PAKISTAN. *J Anim PLANT Sci*. 2015;25(3, 2, SI):628-635.
48. Liang C-C, Wu F-J, Chien T-Y, et al. Effect of ventilation rate on the optimal air quality of trauma and colorectal operating rooms. *Build Environ*. 2020;169:106548. doi:10.1016/j.buildenv.2019.106548
49. El Awady MY, Abd El Rahman AT, Al Bagoury LS, Mossad IM. Air Quality in Ain Shams University Surgery Hospital. *J Egypt Soc Parasitol*. 2014;44(3):749-759. doi:10.12816/0007878
50. Rezapoor M, Alvand A, Jacek E, Paziuk T, Maltenfort MG, Parvizi J. Operating Room Traffic Increases Aerosolized Particles and Compromises the Air Quality: A Simulated

Study. *J Arthroplasty*. 2018;33(3):851-855. doi:10.1016/j.arth.2017.10.012
51. Young RS, O'Regan DJ. Cardiac surgical theatre traffic: time for traffic calming measures? *Interact Cardiovasc Thorac Surg*. 2010;10(4):526-529. doi:10.1510/icvts.2009.227116
52. Roth JA, Juchler F, Dangel M, Eckstein FS, Battegay M, Widmer AF. Frequent door openings during cardiac surgery are associated with increased risk for surgical site infection: A prospective observational study. *Clin Infect Dis*. 2019;69(2):290-294. doi:10.1093/cid/ciy879
53. Andersson AE, Bergh I, Karlsson J, Eriksson BI, Nilsson K. Traffic flow in the operating room: An explorative and descriptive study on air quality during orthopedic trauma implant surgery. *Am J Infect Control*. 2012;40(8):750-755. doi:10.1016/j.ajic.2011.09.015
54. Fu Shaw L, Chen IH, Chen CS, et al. Factors influencing microbial colonies in the air of operating rooms. *BMC Infect Dis*. 2018;18:1-8. doi:10.1186/s12879-017-2928-1
55. Wang C, Holmberg S, Sadrizadeh S. Impact of door opening on the risk of surgical site infections in an operating room with mixing ventilation. *Indoor Built Environ*. 2021;30(2):166-179. doi:10.1177/1420326X19888276
56. Taaffe K, Lee B, Ferrand Y, et al. The Influence of Traffic, Area Location, and Other Factors on Operating Room Microbial Load. *Infect Control Hosp Epidemiol*. 2018;39(4):391-397. doi:10.1017/ice.2017.323
57. Stauning MT, Bediako-Bowan A, Andersen LP, et al. Traffic flow and microbial air contamination in operating rooms at a major teaching hospital in Ghana. *J Hosp Infect*. 2018;99(3):263-270. doi:10.1016/j.jhin.2017.12.010
58. Smith EB, Raphael IJ, Maltenfort MG, Honsawek S, Dolan K, Younkins EA. The Effect of Laminar Air Flow and Door Openings on Operating Room Contamination. *J Arthroplasty*. 2013;28(9):1482-1485. doi:10.1016/j.arth.2013.06.012
59. Stocks GW, O'Connor DP, Self SD, Marcek GA, Thompson BL. Directed Air Flow to Reduce Airborne Particulate and Bacterial Contamination in the Surgical Field During Total Hip Arthroplasty. *J Arthroplasty*. 2011;26(5):771-776. doi:10.1016/j.arth.2010.07.001
60. Sadrizadeh S, Pantelic J, Sherman M, Clark J, Abouali O. Airborne particle dispersion to an operating room environment during sliding and hinged door opening. *J Infect Public Health*. 2018;11(5):631-635. doi:10.1016/j.jiph.2018.02.007
61. Villafruela JM, San José JF, Castro F, Zarzuelo A. Airflow patterns through a sliding door during opening and foot traffic in operating rooms. *Build Environ*. 2016;109:190-198. doi:10.1016/j.buildenv.2016.09.025
62. Balocco C, Petrone G, Cammarata G. Assessing the effects of sliding doors on an operating theatre climate. *Build Simul*. 2012;5(1):73-83. doi:10.1007/s12273-012-0071-x
63. Teter J, Guajardo I, Al-Rammah T, Rosson G, Perl TM, Manahan M. Assessment of operating room airflow using air particle counts and direct observation of door openings. *Am J Infect Control*. 2017;45(5):477-482. doi:10.1016/j.ajic.2016.12.018
64. Mears SC, Blanding R, Belkoff SM. Door Opening Affects Operating Room Pressure During Joint Arthroplasty. *Orthopedics*. 2015;38(11):e991-e994. doi:10.3928/01477447-20151020-07
65. Perez P, Holloway J, Ehrenfeld L, et al. Door openings in the operating room are associated with increased environmental contamination. *Am J Infect Control*. 2018;46(8):954-956. doi:10.1016/j.ajic.2018.03.005
66. Birgand G, Azevedo C, Rukly S, et al. Motion-capture system to assess intraoperative staff movements and door openings: Impact on surrogates of the infectious risk in surgery. *Infect Control Hosp Epidemiol*. 2019;40(5):566-573. doi:10.1017/ice.2019.35
67. Bediako-Bowan AAA, Mølbak K, Kurtzhals JAL, Owusu E, Debrah S, Newman MJ. Risk factors for surgical site infections in abdominal surgeries in Ghana: emphasis on the

impact of operating rooms door openings. *Epidemiol Infect*. 2020;148, e147:1-5. doi:10.1017/S0950268820001454
68. Taaffe KM, Allen RW, Fredendall LD, et al. Simulating the effects of operating room staff movement and door opening policies on microbial load. *Infect Control Hosp Epidemiol*. Published online December 21, 2020:1-5. doi:10.1017/ice.2020.1359
69. Mathijssen NMC, Hannink G, Sturm PDJ, et al. The Effect of Door Openings on Numbers of Colony Forming Units in the Operating Room during Hip Revision Surgery. *Surg Infect (Larchmt)*. 2016;17(5):535-540. doi:10.1089/sur.2015.174
70. Peters PG, Laughlin RT, Markert RJ, Nelles DB, Randall KL, Prayson MJ. Timing of C-Arm Drape Contamination. *Surg Infect (Larchmt)*. 2012;13(2):110-113. doi:10.1089/sur.2011.054
71. Balocco C, Petrone G, Cammarata G, Vitali P, Albertini R, Pasquarella C. Experimental and numerical investigation on airflow and climate in a real operating theatre under effective use conditions. *Int J Vent*. 2015;13(4):351-368. doi:10.1080/14733315.2015.11684060
72. Weiser MC, Shemesh S, Chen DD, Bronson MJ, Moucha CS. The Effect of Door Opening on Positive Pressure and Airflow in Operating Rooms. *J Am Acad Orthop Surg*. 2018;26(5):e105-e113. doi:10.5435/JAAOS-D-16-00891
73. Zhou B, Ding L, Li F, Xue K, Nielsen P V., Xu Y. Influence of opening and closing process of sliding door on interface airflow characteristic in operating room. *Build Environ*. 2018;144(August):459-473. doi:10.1016/j.buildenv.2018.08.050
74. Alsved M, Civilis A, Ekolind P, et al. Temperature-controlled airflow ventilation in operating rooms compared with laminar airflow and turbulent mixed airflow. *J Hosp Infect*. 2018;98(2):181-190. doi:10.1016/j.jhin.2017.10.013
75. Lydon GP, Ingham DB, Mourshed MM. Ultra clean ventilation system performance relating to airborne infections in operating theatres using CFD modelling. *Build Simul*. 2014;7(3):277-287. doi:10.1007/s12273-013-0145-4
76. Hirsch T, Hubert H, Fischer S, et al. Bacterial burden in the operating room: Impact of airflow systems. *Am J Infect Control*. 2012;40(7):e228-e232. doi:10.1016/j.ajic.2012.01.007
77. Yau YH, Ding LC. A case study on the air distribution in an operating room at Sarawak General Hospital Heart Centre (SGHHC) in Malaysia. *Indoor Built Environ*. 2014;23(8):1129-1141. doi:10.1177/1420326X13499359
78. Yau YH, Ding LC. A comprehensive computational fluid dynamics simulation on the air distribution in an operating room at University of Malaya Medical Centre Malaysia. *Indoor Built Environ*. 2015;24(3):355-369. doi:10.1177/1420326X13516349
79. Sadrizadeh S, Holmberg S, Tammelin A. A numerical investigation of vertical and horizontal laminar airflow ventilation in an operating room. *Build Environ*. 2014;82:517-525. doi:10.1016/j.buildenv.2014.09.013
80. Birgand G, Toupet G, Rukly S, et al. Air contamination for predicting wound contamination in clean surgery: A large multicenter study. *Am J Infect Control*. 2015;43(5):516-521. doi:10.1016/j.ajic.2015.01.026
81. Oguz R, Diab-Elschahawi M, Berger J, et al. Airborne bacterial contamination during orthopedic surgery: A randomized controlled pilot trial. *J Clin Anesth*. 2017;38:160-164. doi:10.1016/j.jclinane.2017.02.008
82. Pinder EM, Bottle A, Aylin P, Loeffler MD. Does laminar flow ventilation reduce the rate of infection? an observational study of trauma in England. *Bone Joint J*. 2016;98-B(9):1262-1269. doi:10.1302/0301-620X.9869.37184
83. Alfonso-Sanchez JL, Martinez IM, Martín-Moreno JM, González RS, Botía F. Analyzing the risk factors influencing surgical site infections: The site of environmental factors. *Can J Surg*. 2017;60(3):155-161. doi:10.1503/cjs.017916

84. Agirman A, Cetin YE, Avci M, Aydin O. Effect of laminar airflow unit diffuser size on pathogen particle distribution in an operating room. *Sci Technol Built Environ*. 2020;0(0):1-12. doi:10.1080/23744731.2020.1816405
85. Lee S-T, Liang C-C, Chien T-Y, Wu F-J, Fan K-C, Wan G-H. Effect of ventilation rate on air cleanliness and energy consumption in operation rooms at rest. *Environ Monit Assess*. 2018;190(3). doi:10.1007/s10661-018-6556-z
86. Wong KY, M. Kamar H, Kamsah N. Enhancement of Airborne Particles Removal in a Hospital Operating Room. *Int J Automot Mech Eng*. 2019;16(4):7447-7463. doi:10.15282/ijame.16.4.2019.17.0551
87. Montagna MT, Rutigliano S, Trerotoli P, et al. Evaluation of Air Contamination in Orthopaedic Operating Theatres in Hospitals in Southern Italy: The IMPACT Project. *Int J Environ Res Public Health*. 2019;16(19):3581. doi:10.3390/ijerph16193581
88. Nasir ZA, Mula V, Stokoe J, Colbeck I, Loeffler M. Evaluation of total concentration and size distribution of bacterial and fungal aerosol in healthcare built environments. *Indoor Built Environ*. 2015;24(2):269-279. doi:10.1177/1420326X13510925
89. Loth AG, Guderian DB, Haake B, Zacharowski K, Stöver T, Leinung M. Aerosol Exposure During Surgical Tracheotomy in SARS-CoV-2 Positive Patients. *Shock*. 2021;Publish Ah. doi:10.1097/SHK.0000000000001655
90. Pereira ML, Vilain R, Galvão FHF, Tribess A, Morawska L. Experimental and numerical analysis of the relationship between indoor and outdoor airborne particles in an operating room. *Indoor Built Environ*. 2013;22(6):864-875. doi:10.1177/1420326X12460707
91. Xue K, Cao G, Liu M, et al. Experimental study on the effect of exhaust airflows on the surgical environment in an operating room with mixing ventilation. *J Build Eng*. 2020;32. doi:10.1016/j.jobe.2020.101837
92. Pasquarella C, Barchitta M, D'Alessandro D, et al. Heating, ventilation and air conditioning (HVAC) system, microbial air contamination and surgical site infection in hip and knee arthroplasties: The GISIO-SItI Ischia study. *Ann di Ig*. 2018;30(5 Suppl. 2):22-35. doi:10.7416/ai.2018.2248
93. Shirozu K, Setoguchi H, Araki K, Ando T, Yamaura K. Impact of air-conditioner outlet layout on the upward airflow induced by forced air warming in operating rooms. *Am J Infect Control*. 2021;49(1):44-49. doi:10.1016/j.ajic.2020.06.202
94. Diab-Elschahawi M, Berger J, Blacky A, et al. Impact of different-sized laminar air flow versus no laminar air flow on bacterial counts in the operating room during orthopedic surgery. *Am J Infect Control*. 2011;39(7):e25-e29. doi:10.1016/j.ajic.2010.10.035
95. Barbadoro P, Bruschi R, Martini E, et al. Impact of laminar air flow on operating room contamination, and surgical wound infection rates in clean and contaminated surgery. *Eur J Surg Oncol*. 2016;42(11):1756-1758. doi:10.1016/j.ejso.2016.06.409
96. Agirman A, Cetin YE, Avci M, Aydin O. Effect of air exhaust location on surgical site particle distribution in an operating room. *Build Simul*. 2020;13(5, SI):979-988. doi:10.1007/s12273-020-0642-1
97. Gormley T, Markel TA, Jones H, et al. Cost-benefit analysis of different air change rates in an operating room environment. *Am J Infect Control*. 2017;45(12):1318-1323. doi:10.1016/j.ajic.2017.07.024
98. Traversari AAL, van Heumen SPM, van Tiem FLJ, Bottenheft C, Hinkema MJ. Design variables with significant effect on system performance of unidirectional displacement airflow systems in hospitals. *J Hosp Infect*. 2019;103(1):e81-e87. doi:10.1016/j.jhin.2019.03.009
99. Wagner JA, Greeley DG, Gormley TC, Markel TA. Comparison of operating room air distribution systems using the environmental quality indicator method of dynamic simulated surgical procedures. *Am J Infect Control*. 2019;47(1):e1-e6. doi:10.1016/j.ajic.2018.07.020

100. Din A, Foden P, Mathew M, Periasamy K. Does laminar flow reduce the risk of early surgical site infection in hip fracture patients? *J Orthop*. 2020;18:13-15. doi:10.1016/j.jor.2019.08.026
101. Baracat TM, da Silva CA, Lofrano FC, Kurokawa FA. Assessment of the performance of airflow in an operating rooms using ceiling supply and sidewall inlet systems. *J BRAZILIAN Soc Mech Sci Eng*. 2020;42(41):1-13. doi:10.1007/s40430-019-2117-9
102. Khankari K. Computational Fluid Dynamics (CFD) Analysis of Hospital Operating Room Ventilation Systems - Part I: Analysis of Air Change Rates. *ASHRAE J*. 2018;60(5):14-26.
103. Khankari K. Computational Fluid Dynamics (CFD) Analysis of Hospital Operating Room Ventilation Systems - Part II: Analyses of HVAC Configurations. *ASHRAE J*. 2018;60(6):16-26.
104. Chidambaram S, Vasudevan MC, Nair MN, Joyce C, Germanwala A V. Impact of Operating Room Environment on Postoperative Central Nervous System Infection in a Resource-Limited Neurosurgical Center in South Asia. *World Neurosurg*. 2018;110:e239-e244. doi:10.1016/j.wneu.2017.10.142
105. Vonci N, De Marco MF, Grasso A, Spataro G, Cevenini G, Messina G. Association between air changes and airborne microbial contamination in operating rooms. *J Infect Public Health*. 2019;12(6):827-830. doi:10.1016/j.jiph.2019.05.010
106. Agarwal SK, Khan AA, Solan M, Lemon M. Hip fracture surgery in mixed-use emergency theatres: is the infection risk increased? A retrospective matched cohort study. *Ann R Coll Surg Engl*. 2017;99(8):641-644. doi:10.1308/rcsann.2017.0183
107. Wagner JA, Schreiber KJ, Cohen R. Using Cleanroom Technology: Improving Operating Room Contamination Control. *ASHRAE J*. 2014;56(2):18-27.
108. Wagner JA, Dexter F, Greeley DG, Schreiber K. Operating room air delivery design to protect patient and surgical site results in particles released at surgical table having greater concentration along walls of the room than at the instrument tray. *Am J Infect Control*. 2020;000:1-4. doi:10.1016/j.ajic.2020.10.003
109. Langvatn H, Schrama JC, Cao G, et al. Operating room ventilation and the risk of revision due to infection after total hip arthroplasty: assessment of validated data in the Norwegian Arthroplasty Register. *J Hosp Infect*. 2020;105(2):216-224. doi:10.1016/j.jhin.2020.04.010
110. Romano F, Milani S, Ricci R, Joppolo CM. Operating Theatre Ventilation Systems and Their Performance in Contamination Control: “At Rest” and “In Operation” Particle and Microbial Measurements Made in an Italian Large and Multi-Year Inspection Campaign. *Int J Environ Res Public Health*. 2020;17(19):7275. doi:10.3390/ijerph17197275
111. Bahador M, Keshtkar MM. Reviewing and modeling the optimal output velocity of slot linear diffusers to reduce air contamination in the surgical site of operating rooms. *Int J Comput Sci Netw Secur*. 2017;17(8):82-89.
112. Srivastava S, Vasavada V, Vasavada AR, Sudhalkar A, Kothari A, Vasavada SA. Realtime Imaging of Airflow Patterns and Impact of Infection Control Measures in Ophthalmic Practice. *J Cataract Refract Surg*. 2020;Publish Ah. doi:10.1097/j.jcrs.0000000000000538
113. Breier A-C, Brandt C, Sohr D, Geffers C, Gastmeier P. Laminar Airflow Ceiling Size: No Impact on Infection Rates Following Hip and Knee Prosthesis. *Infect Control Hosp Epidemiol*. 2011;32(11):1097-1102. doi:10.1086/662182
114. Chien T-Y, Liang C-C, Wu F-J, Chen C-T, Pan T-H, Wan G-H. Comparative Analysis of Energy Consumption, Indoor Thermal-Hygrometric Conditions, and Air Quality for HVAC, LDAC, and RDAC Systems Used in Operating Rooms. *Appl Sci*. 2020;10(11):1-16. doi:10.3390/app10113721
115. Abed IM, Amer R. Modeling and Experimental Investigation of Laminar Ceiling Air Distribution System for Operating Room in Merjan Teaching Hospital. *J Eng Technol Sci*.

2018;50(6):870-883. doi:10.5614/j.eng.technol.sci.2018.50.6.9
116. Squeri R, Genovese C, Trimarchi G, et al. Nine years of microbiological air monitoring in the operating theatres of a university hospital in Southern Italy. *Ann DI Ig Med Prev E DI COMUNITA*. 2019;31(1, SI):1-12. doi:10.7416/ai.2019.2272
117. Sajadi B, Saidi MH, Ahmadi G. Numerical evaluation of the operating room ventilation performance: Ultra-Clean Ventilation (UCV) systems. *Sci Iran*. 2019;26(4):2394-2406. doi:10.24200/sci.2018.5431.1269
118. Wang C, Holmberg S, Sadrizadeh S. Numerical study of temperature-controlled airflow in comparison with turbulent mixing and laminar airflow for operating room ventilation. *Build Environ*. 2018;144:45-56. doi:10.1016/j.buildenv.2018.08.010
119. Kirschbaum S, Hommel H, Strache P, Horn R, Falk R, Perka C. Laminar air flow reduces particle load in TKA-even outside the LAF panel: a prospective, randomized cohort study. *Knee Surgery, Sport Traumatol Arthrosc*. Published online 2020:1-9. doi:10.1007/s00167-020-06344-3
120. Teo BJX, Woo YL, Phua JKS, Chong H-C, Yeo W, Tan AHC. Laminar flow does not affect risk of prosthetic joint infection after primary total knee replacement in Asian patients. *J Hosp Infect*. 2020;104(3):305-308. doi:10.1016/j.jhin.2019.12.014
121. Hooper GJ, Rothwell AG, Frampton C, Wyatt MC. Does the use of laminar flow and space suits reduce early deep infection after total hip and knee replacement?: the ten-year results of the New Zealand Joint Registry. *J BONE Jt SURGERY-BRITISH Vol*. 2011;93-B(1):85-90. doi:10.1302/0301-620X.93B1.24862
122. Ufat H, Kaynakli O, Yamankaradeniz N, Yamankaradeniz R. Three-dimensional air distribution analysis of different outflow typed operating rooms at different inlet velocities and room temperatures. *Adv Mech Eng*. 2017;9(7):1-12. doi:10.1177/1687814017707414
123. Balocco C, Petrone G, Cammarata G. Thermo-fluid dynamics analysis and air quality for different ventilation patterns in an operating theatre. *Int J Heat Technol*. 2015;33(4):25-32. doi:10.18280/ijht.330404
124. Albertini R, Colucci ME, Turchi S, Vitali P. The management of air contamination control in operating theaters: the experience of the Parma University Hospital (IT). *Aerobiologia (Bologna)*. 2020;36(1, SI):119-123. doi:10.1007/s10453-019-09572-4
125. Zhai ZJ, Osborne AL. Simulation-based feasibility study of improved air conditioning systems for hospital operating room. *Front Archit Res*. 2013;2(4):468-475. doi:10.1016/j.foar.2013.09.003
126. Fischer S, Thieves M, Hirsch T, et al. Reduction of airborne bacterial burden in the OR by installation of unidirectional displacement airflow (UDF) systems. *Med Sci Monit*. 2015;21:2367-2374. doi:10.12659/MSM.894251
127. Yunusa U, Golfa T, Dathini H. Prognosticators of surgical site infections (SSIs) among patients undergoing major surgery at general hospital Funtua, Katsina State, Nigeria. *Pielęgniarstwo Chir i Angiol*. 2015;2:111-117.
128. Liu Z, Liu H, Yin H, Rong R, Cao G, Deng Q. Prevention of surgical site infection under different ventilation systems in operating room environment. *Front Environ Sci Eng*. 2021;15(3):36. doi:10.1007/s11783-020-1327-9
129. Wang F, Hung J, Chen Y, Hsu C. Performance evaluation for operation rooms by numerical simulation and field measurement. *Int J Vent*. 2017;16(3):189-199. doi:10.1080/14733315.2017.1299515
130. Smith JO, Frampton CMA, Hooper GJ, Young SW. The Impact of Patient and Surgical Factors on the Rate of Postoperative Infection After Total Hip Arthroplasty—A New Zealand Joint Registry Study. *J Arthroplasty*. 2018;33(6):1884-1890. doi:10.1016/j.arth.2018.01.021
131. Romano F, Milani S, Gustén J, Joppolo CM. Surgical Smoke and Airborne Microbial Contamination in Operating Theatres: Influence of Ventilation and Surgical Phases. *Int J*

*Environ Res Public Health*. 2020;17(15):5395. doi:10.3390/ijerph17155395
132. Singh VK, Hussain S, Javed S, Singh I, Mulla R, Kalairajah Y. Sterile surgical helmet system in elective total hip and knee arthroplasty. *J Orthop Surg (Hong Kong)*. 2011;19(2):234-237. doi:10.1177/230949901101900222
133. Rahate SD, Sarode AD. Design of Air Distribution System for Operation Theatre Using Flow Visualization Techniques to Improve Flow Characteristics. *Int J Eng*. 2020;33(1):164-169. doi:10.5829/ije.2020.33.01a.19
134. Khankari K. Dynamics of unidirectional airflow. *ASHRAE J*. 2019;61(7):20-39.
135. Ufat H, Kaynakli O, Yamankaradeniz N, Yamankaradeniz R. Investigation of the number of particles in an operating room at different ambient temperatures and inlet velocities. *Int J Vent*. 2018;17(3):209-223. doi:10.1080/14733315.2017.1392107
136. Jeong SJ, Ann HW, Kim JK, et al. Incidence and risk factors for surgical site infection after gastric surgery: A multicenter prospective cohort study. *Infect Chemother*. 2013;45(4):422-430. doi:10.3947/ic.2013.45.4.422
137. Traversari AAL, Bottenheft C, van Heumen SPM, Goedhart CA, Vos MC. Effect of switching off unidirectional downflow systems of operating theaters during prolonged inactivity on the period before the operating theater can safely be used. *Am J Infect Control*. 2017;45(2):139-144. doi:10.1016/j.ajic.2016.07.019
138. Gormley T, Markel TA, Jones HW, et al. Methodology for analyzing environmental quality indicators in a dynamic operating room environment. *Am J Infect Control*. 2017;45(4):354-359. doi:10.1016/j.ajic.2016.11.001
139. Romano F, Gusten J, De Antonellis S, Joppolo CM. Electrosurgical Smoke: Ultrafine Particle Measurements and Work Environment Quality in Different Operating Theatres. *Int J Environ Res Public Health*. 2017;14(2). doi:10.3390/ijerph14020137
140. Keshtkar MM, Nafteh M. Investigation of influence of linear diffuser in the ventilation of operating rooms. *Adv ENERGY Res*. 2016;4(3):239-253. doi:10.12989/eri.2016.4.3.239
141. Pereira ML, Vilain R, Kawase PR, Tribess A, Morawska L. Impact of Filtration Conditions on Air Quality in an Operating Room. *Int J Environ Res*. 2020;14(6):685-692. doi:10.1007/s41742-020-00286-x
142. Amiraslanpour M, Ghazanfarian J, Nabaei H, Taleghani MH. Evaluation of laminar airflow heating, ventilation, and air conditioning system for particle dispersion control in operating room including staffs: A non-Boussinesq Lagrangian study. *J Build Phys*. 2020;00(0):1-29. doi:10.1177/1744259120932932
143. Sadrizadeh S, Holmberg S. Surgical clothing systems in laminar airflow operating room: a numerical assessment. *J Infect Public Health*. 2014;7(6):508-516. doi:10.1016/j.jiph.2014.07.011
144. Zhang Y, Cao G, Feng G, et al. The impact of air change rate on the air quality of surgical microenvironment in an operating room with mixing ventilation. *J Build Eng*. 2020;32. doi:10.1016/j.jobe.2020.101770
145. Stather P, Salji M, Hassan SU, et al. A comparison of airborne bacterial fallout between orthopaedic and vascular surgery. *Ann R Coll Surg Engl*. 2017;99(4):295-298. doi:10.1308/rcsann.2016.0352
146. Elnour AA, Abdelfattah MM, Negm S, Kassim T. Microbiological Surveillance of Air Quality: A comparative Study Using Active and Passive Methods in Operative Theater. *Int J Pharm Phytopharm Res*. 2018;8(1):33-38.
147. Bosanquet D, Jones CN, Gill N, Jarvis P, Lewis MH. Laminar flow reduces cases of surgical site infections in vascular patients. *Ann R Coll Surg Engl*. 2013;95(1):15-19. doi:10.1308/003588413X13511609956011
148. Morris BJ, Kiser CJ, Laughlin MS, et al. A localized laminar flow device decreases airborne particulates during shoulder arthroplasty: a randomized controlled trial. *J Shoulder Elb Surg*. 2021;30(3):580-586. doi:10.1016/j.jse.2020.08.035

149. Nilsson K, Lundholm R, Friberg S. Assessment of horizontal laminar air flow instrument table for additional ultraclean space during surgery. *J Hosp Infect*. 2010;76(3):243-246. doi:10.1016/j.jhin.2010.05.016
150. Sadrizadeh S, Holmberg S. Effect of a portable ultra-clean exponential airflow unit on the particle distribution in an operating room. *PARTICUOLOGY*. 2015;18:170-178. doi:10.1016/j.partic.2014.06.002
151. Sossai D, Dagnino G, Sanguineti F, Franchin F. Mobile laminar air flow screen for additional operating room ventilation: reduction of intraoperative bacterial contamination during total knee arthroplasty. *J Orthop Traumatol*. 2011;12(4):207-211. doi:10.1007/s10195-011-0168-5
152. Lapid-Gortzak R, Traversari R, van der Linden JW, Lesnik Oberstein SY, Lapid O, Schlingemann RO. Mobile ultra-clean unidirectional airflow screen reduces air contamination in a simulated setting for intra-vitreal injection. *Int Ophthalmol*. 2017;37(1):131-137. doi:10.1007/s10792-016-0236-1
153. Sadrizadeh S, Holmberg S, Nielsen P V. Three distinct surgical clothing systems in a turbulent mixing operating room equipped with mobile ultraclean laminar airflow screen: A numerical evaluation. *Sci Technol Built Environ*. 2016;22(3):337-345. doi:10.1080/23744731.2015.1113838
154. Sadrizadeh S, Tammelin A, Nielsen P V, Holmberg S. Does a mobile laminar airflow screen reduce bacterial contamination in the operating room? A numerical study using computational fluid dynamics technique. *Patient Saf Surg*. 2014;8(27). doi:10.1186/1754-9493-8-27
155. Casagrande D, Piller M. Conflicting effects of a portable ultra-clean airflow unit on the sterility of operating rooms: A numerical investigation. *Build Environ*. 2020;171. doi:10.1016/j.buildenv.2020.106643
156. von Vogelsang A-C, Förander P, Arvidsson M, Löwenhielm P. Effect of mobile laminar airflow units on airborne bacterial contamination during neurosurgical procedures. *J Hosp Infect*. 2018;99(3):271-278. doi:10.1016/j.jhin.2018.03.024
157. Li H, Zhong K, Zhai Z (John). Investigating the influences of ventilation on the fate of particles generated by patient and medical staff in operating room. *Build Environ*. 2020;180:107038. doi:10.1016/j.buildenv.2020.107038
158. Loomans MGLC, de Visser IM, Loogman JGH, Kort HSM. Alternative ventilation system for operating theaters: Parameter study and full-scale assessment of the performance of a local ventilation system. *Build Environ*. 2016;102:26-38. doi:10.1016/j.buildenv.2016.03.012
159. Page M, McKenzie J, Bossuyt P, Boutron I, Hoffmann T, Mulrow C. The PRISMA 2020 statement: an updated guideline for reporting systematic reviews. *BMJ*. 2021;372(71). doi:https://doi.org/10.1136/bmj.n71
